# COVID-19 effective reproductive ratio determination: An application, and analysis of issues and influential factors

**DOI:** 10.1101/2020.07.15.20154039

**Authors:** Luis Alfredo Bautista Baibás, Mario Gil Conesa, Gil Rodríguez Caravaca, Blanca Bautista Baibás

## Abstract

An essential indicator of COVID-19 transmission is the effective reproduction number (*R_t_*), the number of cases which an infected individual is expected to infect at a particular point in time; curves of the evolution of *R_t_* over time (transmission curves) reflect the impact of preventive measures and whether an epidemic is controlled. We have created a Shiny/R web application (https://alfredob.shinyapps.io/estRO/) with user-selectable features: open data sources with daily COVID-19 incidences from all countries and many regions, customizable preprocessing options (smoothing, proportional increment, backwards distribution of negative corrections, etc), different MonteCarlo-Markov-Chain estimates of the generation time or serial interval distributions and state-of-the-art *R_t_* estimation frameworks (EpiEstim, R0). We have analyzed the impact of these factors in the obtained transmission curves. We also have obtained curves at the national and sub-national level and analyzed the impact of epidemic control strategies, superspreading events, socioeconomic factors and outbreaks.

We conclude that country wealth and, to a lesser extent, mitigation strategies, were associated with poorer epidemic control. Dataset quality was an important factor, and sometimes dictated the necessity of time series smoothing. We couldn’t find conclusive evidence regarding the impact of alleged superspreading events. In the reopening phase, outbreaks had an impact on transmission curves. This application could be used interactively as a tool both to obtain transmission estimates and to perform interactive sensitivity analysis.

## 1 Introduction

The *basic reproduction number* or basic reproductive ratio, *R_0_*, is the average number of secondary cases of disease caused by a single infected individual over his infectious period, when all individuals are susceptible to infection; some authors also include the absence of preventive measures in this definition. It depends on the transmissibility (probability of infection, given contact), the average rate of contacts and the average duration of the disease. Although it can be considered as intrinsic to a particular infectious disease, the contact rate is influenced by demography, population density, cultural habits, seasonality and other factors; so this parameter can vary widely between different studies (1). The *R_0_* can be also understood in terms of the SIR Epidemic models (and other compartmental models): the *R_0_* is the ratio between the effective contact rate *β* and the removal rate *ν*, and this family of models have been used to estimate it (2)(3).

The *effective reproduction number* (*R_t_*) is the actual number of cases that an infected individual is expected to infect at a particular point in time, given the current state of the population and the implemented measures, and it typically smaller than *R_0_. R_t_* can be analyzed as a function of mobility restrictions and confinement measures(4). *R_t_* also decreases when the proportion of susceptible individuals declines.

Estimating the COVID-19 reproduction number is important to get direct knowledge about the evolution of the pandemic and implement public health regulations and preventive measures. Public health strategies include infection prevention (masks, hand-washing…), social distancing (including schools closures, teleworking, physical distancing, cancellation of mass gatherings…), travel-related measures (travel advice and/or restrictions, area quarantines/”cordon sanitaire”, traveller screening…), environmental cleaning, etc… (5). According to their objective, public health strategies can be classified in mitigation and suppression. The **mitigation** strategy aims to slow down the transmission in order to prevent healthcare system collapse, without necessarily stopping epidemic spread, thereby reaching *R* values close to 1. Mitigation measures include case isolation, contacts quarantine and social distancing for vulnerable groups. On the other hand,the **suppression** strategy aims to reduce the *R* values below 1, including measures such as social distancing for all groups, household quarantine and closing education institutions (6)(7). According to their content and epidemic phase, strategies can also be classified (anticipation, early detection, containment, control and mitigation, and elimination). **Containment** can be applied when there is localized-transmission, it aims to minimize the transmission risk, emphasizing epidemiological investigation, early detection and quarantine of cases and requires significant resources (isolation, airborne protection, etc). The appearance of many cases outside the containment areas (disease amplification) renders this approach ineffective(8); and countries might have to switch to **control and mitigation** strategies that aim to reduce both the socioeconomic disruptions and the health impact(9).

On 2-Mar-2020 the European CDC published an update that included a 5-scenario analysis (0 to 4); with scenario-bound proposed measures and objectives ranging from containment to mitigation(5). A report by the Imperial College COVID-19 Response Team on 16-Mar-2020 remarked that the preferred policy option, for countries able to attain it, was suppression instead of mitigation because a mitigated epidemic would result in thousands of deaths(7). In March and April 2020 the epidemic outbreak materialized in Europe; 30 out of 31 European Union/European Economic Area and UK (EU/EEA) countries implemented public space closures, 17 out of 31 had mandatory stay-at-home policies for the general population and all countries canceled mass gatherings and closed higher education or secondary schools (10).

After the outbreak was controlled, many countries have implemented multistage dynamic scenarios (the “New Normality” in Spain(ll), the phased recovery in UK(12), the regional semaphore plans in Mexico(13), etc). Both the lockdown and the reopening phases require different transmission indicators, including the reproduction numbers(14). For instance, the French Government has defined three *R* levels: green (<1), orange (1-1.5) or red (>1.5). In the Russian Federation the *R_t_* has been a main indicator, along with bed availability and test capacity; proposed cutoffs for the *R_t_* include ≤ 1 to advance to phase I, ≤ 0.8 to advance to phase II and ≤ 0.5 to advance to phase III (15). The WHO recognized a *R_t_* as the best indication that an epidemic is controlled and declining, and recommended large countries should estimate it at subnational level(16).

An important component of transmission are superspreading patients (or events), those who allegedly transmit an infection to a large number of individuals, due to factors like high viral load, asymptomatic cases and extensive social interactions (17). Mass media has reported several events that could qualify as such: The football match between Atalanta and Valencia FC in February, 19(18), the 8 March events in Spain (19) (20) and other countries (including International Women’s Day demonstrations), the annual gathering of the Christian Open Door Church in France (14-24 February) in Mulhouse, France(21), the Spring Break in the US (varying dates depending on the college/institution, peaking March 7-14)(22) and the riots in USA after the death of George Floyd. Superspreading events are associated with both explosive growth early in an outbreak and sustained transmission in later stages, and small delays in their prevention can be associated with significant morbidity(23). In the SARS outbreak in Hong Kong in 2003 the estimated *R_t_* peaks coincided with known superspreading events (24).

Knowing the generation time is particularly important to estimate the reproduction number; the generation time (or generation interval) is the period between infection events in an infection sequence, that is to say, the period between the start of the infection in the primary patient (the infector) and the start of the infection in the individual receiving the infection (the infectee). As the generation time is not easily determined, the serial interval is often used as a surrogate(25). The serial interval (*si)* is the interval between the onset of symptoms in the infector and the onset of symptoms in the infectee; it has the same mean as the generation time, but larger variance due to the presence of asymptomatic transmission during the incubation period(26). It might become negative if asymptomatic transmission takes place and symptoms start earlier in the infectee than in the infector. Coarse data (i.e. data in which the onset of the symptoms is not precisely known in either the infector or the infectee) can still be used to estimate the incubation period(27).

Several descriptions of the COVID-19 serial interval have been made available. Nishiura et al collected information from 28 research articles and case investigation reports and described a serial interval with median 4.0 that was fitted to several distributions including a log-normal distribution; authors noted that the serial interval was shorter than incubation period due to presymptomatic transmission(28). Zhao et al used data from 21 transmission chains in Hong Kong in late January and February 2020(29). Du et al used data from 468 online reports from China in order to estimate the serial interval (30). 12.6% of case reports had negative serial intervals, i.e. the symptoms stared earlier in the infected than in the infectee and the authors attributed this to presymptomatic transmission and symptoms starting in the infectee earlier than in the infector. Estimations also exist for the generation time, obtained from clusters from Singapore (91 cases, 48% asymptomatic transmission) and Tianjin (105 cases, 62% asymptomatic transmission) by Ganyani et al(26). It is not clear how different intervals might influence the *R_t_* estimates and the practical information obtained from the *R_t_* curves.

Mathematical relationships between generation times, epidemic growth rates and reproductive numbers have been described elsewhere(31)(32)(33). In time-since-infection-models, the Incidence rate /(t) changes depending on the transmissibility *β*(*t*, *τ*) where t denotes calendar time and *tau* denotes time since infection. (Transmission is defined as a Poisson process such that an individual has a probability *β*(*t*, *τ*)*δ* of infecting another individual). Infectiousness can be decomposed into the product of *β*(*t*, *τ*) = *R(t)w(τ*) where *w*(*τ*) is the idealized generation time distribution (i.e. the probability distribution of new infection events as a function of time since infection t, assuming contact rates are constant and not time-dependent) and *β*(t) is the instantaneous reproduction number. Different estimates exist for the time-varying effective reproduction numbers(32): The **case reproduction number** at time *t* (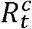), is the average number of cases an infected individual at time *t* will infect; the 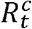 can be estimated with the method developed by Wallinga and Teunis(33) and it reflects transmissibility after time t. On the other hand, the **instantaneous reproduction number** *R_t_* is the average number of people some individual infected at time t could be expected to infect if conditions remain unchanged; it is estimated by the ratio of new infections generated at time step t, to the total infectiousness of infected individuals at time *t*(24), which is defined by 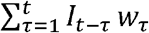.

In this report we focus on the effective reproduction number estimates, and the curves of time-varying effective reproduction number vs time (**transmission curves**, as an analog of epidemic curves which display daily incidence data). We attempt to evaluate the nuances of calculating the effective reproduction number with modern methodologies and the factors influencing this estimation, and how transmission curves have evolved in the last months.

## 2 Materials and methods

A web application in R/Shiny has been developed in order to estimate effective reproductive ratios (https://alfredob.shinyapps.io/estRO/). This application leverages and combines state-of-the-art reproductive ratio estimation tools, open COVID-19 incidence datasets, generation time/serial interval descriptions and the Shiny framework for web development, which is a R package that allows users to easily build and host interactive web applications (34).

It uses several COVID-19 open data sources: COVID-19 Data Repository by the Center for Systems Science and Engineering (CSSE) at Johns Hopkins University, European CDC COVID-19 data, Narrativa COVID-19 tracking project, the New York Times COVID-19 data source, the Spanish Public Health Surveillance (ReNaVE) COVID-19 dataset, the Brazil Covid dataset(35), the official Ministry of Health/Government COVID-19 datasets from Belgium, Canada, Chile, Colombia, Italy, Spain and Mexico, and the Catalonia Open data catalog COVID-19 sources. In many countries, volunteers perform a compilation and curation of daily reports carried out by health authorities; these unofficial/semiofficial sources of data have been included: United Kingdom (Tom White, Unlicense), Germany (J Gehrcke, MIT license), Perú (JM Castagnetto, MIT license), Portugal (Data Science for the Social Good, GPL 3.0), etc. Proper credit is given to these authors in the app.

COVID-19 cases are defined here by having a positive PCR. Datasets can have many problems or issues: data corrections or consolidations (large peaks or valleys), noise (small peaks and variability, possibly arising from a small number of tests performed), an uncertain and time-dependent proportion of undetected cases, delays between case diagnosis and report, etc. Health authorities often report cumulative incidence every day (“number of cases so far”); in this case a time series of approximate daily incidences is obtained by differentiation. As a consequence; this might introduce spurious peaks if the cumulative report includes both new cases and previous cases that were not reported, or negative values if corrections of previous reports are performed. The data quality of some COVID-19 datasets has been examined, along with the impact on *R_t_* estimations, as explained later in this section.

Graphs and tables have been implemented in the application so that the user can examine the dataset before and after preprocessing, before drawing any conclusion.

Several preprocessing steps were implemented:

- The total number of cases can-be increased by a user-defined constant proportion (ranging from 0-99%) to account for a constant proportion of undetected cases (the **undetected rate**). In some studies this proportion of undetected infections has been described to be 80-90% in Iceland (36), 72% in Italy(37) or 56.5% in Austria (38).
- The epidemic start can be defined as the date with at least an user-defined number of cases per three day period. This is important, as calculating the *R_t_* too early in the epidemic does not provide accurate or useful estimates.
- Smoothing can be performed in a variety of ways: Lowess (locally-weighted polynomial regression) and moving averaging (EMA, WMA, SMA, etc). In the Lowess smoothing, the number of points that influence each point can be adjusted (the smoothing span). Mean averaging has been tried and implemented using the previous points, the future points or both for averaging.
- Negative incidences are invalid by definition and reveal an underlying incidence reporting problem; in some series these values reflect data verification and consolidation, implying that these values are meant to compensate cases counted more than one time. This situation might arise for example if an authority reports the number of positive tests every day and then groups them by individual patients every week; or if an authority aggregates data from several institutions every day without considering whether a positive case was previously tested in two institutions and reported twice. Centralized databases of non-aggregate data could solve these issues. In our application these values can be eliminated or set to 0 before or after smoothing, or they are assumed to be periodic corrections to the case series and a **backwards distribution of negative values before smoothing** (BDNV) is performed. In the BDNV correction, we propose to smooth them with two assumptions: 1) The corrections *C_j_, C_l_* are negative values at time points *j, l* of a time series (7)); they are meant to correct the data points since the previous correction (or the beginning of the time series, if no previous correction is found). 2) The correction should be applied in a direct proportion to the incidence at each time point. (Therefore, to apply the correction *C_l_* each positive point *T_k_* between the two corrections is divided by the total sum of the points between the corrections, and multiplied by their total sum plus the signed negative correction: 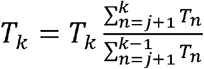. The correction point is set to 0.
- Backwards distribution of spurious peaks can also be chosen by the user. A simple peak detector has also been implemented, and it can be applied before smoothing. We arbitrarily define **spurious peaks** as daily incidences greater than 200; and 300% higher than both the two previous and the two following daily incidences; the definition also requires that the two previous and the two following daily incidences are higher than 75. In this case, the corrected spurious peak *S_jc_* at time *j* is set to the centered mean average of 5 values, including the original peak value *S_j_*, 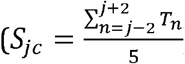) and the value to distribute backwards is the difference between the peak and the mean, 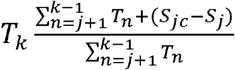).

These parameters are implemented in a Shiny app, so the user can immediately see the effect they have on the daily incidence time series; and choose the ones that better fit the data. Unless otherwise specified, we have used epidemic start after 15 cases/3 days, Simple moving average for smoothing, BDNV, no detection of positive spurious peaks and no proportional correction for undetected cases; the epidemic curves after preprocessing are displayed.

Different methods can be used to estimate the time-varying reproduction number. Wallinga and Teunis method estimate the case reproduction number at time *t* (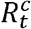), which is the number of cases an infected individual at time *t* will infect. Anne Cori et al described a method to estimate the instantaneous reproduction number *R_t_* as the number of new infections at time step *t* divided by the total infectiousness at that time step(24).

Deriving the reproduction number from incidence data requires knowledge of the generation time or the serial interval. The original serial interval datasets from Nishiura et al (28), from Du et al (30) and from Zhao et al (29) were obtained; providing both exact and censored serial intervals. Cases with non-positive serial interval were excluded from the Du et al dataset, due to estimation requirements and due to the characteristics of the serial interval. These data can be used to apply a Markov Chain MonteCarlo method to estimate the parameters of the lognormal distribution of the serial interval(39); a lognormal distribution was chosen since the original authors reported it provided the best fit. The lognormal distribution can be defined as the distribution of the variable *X* = *e^μ+σZ^*, where *Z* is a standard normal variable, and *μ* and *σ* are two real numbers, which are the mean and standard deviation of the logarithm of *X*. The implementation in the R package coarseDataTools(27) was used (Metropolis-Hastings algorithm, with 6000 iterations and thinning of 1000) and convergence checks were perfomed with the Gelman-Rubin diagnostic). We also used the Generation times from Ganyani et al obtained with data from clusters in Singapore and Tianjin. We took the mean and standard deviation estimates and their credible interval, and used them as the defining parameters of a triangular distribution (where the estimate is the mode and the credible interval the limits). From this we randomly sampled 5000 means and standard deviations of the serial interval, and calculated 5000 gamma distributions of the serial interval.

These distributions are, in turn, used to estimate *R_t_* using the R package EpiEstim(39)(24). Point estimates of *μ* and *σ* were also obtained using the R package coarseDataTools, and these are used to obtain *R^c^_t_* estimates using the R0 package, with the method described by Wallinga and Teunis(33).

This code and application are used to look for answers to several questions:

**Question A: What impact does data quality have on *R_t_* estimations?** Datasets can have many problems or issues. The effect of data quality on *R_t_* estimates (question A) has been analyzed in the Sup. material: The higher-quality COVID-19 incidence reports from the Spanish Public Health Surveillance National Network (RENAVE) have been compared to the initial and deprecated Ministry of Health/Government COVID-19 cumulative incidence reports (5.8); the impact of a spurious peak on the result has been examined (Sup. material, 5.4), and the impact of weekends and smoothing on daily incidences and the subsequent weekly and daily *R_t_* estimations has been evaluated by using official data from the Catalonia regional Government and Belgium Government (Sup. material, 5.10, 5.9).
**Questions B: What impact do preprocessing parameters have on *R_t_* estimations?** Transmission curves have been obtained with different settings. A rudimentary one-at-a-time sensitivity analysis is also performed (Sup Material, 5.6) using the average daily Gap between non overlapping credible *R_t_* intervals as an indicator. Sensible parameters are chosen and used in the rest of the study.
**Question C: What impact do the different serial intervals/generation times and estimation methods have?** Transmission curves for different *si/GT* distributions and estimation methods *[R_t_*, 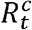) have been analyzed, sensitivity analysis has been performed with different distributions, also using the average daily Gap.
**Question D: What impact do wealth and development have on national transmission curves?** We have used the John Hopkins University dataset to obtain *R_t_* curves in order to analyze differences between developed and developing countries(40), including developed countries that consistently (Sweden) or temporally (United Kingdom) implemented mitigation instead of suppression policies, section 3.4. National Gross Domestic Product (GDP) per capita (purchasing power parity, constant 2017 international dollars) data from World Bank for 2018 (41) was obtained and used to colour the transmission curves so that economic impact can be visually evaluated. Hypothesizing that wealth might play a significant role in dataset quality, the relationship between GDP and dataset variability is also evaluated (Sup. material, 5.3).
**Question E: Do transmission curves follow a similar pattern on a sub-national/regional level?** We have also obtained regional *R_t_* curves by using the data provided by Spain Health Ministry(42); these data include positive PCRs and positive antibodies tests, but only the former was used since the dataset does not report the type of positive antibodies. While a positive IgG result can only be interpreted as a “present/past infection” (prevalence information), a positive IgM result could provide incidence information; however we assume a PCR will be performed in most positive IgM cases, following the standard protocols(43) so the impact of this limitation is expected to be minor. It should be noted that this problem was also present in many other datasets. Additionally, in the Sup. Material, we have compared the *R_t_* between different USA states and between different Perú regions (Sup. material: 5.11, 5.12). Regional GDP per capita (US dollars, current prices, current purchasing power parity) data from OECD for 2018 (if available, USA) or 2017 (Perú) (44) was used to color the transmission curves.
**Question F: Are transmission curves altered in suspected superspreding events?** The possible impact of the previously mentioned superspreading events has been analyzed in Italy, Spain, France and USA. Some of these events have been subjects of political debate; but the question examined here has an epidemiological nature: How did the transmission curves change around these events? Any other implication will not be discussed.
**Question G: Can transmission curves be used to detect outbreaks or to guide epidemiological measures in the reopening phase?** Finally, given the importance of Public Health surveillance in the reopening phase, we have analyzed the correlation between outbreaks and *R_t_* changes in the reopening phase, by using incidence data of several German districts and states (dataset compiled by J Gehrcke with information by the John Köch Institute), and the outbreak in Lleida, Catalonia (dataset by IDESCAT, Salut, Generalitat de Catalunya(45)), in the beginning of July (Sup. material, 5.13, 5.14). In these analysis several subnational divisions of varying entity and size are analyzed, in order to evaluate whether transmission peaks/valleys are reflected in outbreaks (or the lack of thereof). In 5.14 the analysis is complimented with a sensitivity analysis that includes several *si* or *GT* distributions, and an analysis of overall infectivity, as calculated by the EpiEstim package.

Many of these questions are mutually related: Data quality influences and determines *R_t_* estimations, the impact of superspreading events can also be seen on the regional level, COVID-19 data quality can be influenced by economic factors (Sup. material, 5.3), the smoothing might need to be increased in order to correct significant data quality issues, which might in turn reduce the sensitivity of the estimates.

The analysis of the temporal evolution of the *R_t_* (transmission curves) here is mostly performed qualitatively, considering the clinical and epidemiological implications: Whether or not the disease transmission is controlled (*R_t_* < 1), when does this happen and whether or not the disease transmission is increasing or decreasing.

## 3 Results

### 3.1 Question A: Data quality

In the 185 countries in the John Hopkins University COVID-19 dataset, 22 countries had any negative value (Antigua and Barbuda, Benin, Ecuador, Finland, France, Guyana, Honduras, Italy, Jordan, Lithuania, Madagascar, Mauritius, Nepal, New Zealand, Niger, Portugal, San Marino, Spain, Taiwan*, Uganda, Uruguay, Zimbabwe) and 23 countries had spurious positive peaks (Cameroon, Congo (Brazzaville), Congo (Kinshasa), Ecuador, Equatorial Guinea, Ethiopia, France, Gabon, Ghana, Guatemala, Israel, Kazakhstan, Kyrgyzstan, Montenegro, Nicaragua, Peru, Qatar, Singapore, Spain, Sudan, Sweden, Zambia, Zimbabwe). The peak in Spain has been analyzed in the Sup. Material, 5.4. In the European CDC dataset, 11 countries had any negative value (Benin, Ecuador, France, Gibraltar, Italy, Jordan, Lithuania, Portugal, San_Marino, Spain, Uganda). (It is therefore important to start any analysis with the visual exploration of the epidemic curve before and after smoothing.)

We have observed the proportion of positive tests has significantly changed over the course of weeks/months (Figure: 1, source: ECDC weekly data on COVID-19 testing), (Sup. Material, 5.2). In Castile and León, Spain, the daily proportion of positive tests in March nearly reached 60%; in Norway this proportion peaked at 17% in March; in USA it peaked at 23% in March and in Canada it peaked at 10% in April. Both the percentage of positives and the trend were different between USA (a developed country) and Perú (a developing one); in both cases the number of performed PCR tests gradually increased over from April to June 2020, providing more precise estimates of the real incidence as time passes. Other issues have been observed in the datasets: The reporting delay and diagnostic delay and associated factors (Sup. material, 5.1), the availability of different data sources for the same country (Sup. material, 5.8), the presence of large spurious peaks or frequent small peaks and valleys or the impact of the weekday bias.

**Figure 1:**
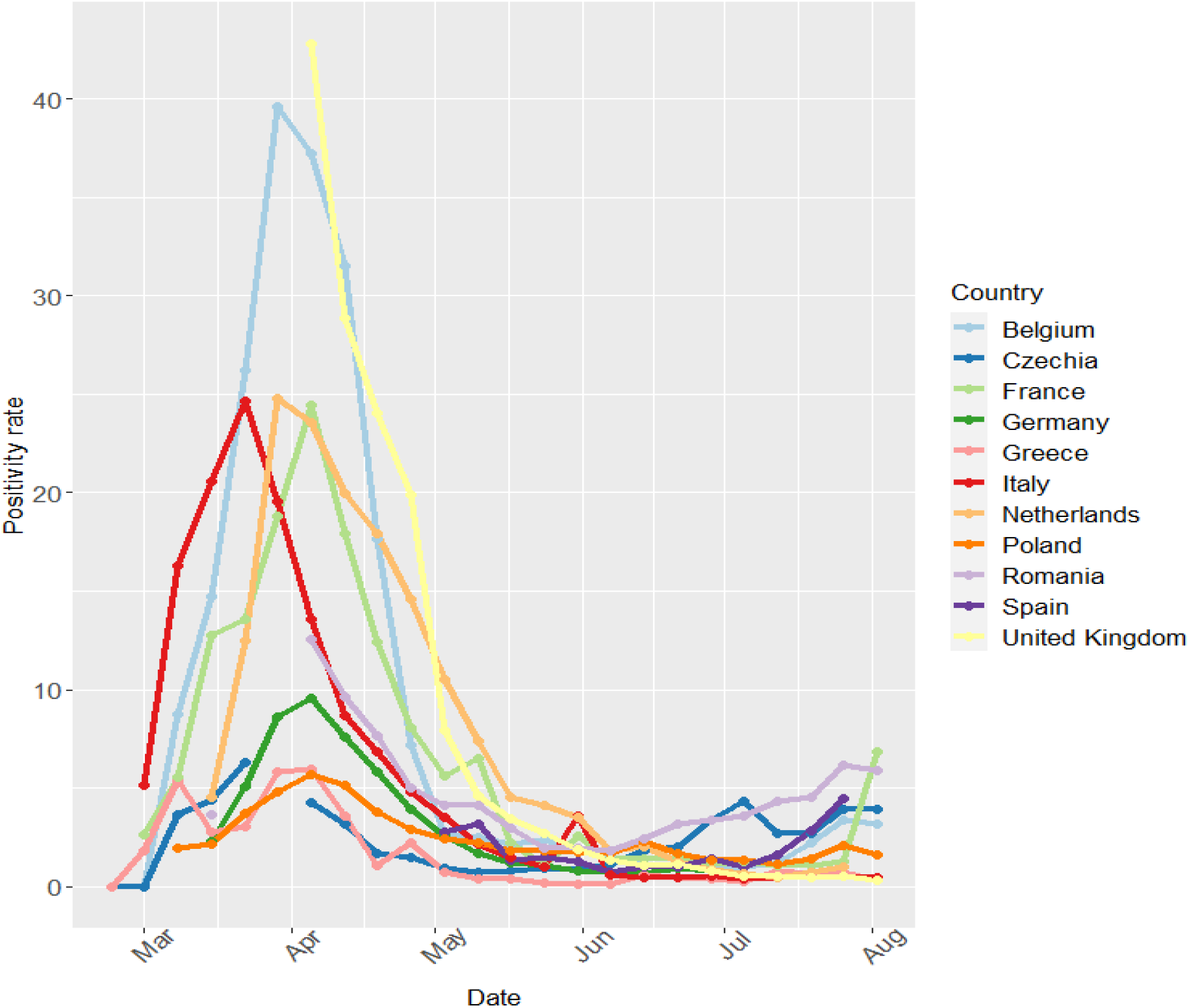
Positivity rate for large European countries.

### 3.2 Serial interval distributions

The point estimates for the serial interval and the reported estimates for the generation time are displayed in the table 1, with 95% credible intervals, along with percentiles. For the Log-normal distribution, *mu* (Mu) and *sigma* (Sigma) are reported; for the Gamma distribution, mean and standard deviation are reported. The MCMC serial interval distributions and bootstrap generation times are available in Figure 2.

**Table 1:** Serial Interval distributions as estimated via MCMC. Point estimates of generation time distributions (Ganyani)

| Source | Number of cases | Location | Distribution | Mu/<br>Mean | Sigma/<br>SD | Median | Mode | Percentile 5 | Percentile 95 |
| --- | --- | --- | --- | --- | --- | --- | --- | --- | --- |
| Du et al (si) | 361 | China | Log-normal | Mu= 1.458<br>(1.379, 1.543) | Sigma= 0.778<br>(0.725, 0.838) | 4.296 | 2.346 | 1.199 | 15.442 |
| Nishiura et al (si) | 28 | Articles and reports | Log-normal | Mu= 1.385<br>(1.167, 1.599) | Sigma= 0.558<br>(0.427, 0.752) | 3.996 | 2.926 | 1.594 | 9.982 |

| Source | Number of cases | Location | Distribution | Mu/<br>Mean | Sigma/<br>SD | Median | Mode | Perce-<br>tile 5 | Perce-<br>tile 95 |
| --- | --- | --- | --- | --- | --- | --- | --- | --- | --- |
| Zhao et al (si) | 21 | Hong Kong | Log-normal | Mu= 1.211<br>(0.852, 1.546) | Sigma= 0.722<br>(0.547, 1.038) | 3.358 | 1.993 | 1.019 | 10.968 |
| Ganyani et al (GT) | 54 | Singapore | Gamma | Mean= 5.2<br>(3.78, 6.78) | SD= 1.72<br>(0.91, 3.73) | 5.012 | 4.631 | 2.729 | 8.314 |
| Ganyani et al (GT) | 135 | Tianjin | Gamma | Mean= 3.95<br>(3.01, 4.91) | SD= 1.51<br>(0.74, 2.97) | 3.759 | 3.373 | 1.834 | 6.716 |

**Figure 2:**
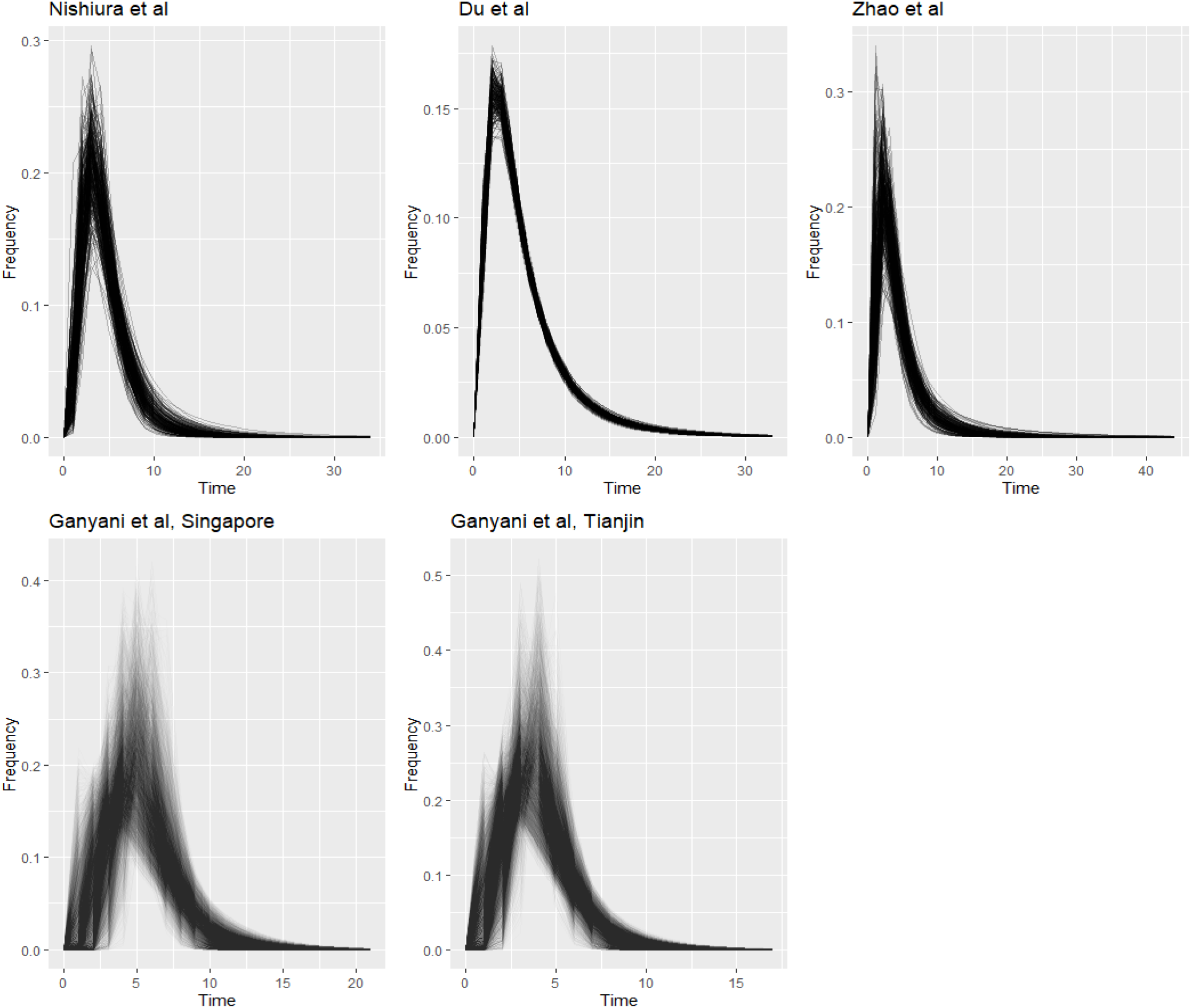
MCMC lognormal distributions of the of the serial interval and bootstrapping distributions of the generation time (Ganyani)

### 3.3 Questions B, C. Estimation parameters: Preprocessing, *GT/si* and estimation method

The different smoothing and preprocessing options did have a minor/moderate impact on the transmission curves (Sup. material, 5.5,5.6); briefly speaking, naïvely multiplying each daily incidence by a constant factor to correct the undetected proportion does not have any noticeable effect on the *R_t_* curves or create any gap between transmission curves, as previously described(24). Both Lowess and SMA smoothing showed good results and performed better than one-sided mean averaging. Smoothing midly influenced the final *R_t_* curves, but their characteristics and relevant trends (faster or slower decrease and epidemic control) are mostly similar. Smoothing can be specially important in datasets with spurious peaks (Sup. material, 5.4), and Lowess smoothing can provide the best results providing that the smoothing span in properly tuned. Smoothing can also correct the weekday-bias present in most datasets (Sup. material, 5.9, 5.10).

Figure 3 shows the influence of different Generation Time/serial interval distributions and estimation methods on the transmission curves, estimated with the JHU dataset (incidences between 20-Feb-2020 and 11-Aug-2020). Lowess smoothing with a span of 6 days was used since it better fit the data. (See sup. material 5.14 for another sensitivity analysis of several *si/GT* distributions).

**Figure 3:**
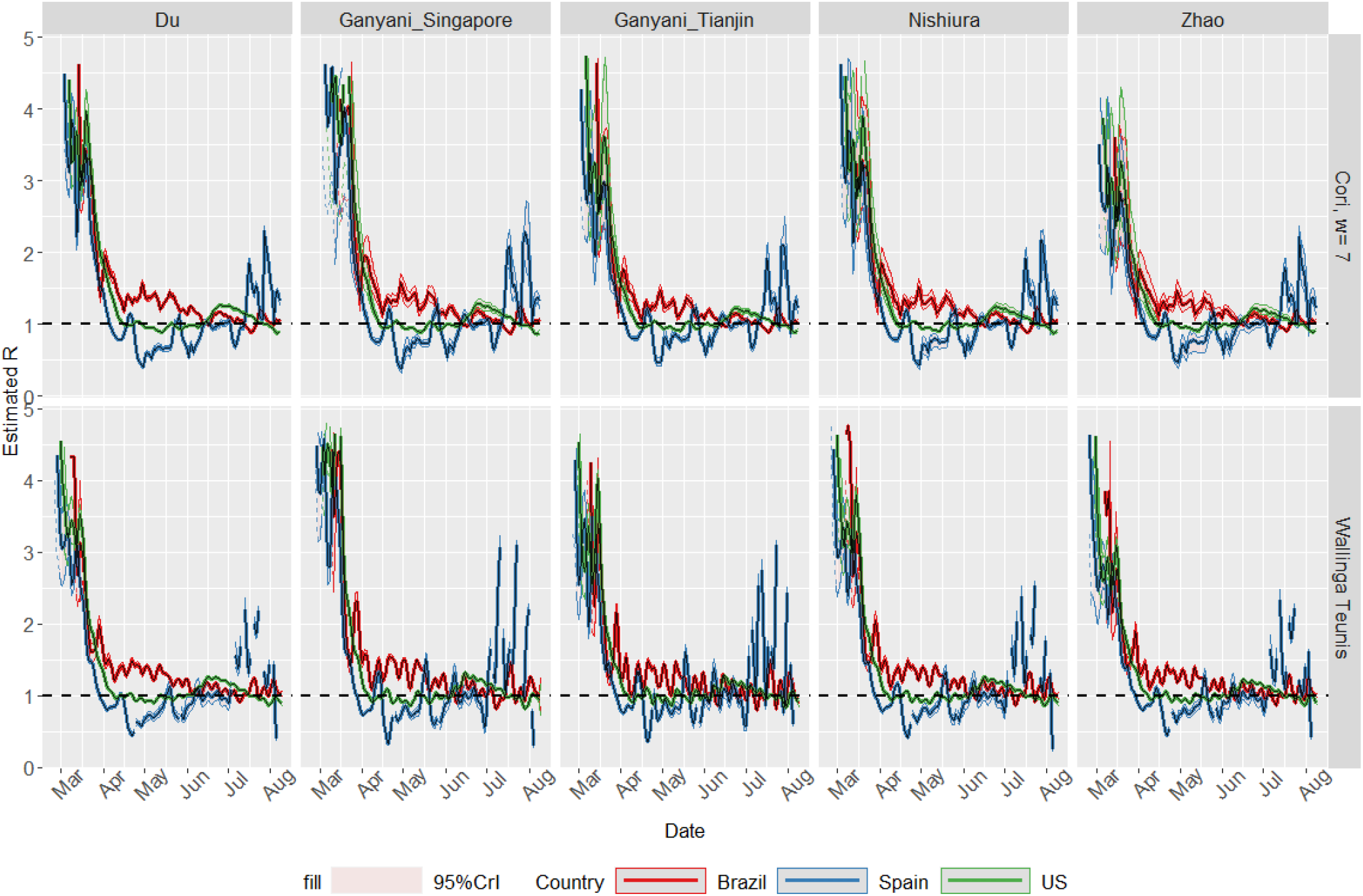
Transmission curves with different methods and distributions.

Some differences can be remarked: The credible intervals of the estimated *R_t_* were narrower when the serial interval distribution obtained from the larger Du et al dataset was used, and they were wider with the smaller Zhao et al serial interval dataset. A visible delay was observed when estimating the *R_t_* using the EpiEstim package (by Cori et al) compared with the R0 package (Wallinga Teunis method); by default, the EpiEstim package calculates the *R_t_* over a window period of seven days and the last date of the period is used to plot the estimate trend in the x-axis.

Apart from this small differences, the results were largely equivalent and both methods deliver similar conclusions. The EpiEstim package will be used in the rest of this study; and the serial interval samples obtained from the Du et al dataset via MCMC.

### 3.4 Question D. Results in several countries

Daily incidences have been obtained for several large countries (Brazil, Belgium, France, Germany, Iran, Italy, Japan, Sweden, United Kingdom, Russia, India, South Africa, Spain, Singapore, US, Colombia, Mexico, Chile, Pakistan, Bangladesh, Perú) which exhibited a large number of cases and/or deaths, for the period between 2020-02-20 and 2020-08-11 using the JHU COVID-19 dataset. Fig 4: epidemic curve of daily incidences after preprocessing; fig 5: transmission curve of *R_t_* by country. Preprocessing parameteris include assuming a time-constant proportion of 40% undetected cases (therefore, increasing the daily incidences by 66%), Lowess smoothing with 10 day span, epidemic start with 30 cases/3 days, *si* from the Nishiura et al dataset. World Bank data (GDP per capita, PPP, in constant 2017 international dollars(41)) from 2019 (2017 for Iran) is used to colour the curves.

**Figure 4:**
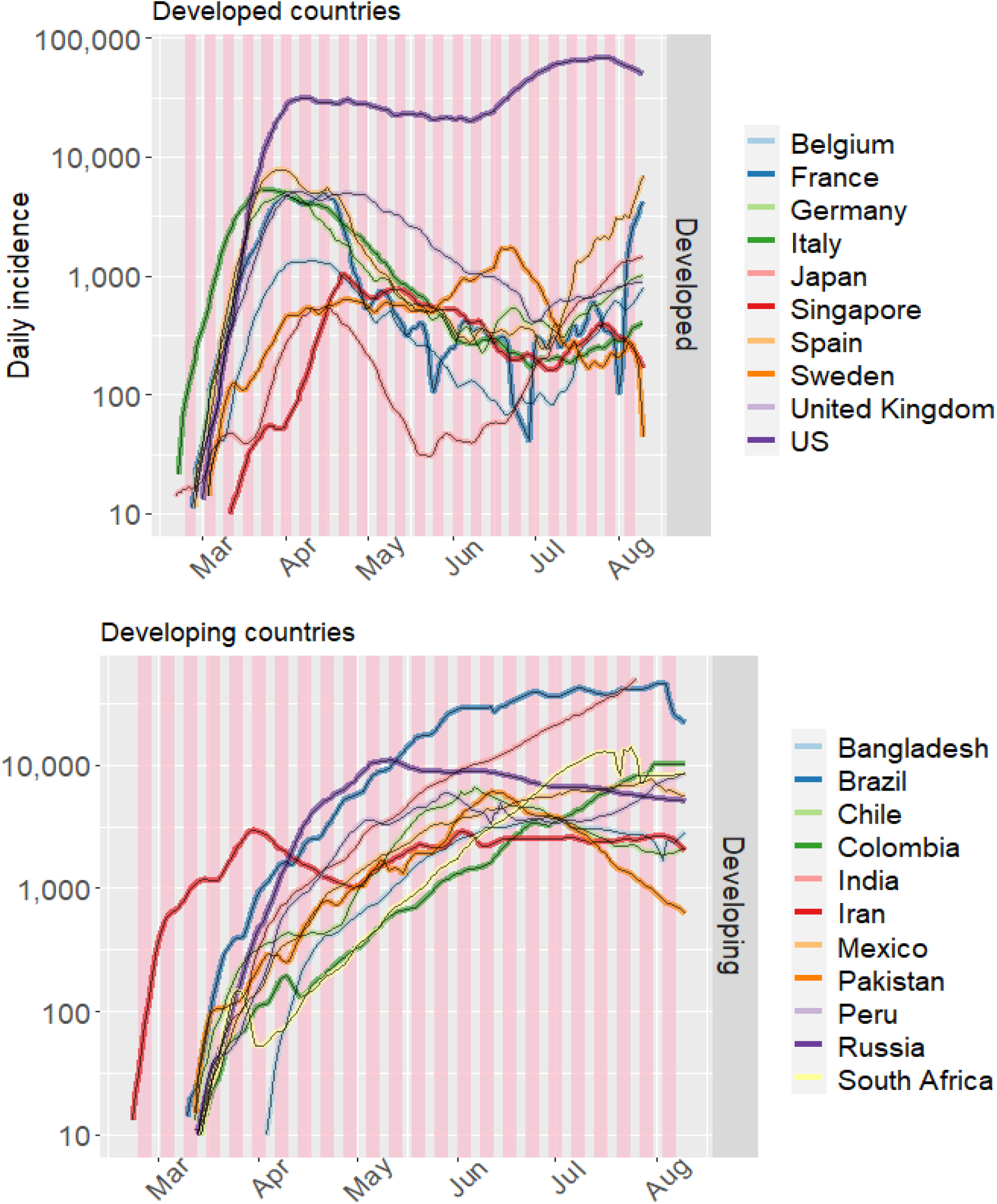
COVID-19 incidence in several countries.

**Figure 5:**
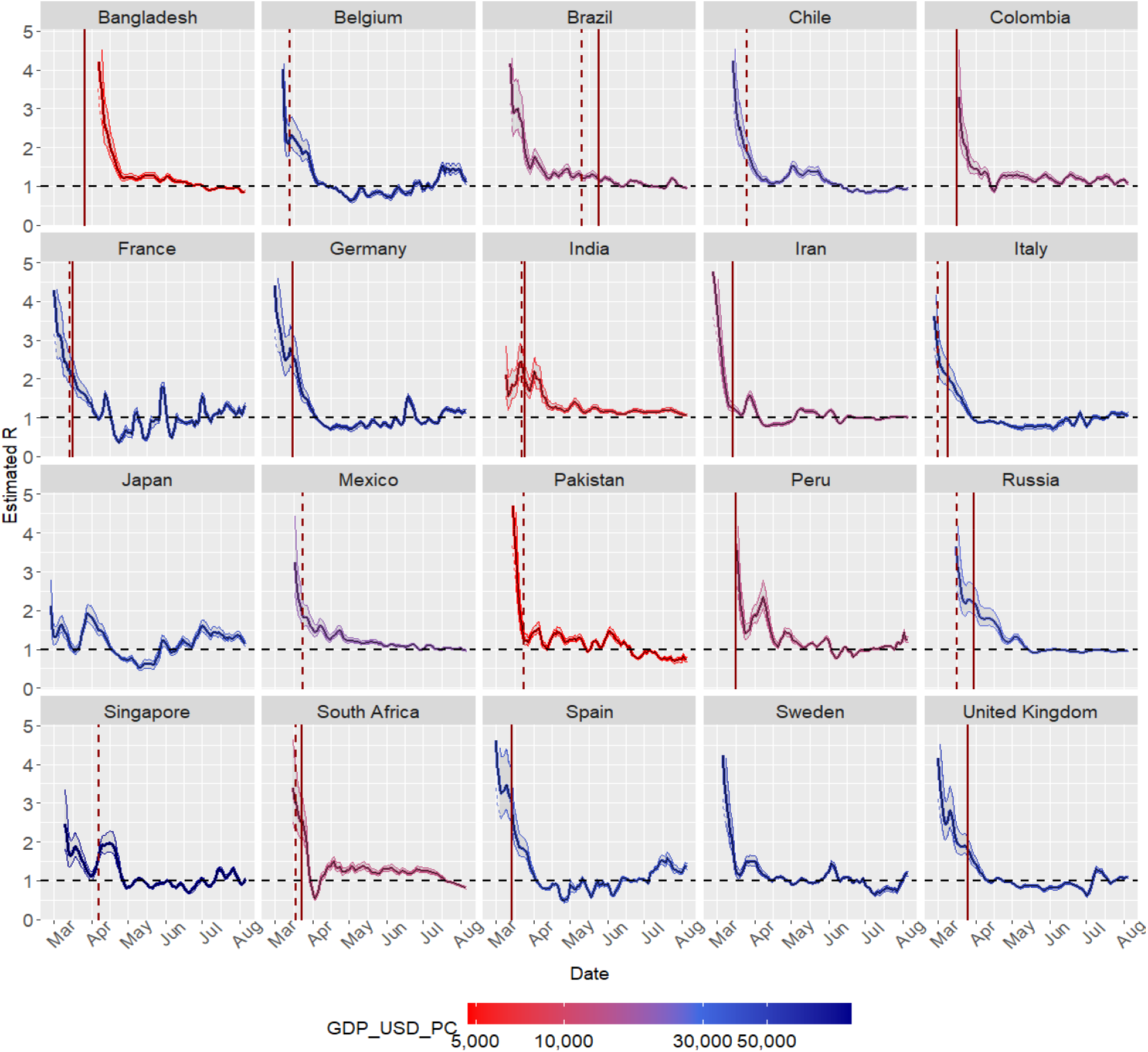
R_t_ estimates for several countries. Highlighted dates: solid line = full scale measures/lockdown; dashed line = smaller scale measures.

All countries experienced **exponential transmission** in March; earlier in some countries (Iran, Italy), and later in others (Russia, South Africa, India, Brazil), and faster in Italy, Iran, and Spain. Most developed countries managed to “bend the incidence curve” by the beggining of April, with decreasing daily incidences and *R_t_* < 1. In the UK epidemic control was reached somewhat later in April; Sweden and USA only tentatively reached this in April/May. Developing countries (Russia, India, Brazil, South Africa) had more difficulties reaching decreasing daily incidences, and their *R_t_s* stubbornly remained above the control level; Iran managed to control the epidemic at the end of March, but increasing incidences appeared again in May.

Italy and in particular Spain COVID-19 had an abrupt and exponential increase in daily incidences in late February / early March, respectively. Italy was the first European country to declare the State of Emergency (on the 31 January). On 6 March the Italian Anesthesiology and Intensive Care Association published their ‘Recommendations for treatment admission and suspension in exceptional conditions of disbalance between needs and available resources’ (46); this attests to the particular healthcare overload seen in this country around this time.

In Japan after an initial “valley” around the 15th March (*R_t_* ~ 1 and *I_t_* = 31), transmission increased, reaching Rt~1.5-1.8 at the beginning of April with *I_t_* = 563 on the 16th of April). *R_t_* started to decline around the 8th April (on 7 April a one-month state of emergency was decreed in the prefectures of Kanagawa, Saitama, Chiba, Osaka, Hyogo, and Fukuoka, and on 16 April the declaration was extended to the rest of the country for an indefinite period). However, no lockdown was implemented.

In late June and July many countries exhibit increases in their incidences.

Figure 6 displays the transmission curves of all world countries, using incidences and population values from the ECDC COVID-19 dataset(47), starting from 15-March-2020, coloured by their GDP per capita (with 2017 data from World Bank, Purchasing Power Parity in Constant 2017 dollars(41)) and using population as curve size. Preprocessing parameters include epidemic start after 50 cases in three days, lowess smoothing with 12 days as smoothing span. *R_t_* estimates have been obtained by using the Du et al serial interval MCMC estimations. Countries with less than one million inhabitants and *R_t_* estimates with 95% credible intervals wider than 1 have been excluded. This shows there is an important trend towards better epidemic control in countries with GDP per capita above 30000 USD dollars, especially in April/May (this difference is reduced in June, in the reopening phase). Many countries show later epidemic starts in April/May.

**Figure 6:**
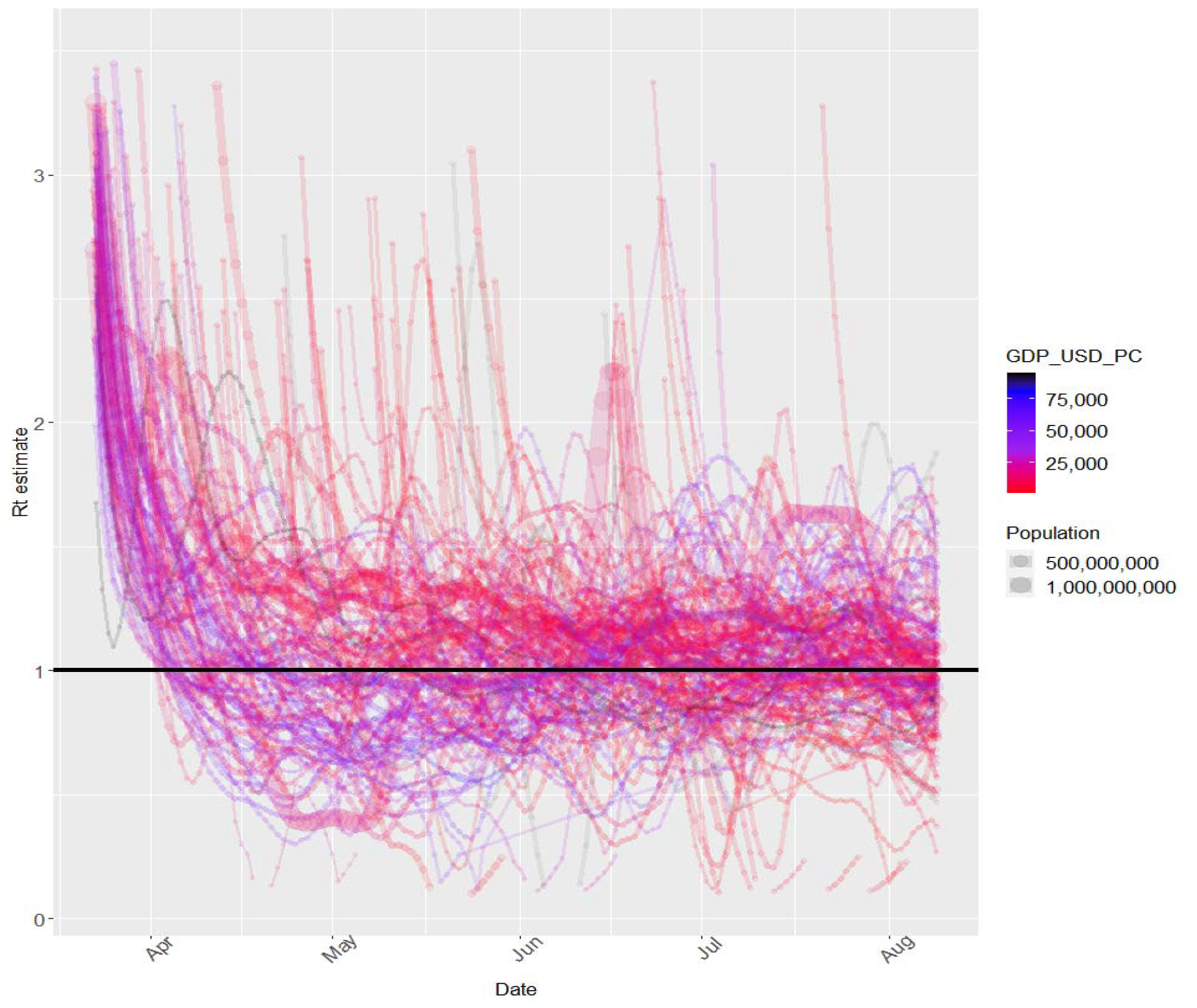
R_t_ estimates for large countries.

### 3.5 Question E. Spain Regional *R_t_* curves

The 9 Autonomous Communities with the most cases as of 2020-08-12 were selected and epidemic curves after preprocessing (Figure: 7) and transmission curves (Figure: 8) were obtained, by using the current official Health Ministry dataset (42), between 25-February-2020 and 2020-07-31. Preprocessing parameters include: centered SMA smoothing, 5 days smooth span, and +40% increase in daily incidences to account for undetected cases (due to the possible underestimation of cases in the initial period).

**Figure 7:**
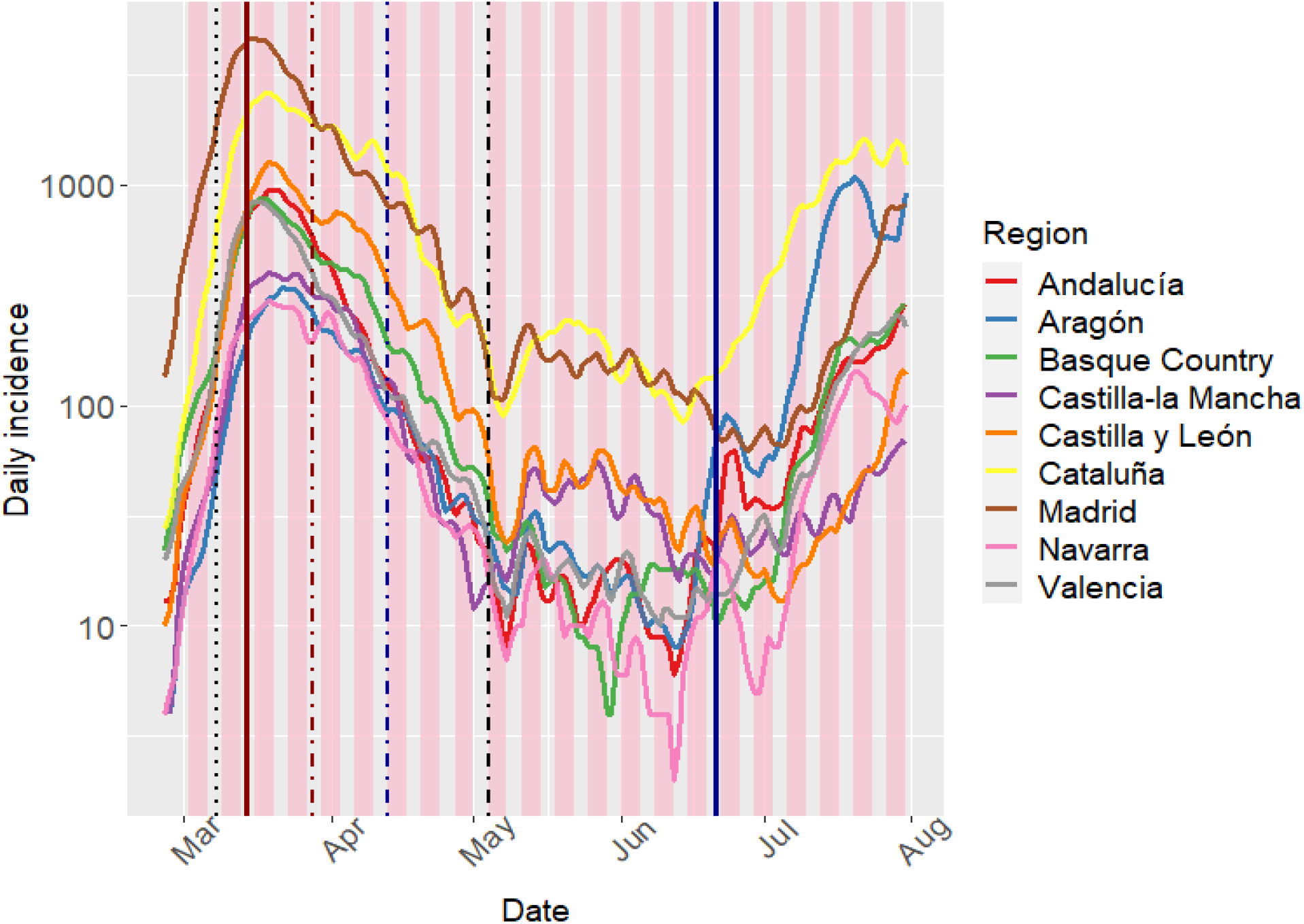
COVID-19 incidence in several Spanish Autonomous Communities. Highlighted dates: 2020-03-08 (8M), 2020-03-14 (State of Alarm), 2020-03-28 (Economic stop), 2020-04-13 (Partial economic restart), 2020-05-04 (Return to ‘normal’, phase 0), 2020-06-21 (End of the state of Alarm)

**Figure 8:**
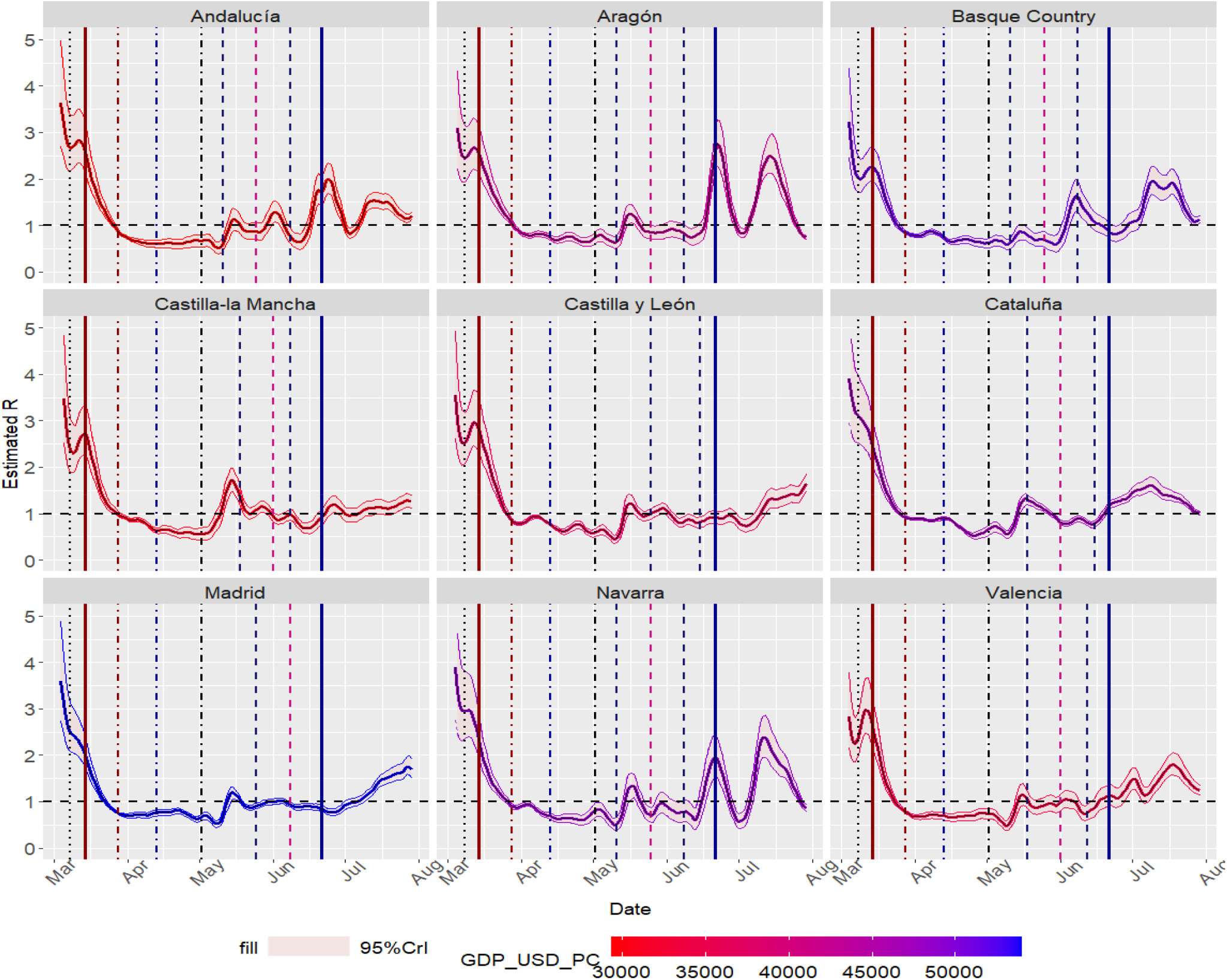
R_t_ estimates for several Spanish Autonomous Communities. Highlighted dates: 2020-03-08 (8M), 2020-03-14 (State of Alarm), 2020-03-28 (Economic stop), 2020-04-13 (Partial economic restart), 2020-05-04 (Return to ‘normal’, phase 0), phases 1 and 2,2020-06-21 (End of the state of Alarm)

Uncontrolled exponential dissemination took place early in March; particularly earlier in Madrid, the Basque Country and Catalonia, some of the most densely populated regions and important economic centers with the 1st, 2nd and 4th highest GDP per capita in the country. On the other hand, the epidemic peak took place later in Castilla-la Mancha. In the week after the 8th of March the *R_t_* started to decline, following the implementation of several contact-prevention measures. The State of Alarm declaration on 14 March was associated with a fast and sharp decline in the virus transmission reaching *R_t_<* 1 before the end of April, effectively heralding adequate epidemic control. Although the exponential increase did not occur at the same time or pace, the decrease was quite similar between regions.

The State of Alarm ended on 21st June 2020, after several weeks of gradual reopening with controlled transmission. After this several regions - not necessarily the wealthier and more densely-populated ones - exhibited a growth in the number of cases preceded by increased transmission (Aragón(48), Andalucía(49)…), eventually affecting Madrid and Catalonia.

Subnational/regional curves for USA (5.11), Perú (5.12), Belgium (5.10), Germany (5.13) and Catalonia (5.9, 5.14) are shown in the Sup. Material. Although they are most informative for large countries, useful information can also be drawn for regional transmission curves.

### 3.6 Question F. Superspreading events

Evaluating the COVID-19 spread in shorter periods requires a careful analysis of the underlying data and its variability and noise. Time series smoothing is moderately required. We have used data from the JHU-CSSE dataset(50) and obtained daily and weekly *R_t_* estimates. Application settings inclulde: Centered SMA smoothing, span of 5 days; +0% undetected cases and the serial interval from the Du et al dataset. Transmission curves: Figure 9.

**Figure 9:**
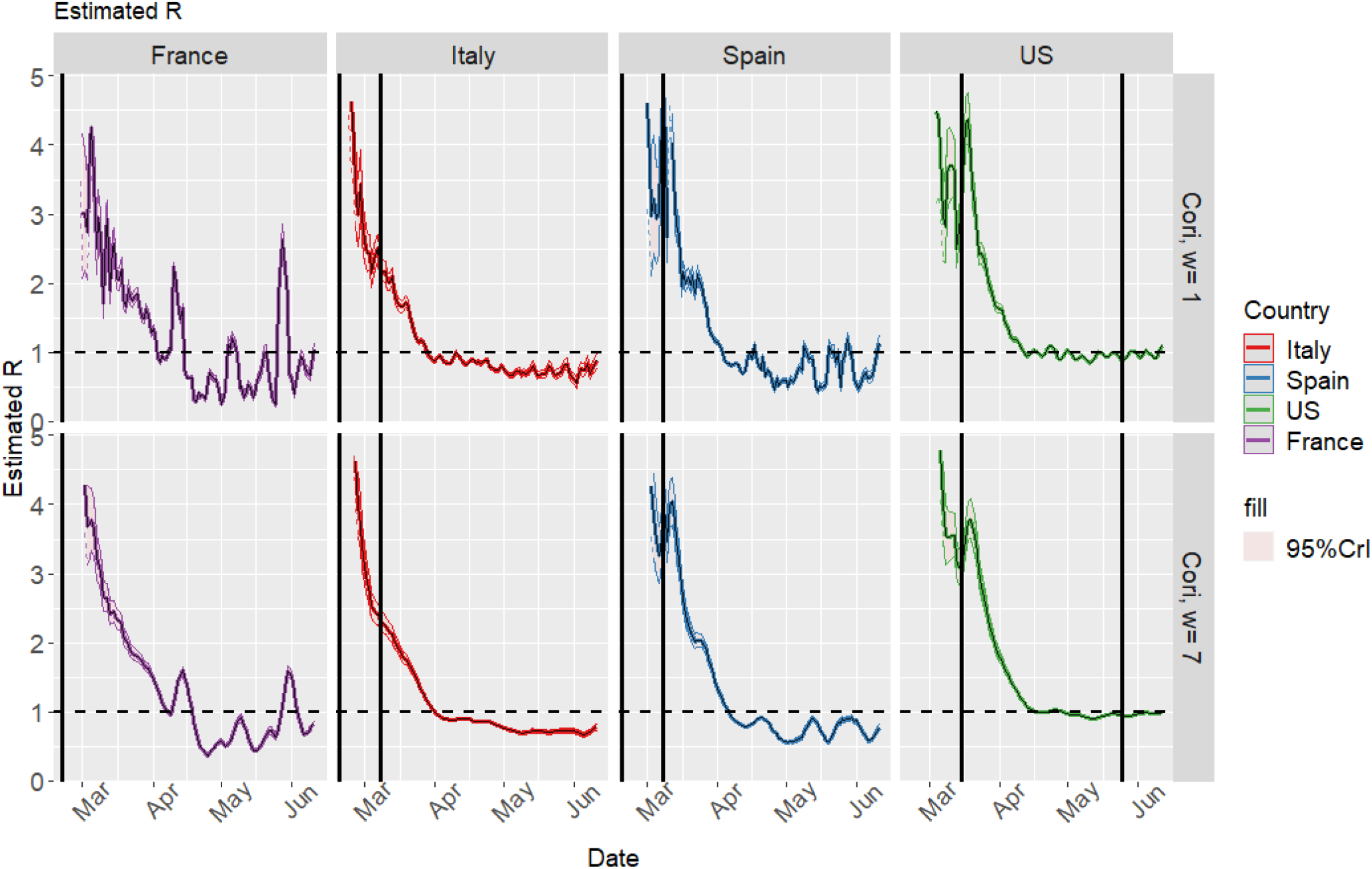
Transmission curves of countries with alleged superspreading events.

Both Italian and French events took place too early in the epidemic to draw any conclussion. In Spain the 8 March demonstrations and events took place several weeks later, in a period of exponential increase in the epidemic curve. A mild increase in *R_t_* is observed around that time, both in daily and weekly estimation windows, but the estimated *R_t_*s are too unstable and the credible intervals too wide. The three-day *R_t_* estimates from Spanish regions don’t exhibit any consistent and relevant increase in the *R_t_* around that date either (Sup. Material, 5.7), although a mild but inconsistent *R_t_* increase around that date is seen when using the initial less-quality cumulative incidence dataset (Sup. material, 5.8).

In the USA there was a transmission surge in late March, but this event did not take place at a well-defined date. *R_t_* estimates did not noticeably increase either in the riots after the death of George Floyd on the 25th of May. State-level estimates are provided in the Sup. material, 5.11; no consistent increase in *R_t_* is seen in early June: in some states a surge is seen (Arizona, Florida), but in most of them it is not, including Minnesotta (the state where this tragic event occurred), California, New York…

We have also noticed the apparition of peaks without any real-life event correlation; these peaks could be caused by noise in the time series or by reduced laboratory activity on the weekends (weekday bias, Sup. material, 5.9, 5.10).

## 4 Discussion

### 4.1 Dataset quality

Many factors can influence the process of obtaining transmission estimates. To start with, daily incidence data is required to perform the estimations, and data quality determines estimates quality. From the data analysis perspective, COVID-19 testing should be regarded a sampling process: Every day a sample of high pre-test probability cases are tested in order to obtain an estimate of daily incidence data; the (downwards) bias in this estimate will be the untested positive cases (with pre-test probability below testing cutoff). A variety of factors can increase the low pre-test probability untested cases or increase the cut-off required for testing: lack of readiness in the start of the pandemic, with some official protocols not requiring PCR confirmation for mild cases ((3),(43)), an increase in low COVID-19 probability cases (due to allergies, common cold, etc.), etc. The lack of adequate contact-tracing infrastructures can also hamper the pre-test probability assessments. We have observed the number of tests performed every day and the positivity rate has greatly varied over time and by location. Additionally smaller sampling sizes in this context can lead to higher variability in the incidence estimates. Therefore, any utilization of COVID-19 incidence/prevalence time series data for most purposes, including modeling or estimation, should be preceded by an assessment of this sampling process.

Increasing daily incidences by a constant proportion did not impact transmission curves, although it narrowed credible intervals. It is expected that a significant proportion of cases is not reported: In Spain 5.0% and 5.2% of the surveyed population were IgG+ against SARS-CoV-2 in the two phases of the National Seroprevalence Study(51), while only 0.42 and 0.52 of the participants in the study and 0.51 of the population on 03-Mar-2020 had had a positive PCR. Around 33% of infections were asymptomatic, in Spain and which is consistent with the 43% of asymptomatic participants in the population-screening study in Iceland(52), and somewhat higher than the 12% asymptomatic percentage of infections in the Colombia dataset(53),(Sup. Material, 5.1). Later epidemiological reports in August showed higher asymptomatic percentages in Spanish Regions (30-70%)(54). This percentage also indirectly informs us on the previously mentioned pre-test probability cut-off for testing: a small proportion of asymptomatic positives implies cases with low pre-test probability might not be adequately screened. Some authorities report suspicious cases; in the case of Catalonia these cases increased some weeks after the epidemic peak, when the epidemic was controlled and opening measures were being implemented; therefore they don’t seem a good surrogate for non-reported cases (Sup. Material, 5.9).

The heterogeneity in data reports/definitions is another important issue that impairs comparisons between countries and regions. In the COVID-19 Tracking Project, each state in the USA is graded according to the quality and completeness of the data they provided. Some countries, states or regions limit their reports to the number of positive cases, and do not inform on the number of tests performed, or do not report PCR and antibodies tests separately, or do not separate IgM and IgG antibodies tests. International standardization or consensus recommendations regarding COVID-19 reporting and data definitions could improve the data quality and facilitate comparisons.

In addition to this, negative incidence values or unexpected peaks can be observed in the temporal series of some countries or Spanish regions(55), which do not correspond to new cases but to delayed reports, data correction, validation and consolidation processes. When individualized epidemiological case-reports-derived statistics were available in May/June in Spain(56)(57), with the incidence defined by the symptoms-onset date, unnecessary delays were eliminated and data accuracy remarkably increased. Some dataset issues (variability, peaks) can be partially solved by smoothing.

Finally time intervals exist between infection and symptoms appearance (incubation period), between symptoms appearance and diagnosis (diagnosis delay) and between diagnosis and report(report delay). Delays have been analyzed in the Colombia dataset (Sup. material, 5.1); a small association with month of the year has been found. In Spain these delays vary between regions (diagnosis delay in August 2020: median: 3 days, IQR: 1-5 (54)), and the imputed diagnostic delay was greater in the initial stages of the (diagnosis delay in May 2020: median: 5 days, IQR: 3-11(58)). The onset-date should be as close as possible to the infection date, and comparisons should be performed with uniformly defined datasets. In Spain, the Community of Madrid, unlike other regions, defines the daily incidence *I_t_* for day t as the number of positives whose sample was taken on day t or the result was obtained(55); this is better suited for temporal series analysis since this eliminates the sample processing and report delay. Most delays could be solved in a similar way to asymptomatic transmission(24): If the distribution of the delay between infection and report is known, this could be used to back-calculate the incidence of infections from the incidence of reports, but this yields oversmoothed estimates (32). Other reproductive number estimation methodologies went further than us by using linedata and bootstrap methods to obtain a delay distribution, which is used to change the incidence data(59). In Spain, case-reports-derived statistics (RENAVE) attempt to report the symptoms onset date, and thus are less prone to delay bias (Sup. Material, 5.8). (This is a partial solution because the symptoms onset is still not known for asymptomatic patients).

### 4.2 Non pharmacological interventions to control the spread

The experience in Wuhan highlights the importance of Public Health Interventions to reduce the spread: The epidemic was resulting in congested hospitals and high rates of transmission^ ~ 4-5); but control (*β*_t_<1) was achieved through a combination of medical and preventive regulations: increases in healthcare capabilities and supplies, adequate patients isolation, transmission routes blocking (public transportation suspension, closure of public places, cordon sanitaire of Wuhan City, etc.) and the prevention of new infections (compulsory face-masks, universal stay-at-home policies and symptom surveys, etc)(60). The *R_t_* values markedly declined around the 24 of January, when measures including city lockdown were implemented.

Both in developed countries and developing countries, the *R_t_* reflects the impact of Public Health Interventions to reduce the spread of the coronavirus. The suppression strategy has proven to be more effective, when widespread community transmission is taking place; however developed countries with mitigation or less-stringent strategies fared better than developing countries. Most countries implemented lockdowns: including non-essential economy halting (Spain, Italy), subnational lockdowns (USA, Chile, Brazil, Russia) or national lockdowns (India, Spain, France, United Kingdom). In the case of Singapore, the third most densely populated country in the world, and South Korea, a successful containment was achieved in February and March 2020. The hallmarks of this strategy are travel restrictions, early detection and early quarantine of positive cases and their contacts, widespread testing, reduction in number of contacts and no or minor restrictions to economic activity(61). However in April in Singapore a new increase in *R_t_* heralded increase a new wave with particular increase in daily incidences, several orders of magnitude greater than the previous wave. In Europe, different non-pharmaceutical interventions have been applied by the authorities; in compartmental models the stay-at-home enforcement yielded the strongest reductions in normalized numbers of contacts (62).

In the case of Spain, initial **containment** efforts (such as the quarantine of an hospital) were successful(63), but soon it was apparent that the uncontrolled and exponential community transmission was taking place, as evidenced in both the epidemic curves and the *R_t_* curves. In the 9th of March **reinforced containment** measures were initiated (halting in-person education, workplace contacts avoiding, etc.) for areas with significant community transmission (which were the region of Madrid and the cities of Vitoria and Labastida)(64); transmission started to decrease as education institutions and public places were gradually closed in the different regions. The Government decreed a State of Alarm on the 14th of March, switching to a **supression strategy** imposing a nation-wide lockdown involving commercial places closures, stay-at-home regulations and a ban on internal travels. Thus a consistently decreasing *R_t_* and the effective control of the disease it hallmarks was reached(6), and Spain managed to successfully ‘bend the curve’. The reproductive ratio has been shown to be highly dependent on mobility(4)(65).

On the other hand, some countries used a **mitigation-oriented strategy:** The Swedish Government declared that they seeked to “reduce the pace of the virus’s spread” in order “to ‘flatten the curve’ so that large numbers of people do not become ill at the same time” (66). The epidemic curve of Sweden was indeed mostly flattened -but not bent-between April and July 2020. It could be argued that whatever policy was followed by developed countries, they are more likely to achieve it than developing countries.

### 4.3 Patterns and analysis of transmission curves: Developed and developing countries and regions

Transmission curves show that both developed and developing countries “attempt” to reduce the transmission; developed countries manage to control it and “bend the curve” faster and better, except for those that did not attempt suppression or attempted suppression later, but they are prone to outbreaks in July, also noted by (67)). Both in developed and developing countries, richer and more densely-populated regions had earlier starts of the exponential propagation and often exhibit higher incidences but better transmission control.

Developing countries can face multiple challenges when it comes to estimating transmission parameters, (lack of testing facilities, lack of compliance with the “social distance” mandates) (68); however we have noticed many developing countries have shown strikingly exhaustive, resourceful and complete datasets(53)(69). In developing countries the rise in the exponential phase of the epidemic was notably slower, but they had more difficulties than developed countries to control the epidemic. At the time of writing this, the *R_t_* of India was and the *R_t_* of Brazil was. In the case of India initial containment seemed successful, but was soon followed by widespread community dissemination that required more stringent preventive measures(70): social distancing, public places closures, whole country lockdown on the 24th of March…

After epidemic control, the apparition of outbreaks is effectively associated with *R_t_* surges, as seen in the cases of Iran, Spanish regions, German regions, Singapore, etc. These outbreaks were not exclusively associated with more developed and wealthier regions, even though these regions were the first ones to suffer the COVID-19. We have shown both incidence curves and transmission curves can be used to detect these outbreaks, but transmission curves should be combined with incidence data.

### 4.4 Superspreading events

Superspreading events range between **smaller collective events** involving hundreds-thousands of participants and being amenable to epidemiological investigation that can properly demonstrate epidemiological relationship (Diamond Princess cruise, Annual Assembly of the Christian Open Church in France); and **larger collective events** involving tens of thousands of participants which cannot be easily tracked (Atalanta vs Valencia FC match, 8M events in Spain). Some of these took place in February or in early March, before lockdown measures were implemented, or despite preventive measures. The European CDC recommended mass gatherings cancellation in exceptional cases for scenarios 1 or 2 in their 2-march Rapid Risk Assessment(5). We couldn’t find any conclusive evidence for any impact on transmission curves: some took place too early in the epidemic, and/or showed questionable and inconstant transmission surges with wide credible intervals, considering the methodological nuances (confirmation bias, weekday bias, correlation vs causation…).

### 4.5 Study limitations and advantages

In addition to the data limitations, using the serial interval as a surrogate for the generation time has a number of potential issues (32), and the serial interval distributions could be biased (overestimation/underestimation) due to several factors(30). For example, the serial interval might be contracted during the epidemic peak due to early quarantine of symptomatic infected individuals. We have not observed marked epidemiologically-relevant differences between the results obtained with any of the five distributions tested; so we believe it is often acceptable in practical terms to use the serial interval as a surrogate for the generation time.

Compartmental models can be used to estimate many infectious disease parameters, including the reproduction numbers(71)(62)(3). These models often divide estimate the initial reproduction number *R_0_* and the control reproduction number *R_c_* (after control measures have been initiated). Stochastic SIR models can also be used to obtain reproduction numbers in the initial phase without preventive measures (*R*_0_) and reproduction numbers after preventive measures have been implemented (*R_c_*, **control reproduction numbers**) which showed some variability between different regions but incontrovertibly demonstrated the efficacy of preventive measures(72). In the UK before the lockdown period the *R_0_* was estimated as 6.94 (6.52-7.39, credible interval) with a SEIR model(3). Initial *R_0_* (February) estimates in Wuhan were slightly lower (average of estimates: 3.28, median: 2.79) than our initial estimated *R_t_*s (73). A compartmental model in Belgium also yielded *R*_0_ = 3.334 *R_c_* = 0.713 (71). The effective reproduction numbers in the control phase we obtained are somewhat similar to the *R_c_*, but the initial reproductive numbers we obtained are somewhat higher than some estimates; these initial *R_t_* seem somewhat unstable in the time-since-infection models used here, particularly if no clear distinction is made between local and imported cases when performing the estimations. Both types of models rely on their own assumptions, and can be considered as complementary. In time-since infection models no assumptions are made regarding the shape of the transmission curve.

One of the advantages of this study and the application is the possibility of interactively performing sensitivity analysis and exploring different smoothing and preprocessing parameters (or lack of them), we have shown smoothing had some influence in the *R_t_* curves. However, the main information conveyed by the *R_t_* estimates - namely, epidemic control - did barely change, although in some cases untreated spurious peaks can lead to wrong outbreak conclusions.

There is not a single definite way to obtain estimates of disease transmission to guide public policies; we have explored some of the most common ones and different parameters and found they have a small impact in transmission curves. The information obtained with this tool, which emphasizes parameters tuning, can be combined with many other modeling tools available on the Internet.

Additionally, one of the most relevant implications for the reopening phase are the outbreaks. Robust epidemiological monitoring and rapid detection of increased transmission are essential to avoid resurgences (67). We have observed outbreaks are amenable to detection with transmission curves, providing that quality datasets are available. Increased testing capacity is observed in many countries in June and July 2020, thus reducing noise and variability and improving incidence data accuracy.

Finally, one of the objectives was to bring epidemic information closer to the general population. We hypothesize that population compliance with preventive measures (social distancing and mobility restrictions) might improve when adequate information is provided. In this application the users can interactively tailor several steps (data source, preprocessing/smoothing, estimation window, serial interval, etc) to obtain slightly different *R_t_* estimations but nevertheless consistent with a fundamental idea: epidemic control requires lowering transmission, and the effect of uncontrolled transmission on incidence is exponential.

## 5 Supplementary material

### 5.1 Question A. Dataset quality: Diagnostic delays

In Spain the National Epidemiological Vigilance Network (RENAVE) initially in May described the interval between symptoms onset and diagnosis (median = 6 days, interquantile range (IQR) = 3-11 days, cases = 157885) and the interval between symptoms onset and case report (median=6 days, IQR = 3-11 days, cases = 169797 at 21-05-2020)(74). In the official Chilean dataset(69), the mean difference between symptom onset and case notification, as calculated from aggregate data, is 3.7.

The Colombia official positive cases dataset(53) has been used to assess the diagnostic delays (defined as the difference between symptoms onset and diagnosis) and the reporting delay (defined as the difference between diagnosis and web report).

11.66% of the reported cases as of 2020-08-12 were asymptomatic. The diagnostic delay for symptomatic cases has the following characteristics:

- Mean= 11.153
- SD= 11.153
- Percentiles 2.5, 25, 50, 75, 97.5 = 2, 7,11,15, 23
- Distribution = As shown in figure 10

**Figure 10:**
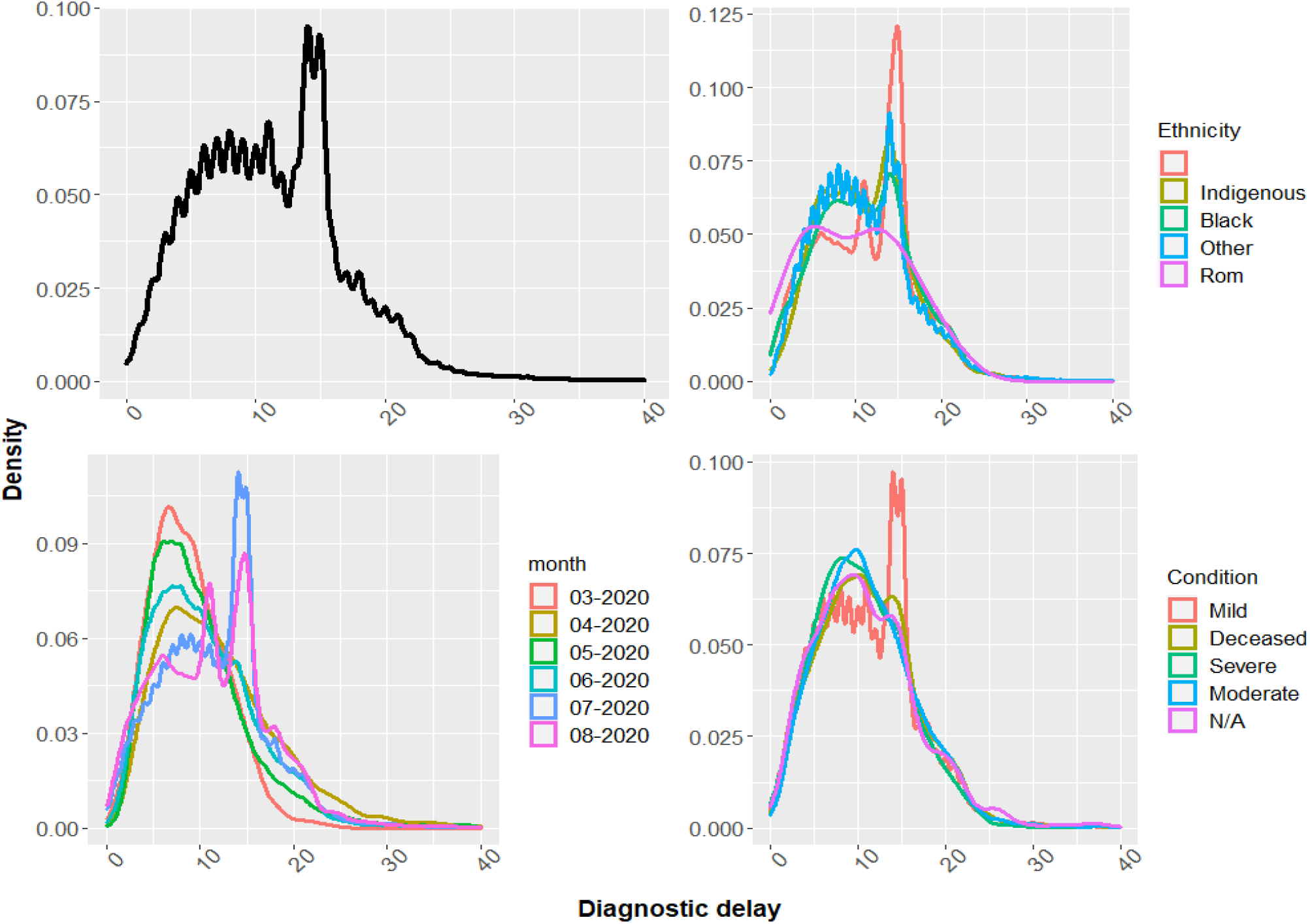
Distribution of the diagnostic delay.

An anomalous peak of mild cases with 13-14 days of diagnostic-delay is observed in July, probably corresponding to data imputation. A linear model has been created with log-transformed diagnostic delay as an dependent variable, and condition, ethnicity and month of diagnosis as predictors (Table: 3). This model was chosen even though log-transformed models are less interpretable. With an adjusted *R^2^* of 0.0052398, the model performance is poor, but many predictors are significant (table 3).

**Table 2:**
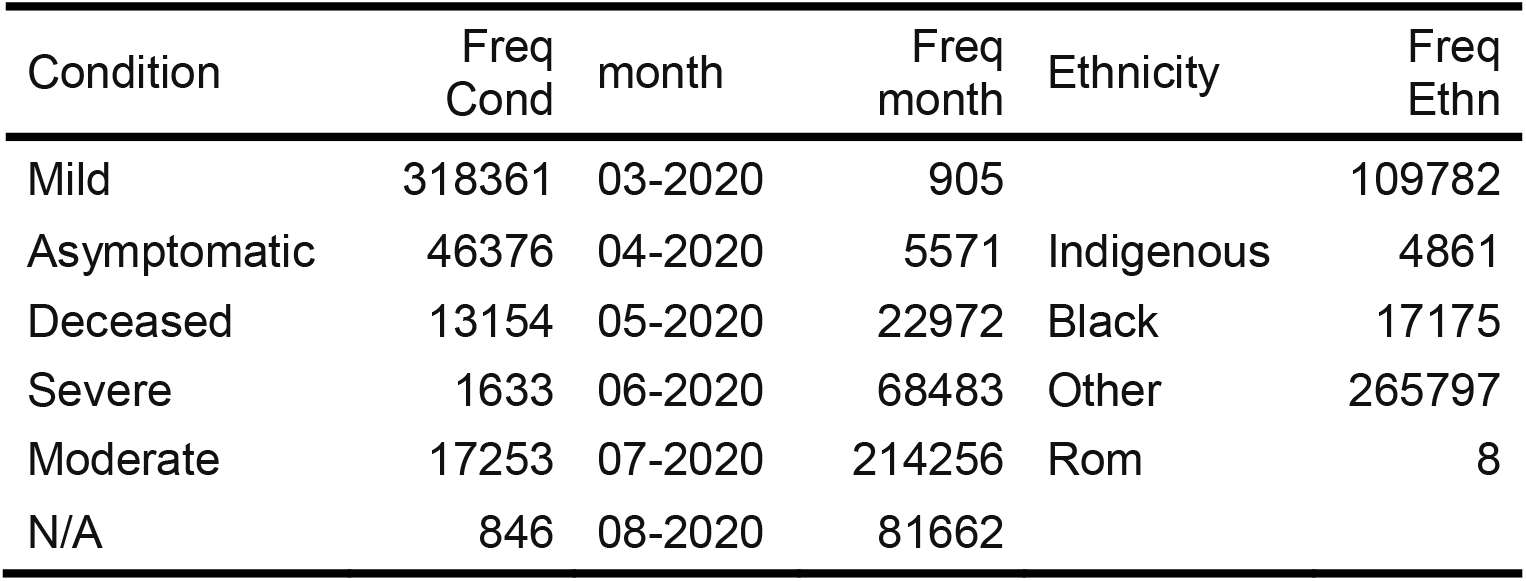
Colombia COVID-19 summary

| Condition | Freq Cond | month | Freq month | Ethnicity | Freq Ethn |
| --- | --- | --- | --- | --- | --- |
| Mild | 318361 | 03-2020 | 905 |  | 109782 |
| Asymptomatic | 46376 | 04-2020 | 5571 | Indigenous | 4861 |
| Deceased | 13154 | 05-2020 | 22972 | Black | 17175 |
| Severe | 1633 | 06-2020 | 68483 | Other | 265797 |
| Moderate | 17253 | 07-2020 | 214256 | Rom | 8 |
| N/A | 846 | 08-2020 | 81662 |  |  |

**Table 3:**
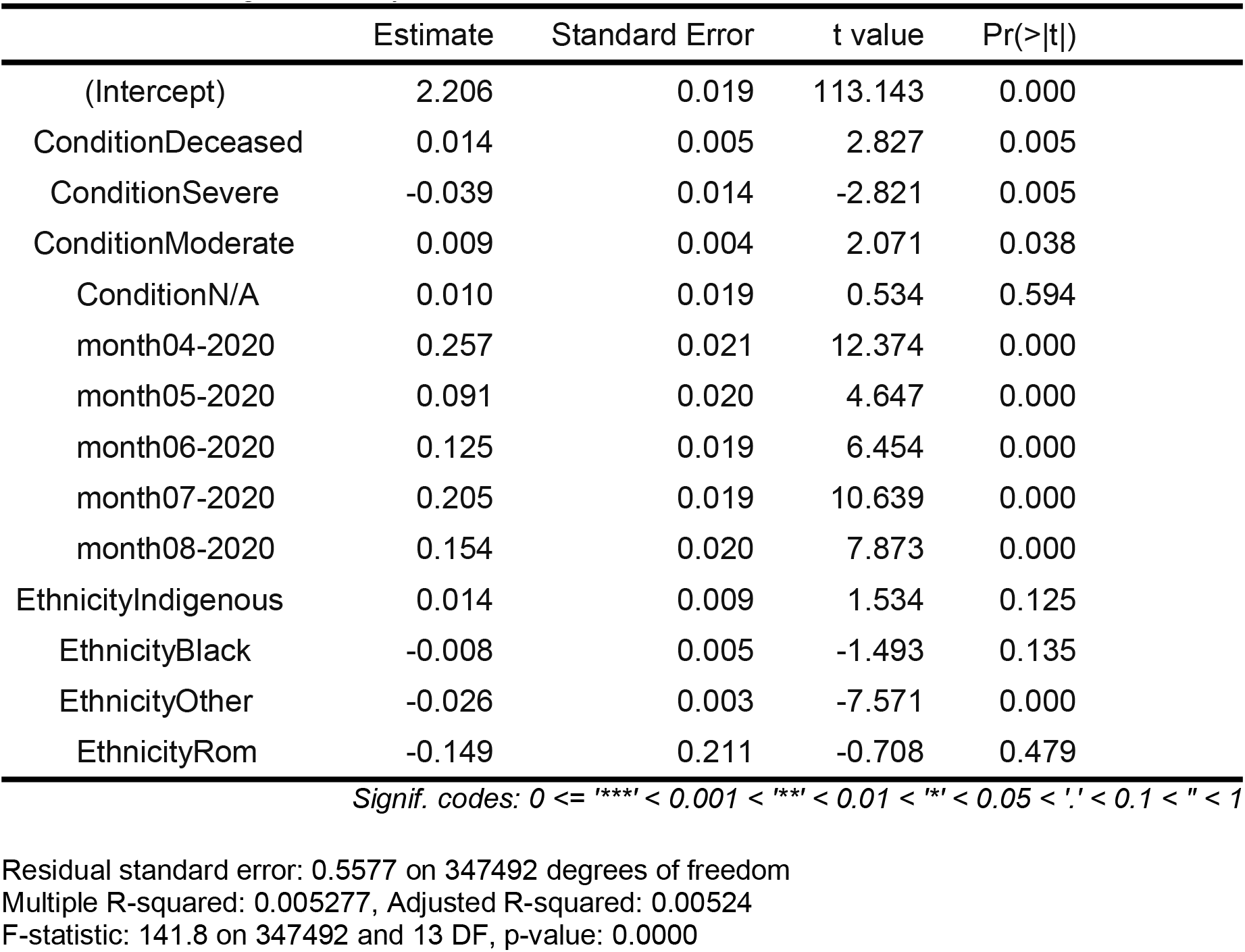
Linear model of diagnostic delay

Mild cases of March with non-specified ethnicity were used as a reference. Diagnostic delay was greater in April, May, June, July and August when compared to March, and greater in patients who died, and greater in black or indigenous patients, but smaller in patients of other ethnicities. However, these differences were mostly small in this dataset.

The following table summarizes the distribution of the reporting delay.

**Table 4:**

| Reporting delay (days) | Number of reports |
| --- | --- |
| <0 | 303 |
| 0 | 181133 |
| 1 to 5 | 187847 |
| >5 | 24566 |

Differences in diagnostic and reporting infrastructure might explain these results.

### 5.2 Question A. Number of tests and rate of positivity

The rate of positivity, the number of samples tested positive for COVID-19 in a given location and period, is defined by the WHO as one of the epidemiological criteria of epidemic control: It is met when less than 5% of samples test positive for COVID-19 for at least the previous 2 weeks, but this can only be interpreted when there is thorough surveillance and testing of positive cases(16); this criteria has been translated into national protocols(14) and monitoring tools(75). Herein we use the number of tests and the rate of positivity to assess temporal fluctuations/differences/constraints in the testing process, so the proportion of undetected cases could also vary over time, and the real incidence might particularly be underestimated when the rate of positivity is high.

Five datasets have been used to analyze the temporal distribution of testing efforts and, when available, the daily rate of positivity: The Perú data provided by the Peruvian Ministry of Health (MINSAL) and compiled by JM Castagnetto (76), the Canadian Government Data on COVID-19 in Canada (77), the USA data from the John Hopkins COVID-19 Testing Insights Initiative (CC BY-NC-4.0 license), as reported by USA states(75), dataset containing the number of COVID-19 tests performed in the region of Cantabria, Spain, between 2020-03-01 and 2020-06-24 and the dataset containing the number of COVID-19 tests performed in the region of Castilla y León, Spain(78), between NA and 18-May-2020 (CC BY-3.0 license).

The number of COVID-19 PCR and antibodies tests performed each day are graphically reported, and the proportion of positive results is also reported, when available. A smoothing LOESS trendline is added due to considerable variability between days and due to a “weekday-bias” effect particularly noticeable in antibodies tests. Figure 11 shows the COVID-19 PCR tests performed in the USA, figure 12 shows the information for Perú, figure 13 shows the tests performed and the positive rate in Canada; with a possible nuance in the former dataset: some USA states might not have clarified what kind of tests they are reporting (PCR or antibodies).

**Figure 11:**
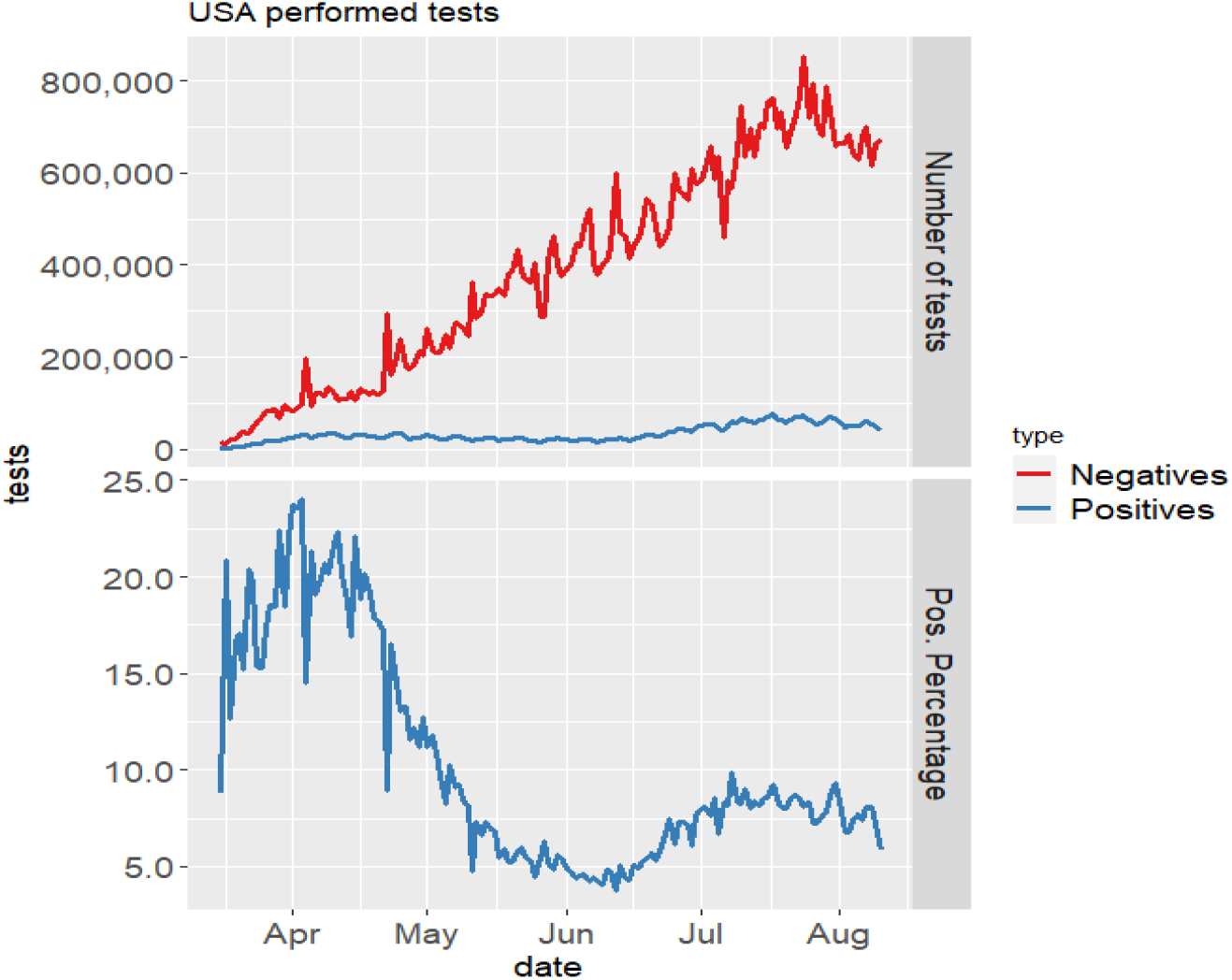
PCR tests performed in the USA.

**Figure 12:**
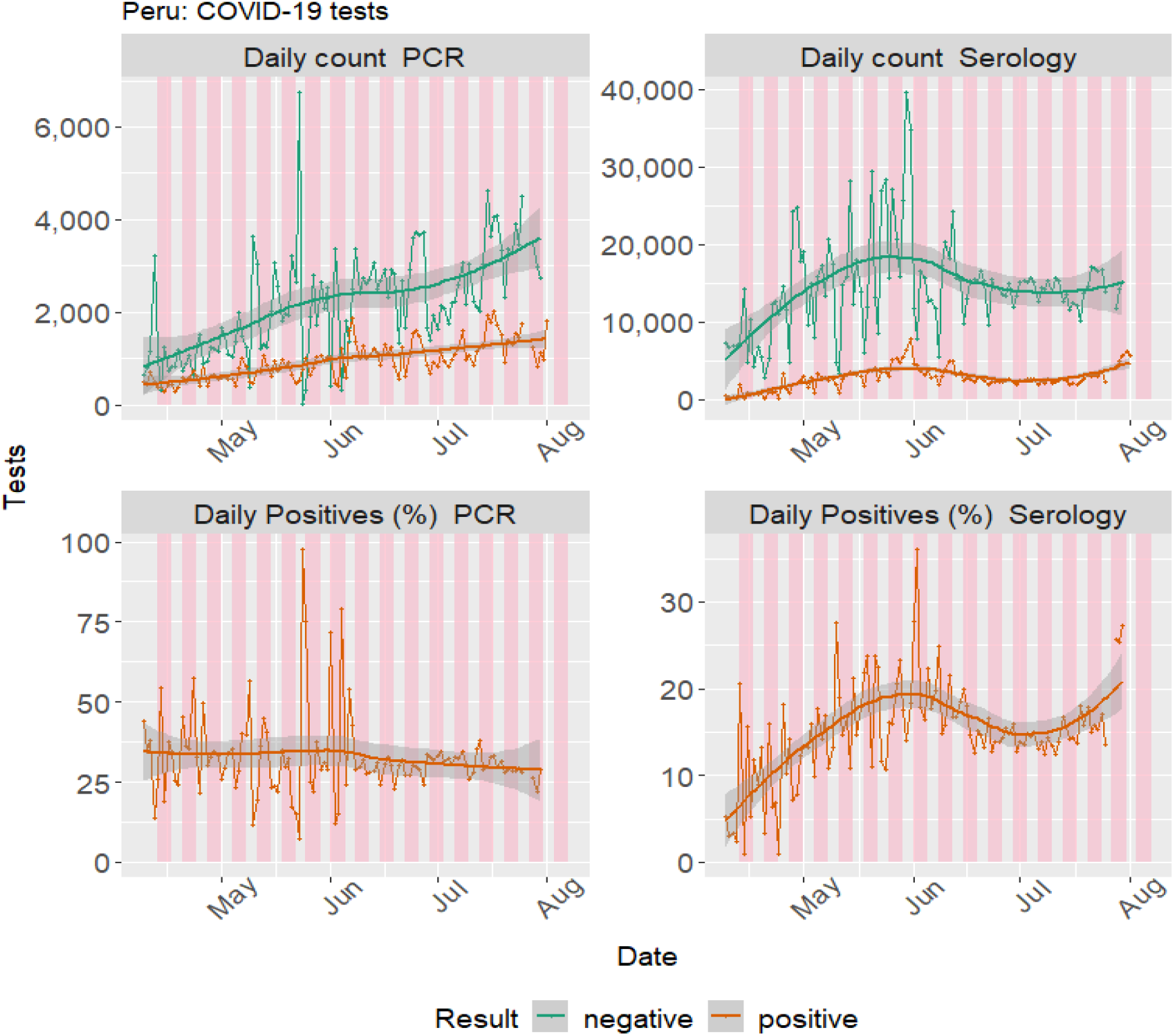
COVID-19 tests performed in Peru.

**Figure 13:**
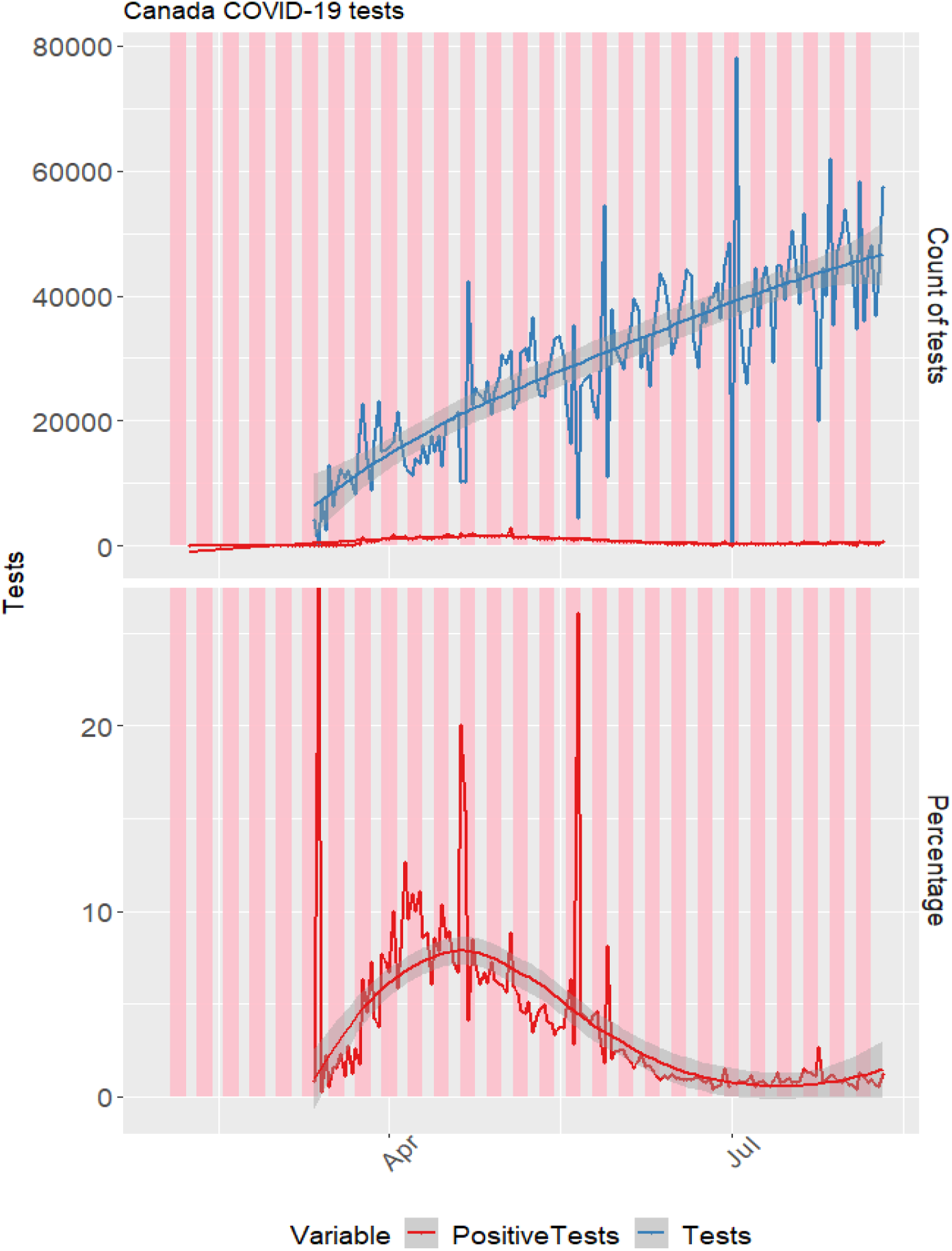
COVID-19 tests performed in Canada.

Both the USA and Canada have shown a steady and almost linear increase in testing capacity over time, reaching 600000 and 40000 daily tests respectively at the end of June. Perú also linearly increased their testing capacity over time. In both countries the daily percentages of positive tests have evolved: in the USA the rate of positivity approached 20-25% in April, but then a marked decline was observed and a plateau of ~5% positive tests was reached in May, but in late June, the rate of positives started to increase mildly. In Canada the peak of positives was reached in mid April, around 9%. In Perú the percentage of positive PCRs remained uniform around 30% between April and June, and no information is available for earlier months.

Figure 14 shows the numbers of PCR and antibodies tests performed in this Northern Spanish region. Only the number of performed tests is reported.

**Figure 14:**
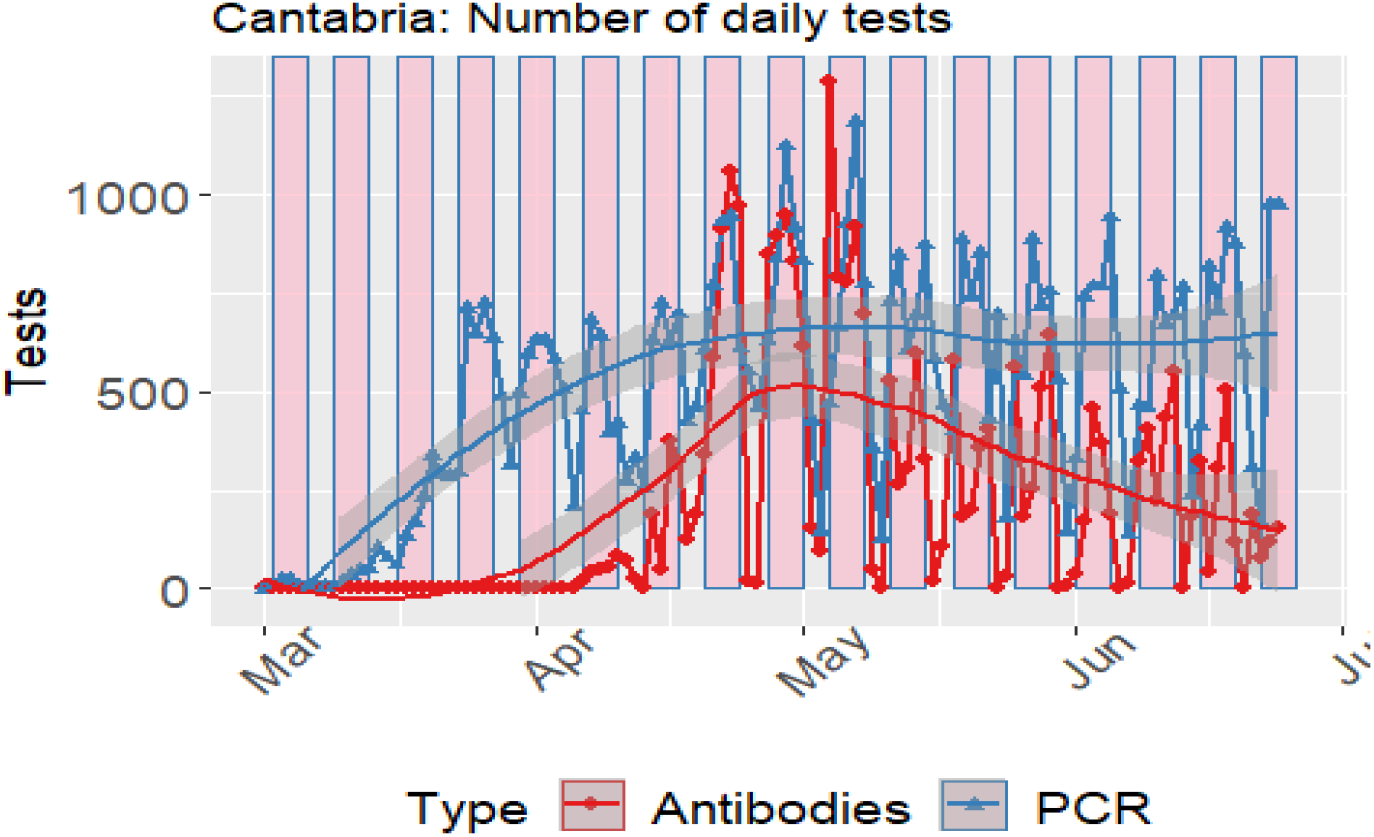
COVID-19 tests performed in Cantabria, Spain.

Figure 15 shows the numbers of PCR and antibodies daily tests performed in Castilla y León, and the percentage of positive results for the whole region; figure 16 show the province-level information.

**Figure 15:**
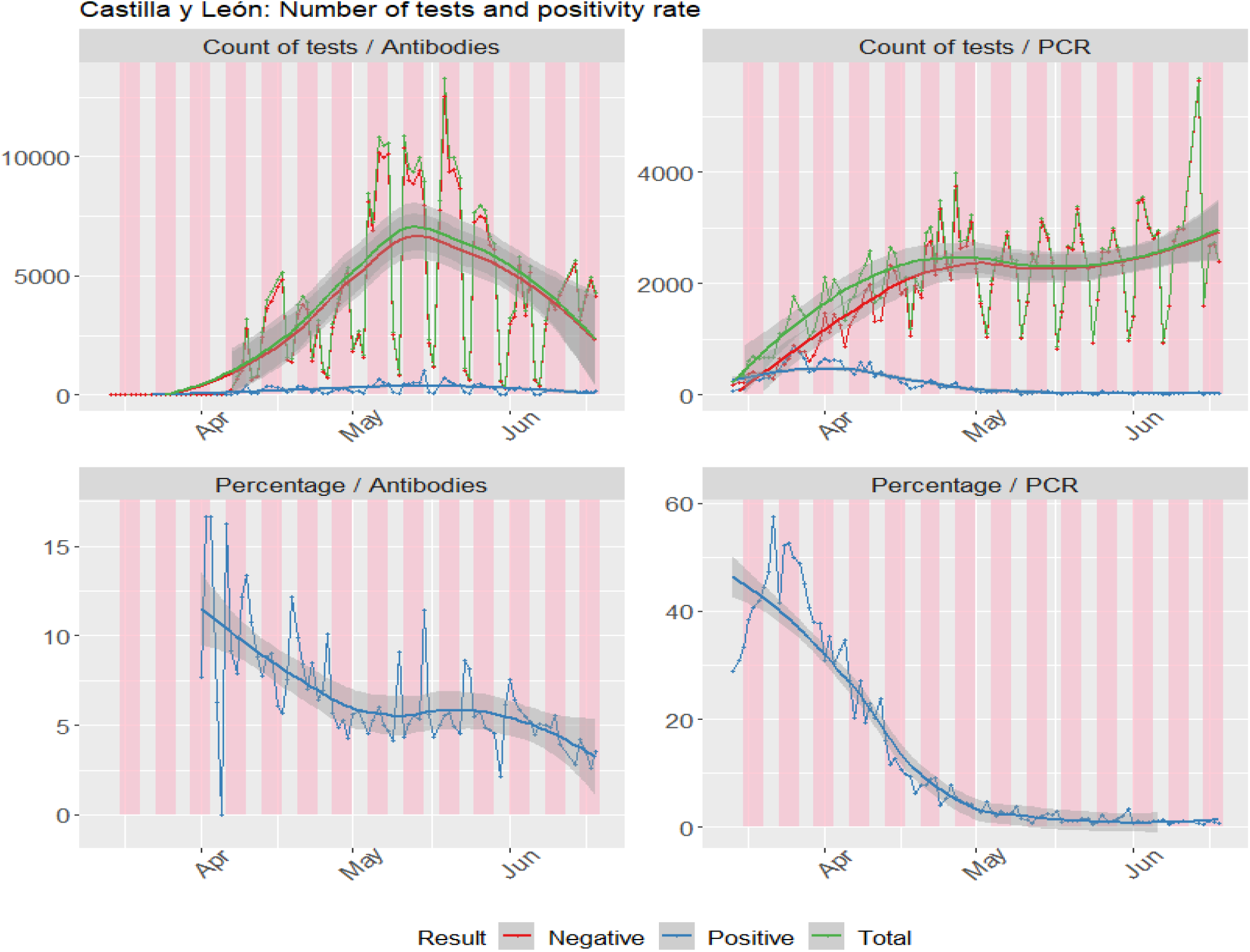
COVID-19 tests performed in Castilla y León, Spain.

**Figure 16:**
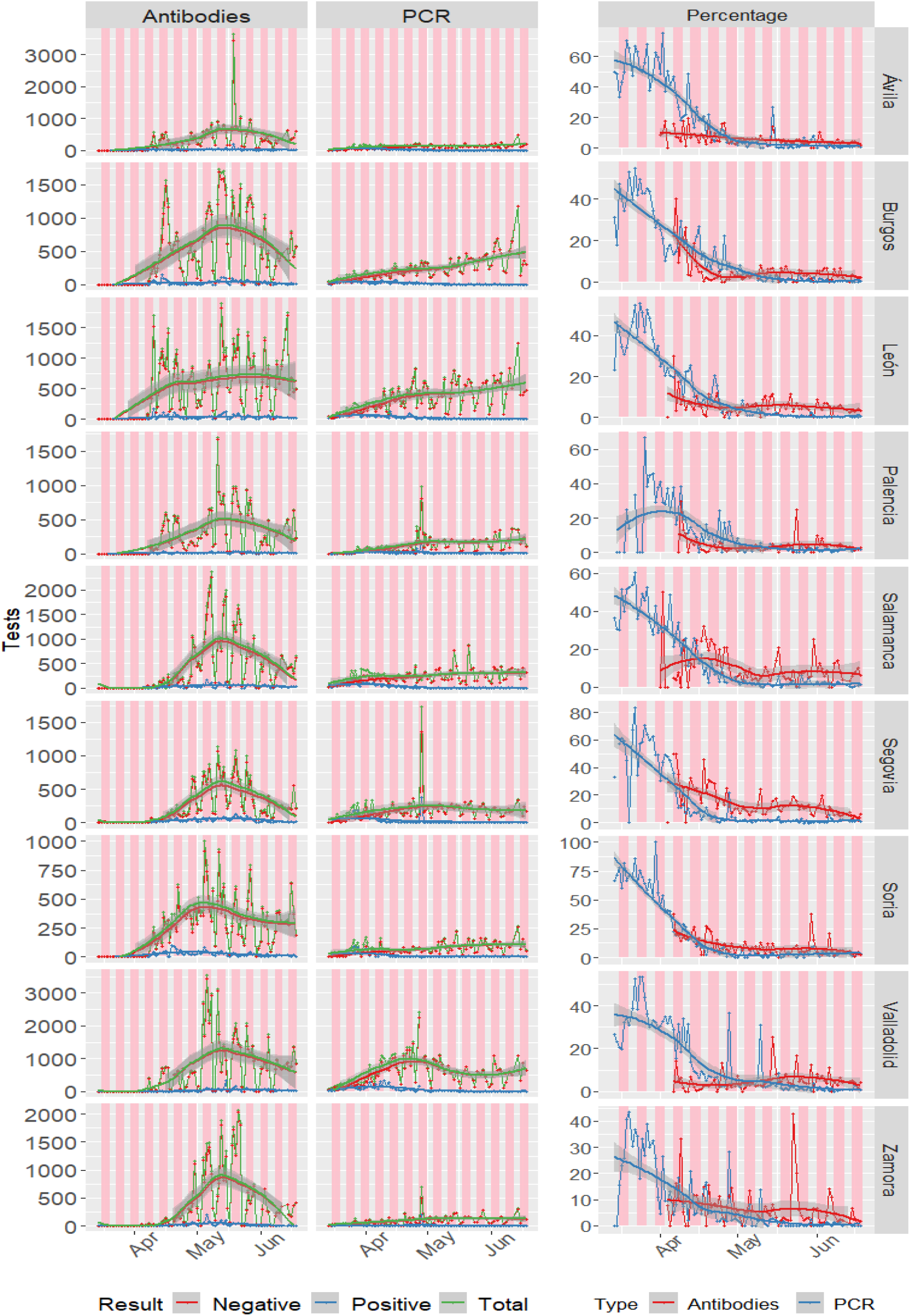
COV1D-19 tests performed in Castillay León provinces, Spain.

In both regions of Spain a considerable **weekday-bias** is observed. The PCR testing capacity of Cantabria increased in March and peaked in May. In Castilla y León the rate of positive tests was higher than 40% in March, which could have implications for underestimation of the daily incidence. Both regions showed a stable number of PCR tests performed in May/June, coinciding with epidemic control and reopening phases.

### 5.3 Questions A, D. Data variability

We define variability here as the mean absolute error of the original incidences vs the Lowess-transformed incidences, using a smoothing span of 7 observations. (This absolute error definition might overestimate the effect of days with large incidences; however the relative counterparts might have the opposite effect). The European CDC dataset (47) has been used to calculate the variability per country and the total sum of cases by country, using data between 25-Feb-2020 and 11-Aug-2020; this information has been combined with the OECD economic information (GPD per capita, in US dollars, PPP, 2018). Figure 17 shows the relationship between variability and total number of cases; countries with less than 100 cases are excluded.

**Figure 17:**
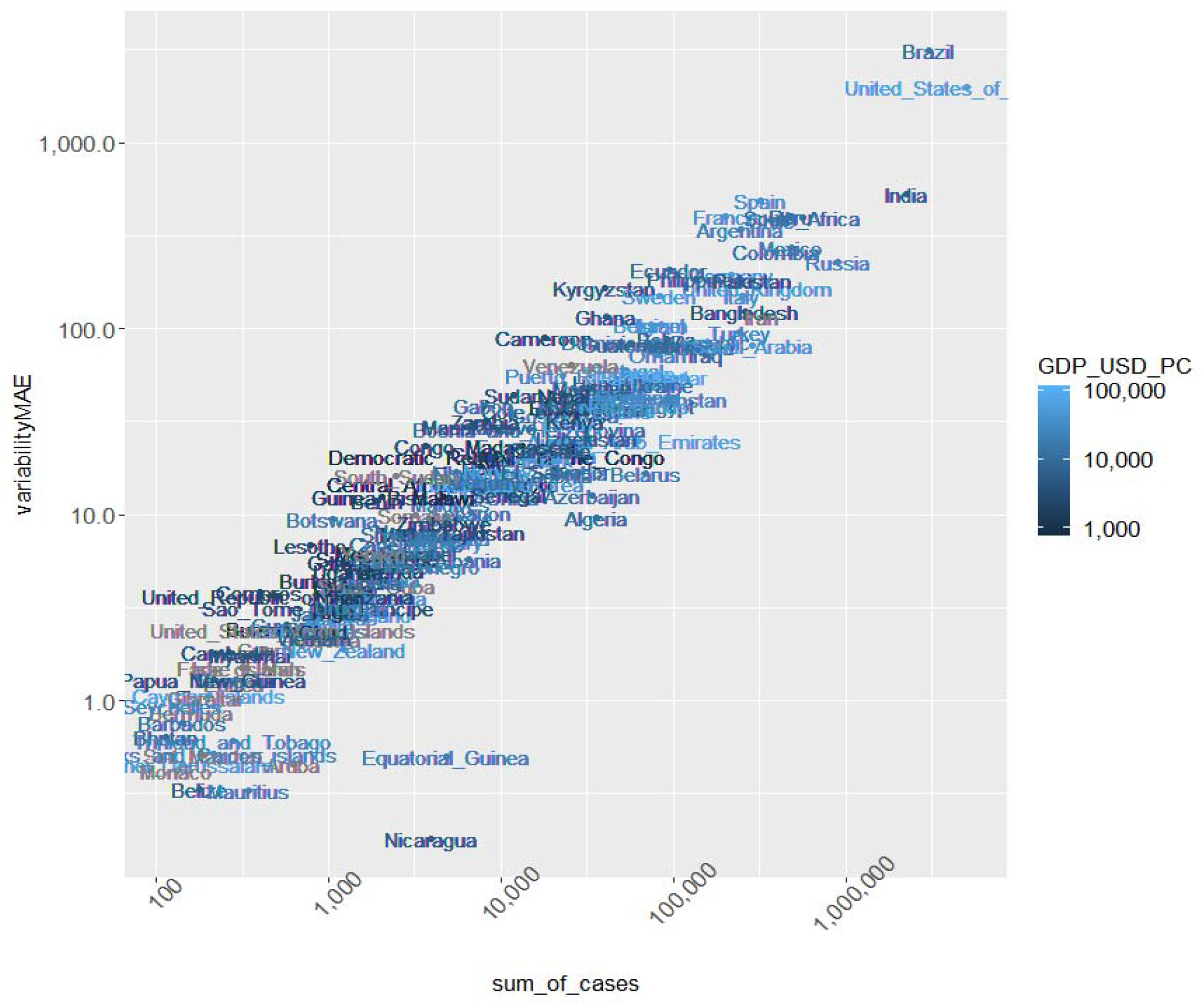
Variability vs total number of cases.

Although the main factor that explains the mean sum of residuals of the Lowess fit, variability is also related to the GDP_per_capita.

A linear model has been fit using log(variability) as the dependent variable and log(sum_of_cases) and log(GDP_per_capita) as predictors, for countries with at least 100 cases as of 2020-08-12. In this model the *R^2^* was as high as 0.888 and the GDP_per_capita reached statistical significance.

**Table 5:** Linear model of variability

| Linear model of variability |  |  |  |  |
| --- | --- | --- | --- | --- |
|  | Estimate | Standard Error | t value | Pr(>\|t\|) |
| (Intercept) | -2.947 | 0.029 | -99.926 | 0.000 |
| $\log(\text{sum\_of\_cases})$ | 0.751 | 0.002 | 473.776 | 0.000 |
| $\log(\text{GDP\_USD\_PC})$ | -0.126 | 0.003 | -41.089 | 0.000 |

*Signif. codes: 0 <= '\*\*\*' < 0.001 < '\*\*' < 0.01 < '\*' < 0.05 < '.' < 0.1 < ' ' < 1*
Residual standard error: 0.5871 on 29162 degrees of freedom Multiple R-squared: 0.8884, Adjusted R-squared: 0.8884 F-statistic: 1.16e+05 on 29162 and 2 DF, p-value: 0.0000

### 5.4 Questions A, B. The impact of spurious peaks: Catalonia

On the 10th May 2020 Catalonia reported 2721 positive cases respective to the previous day; but only 83 of these cases were new cases(55). (This illustrates a problem that might arise when obtaining daily incidence data from the differences of cumulative total cases: Some health authorities often report cumulative total cases every day; these reports in a particular date might include both the new cases diagnosed that day (incident cases) and cases whose date of symptoms or date of diagnosis is not known). Therefore, some cumulative datasets include a disclaimer against the obtention of incidences by differentiation. An older cumulative version of the Spain Ministry of Health COVID-19 dataset (79), accessed on 21-May-2020, has been used; it should be noted that newer versions correct this issue (Sup. material, 5.8).

Here we assess the impact of the spurious peaks *R_t_* estimates and how different smoothing methods deal with them, including smoothing and proper value replacement. The epidemic curves after preprocessing are shown in figure 18 and the transmission curves are shown in figure 19. Standard parameters are used (BDNV), +0% undetected cases).

**Figure 18:**
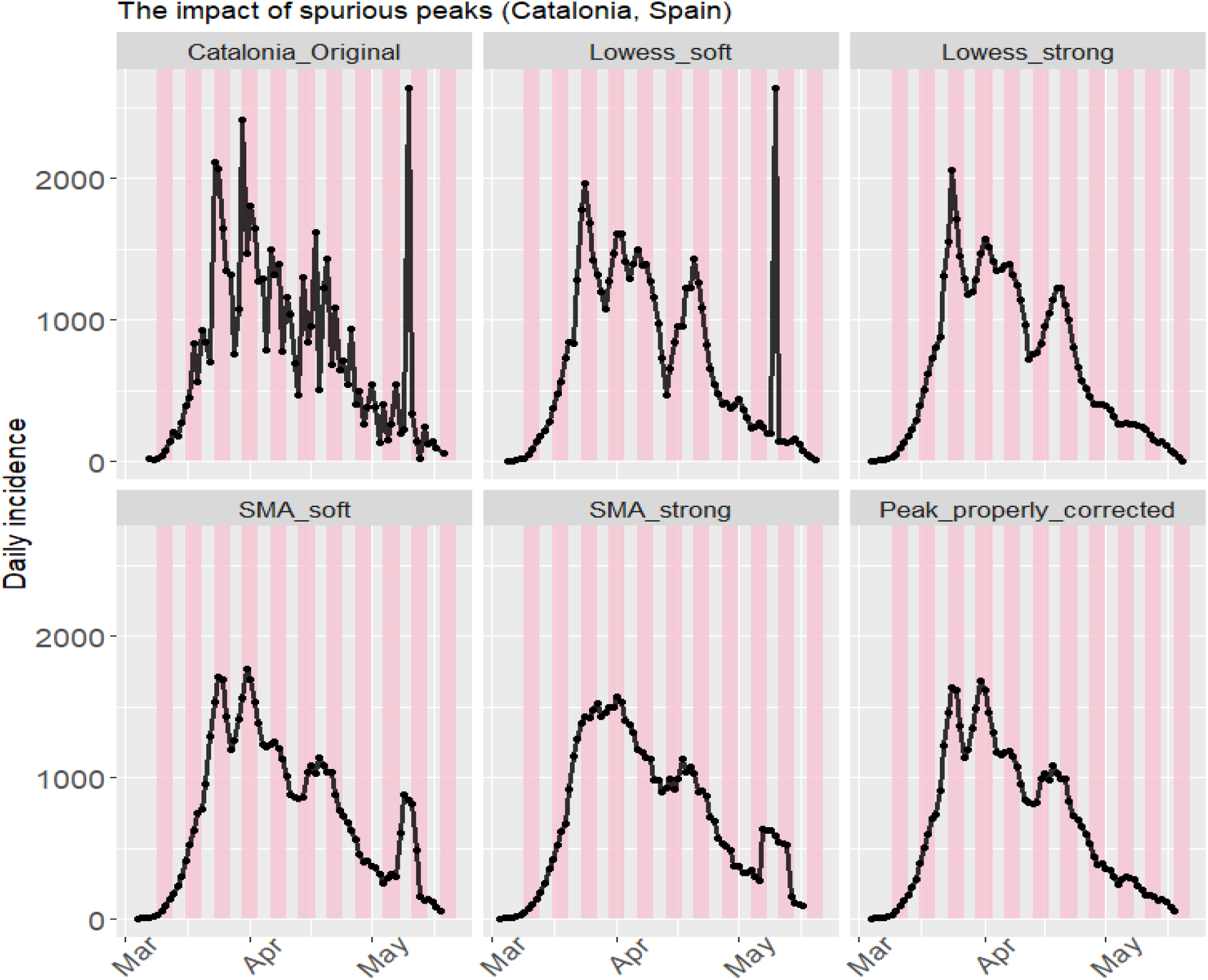
Epidemic curves before and after several smoothing methods.

**Figure 19:**
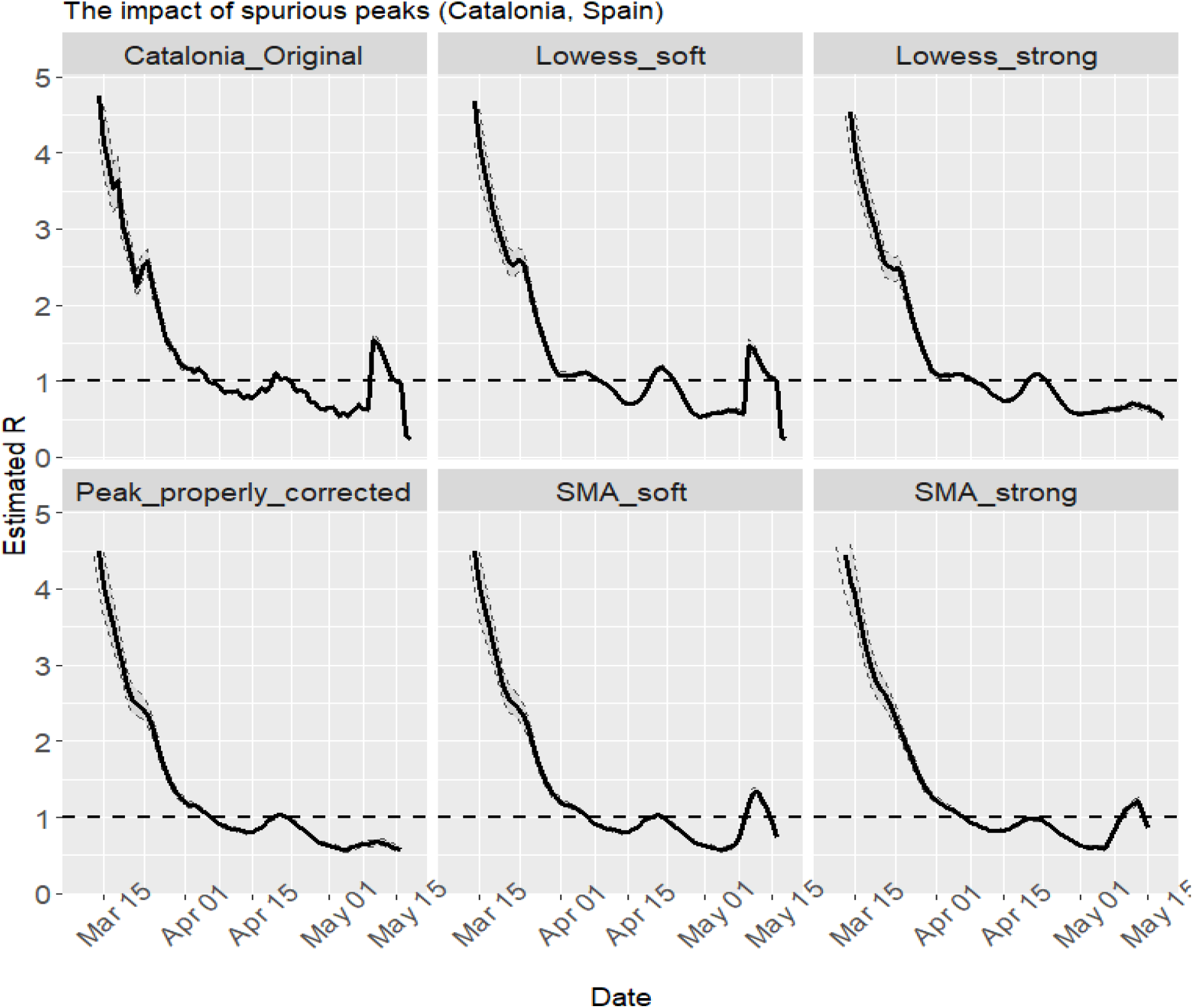
Transmission curves before and after several smoothing methods.

Compared to the SMA, the Lowess smoothing is more tolerant with peaks (and in some settings might not correct them); and the SMA is more influenced by these peaks. This reflects the different philosophy of these methods (mean averaging vs polynomial regression). A second-pass SMA might be useful in some particular cases.

The spurious peak causes spurious *R_t_* estimates if it is not properly corrected. Lowess correction seemed a reasonable method when dealing with these peaks, but proper visually-guided tuning of smoother span is recommended, as small spans could left some peaks. When the SMA correction was applied, spurious *R_t_* estimates were obtained.

### 5.5 Question B. The impact of different preprocessing options

The impact of the different preprocessing options on the transmission curves is evaluated in the figure 20. The following parameters have been used: JHU dataset, dates between 10-March-2020 and 11-August-2020, different smoothing parameters and spans.

**Figure 20:**
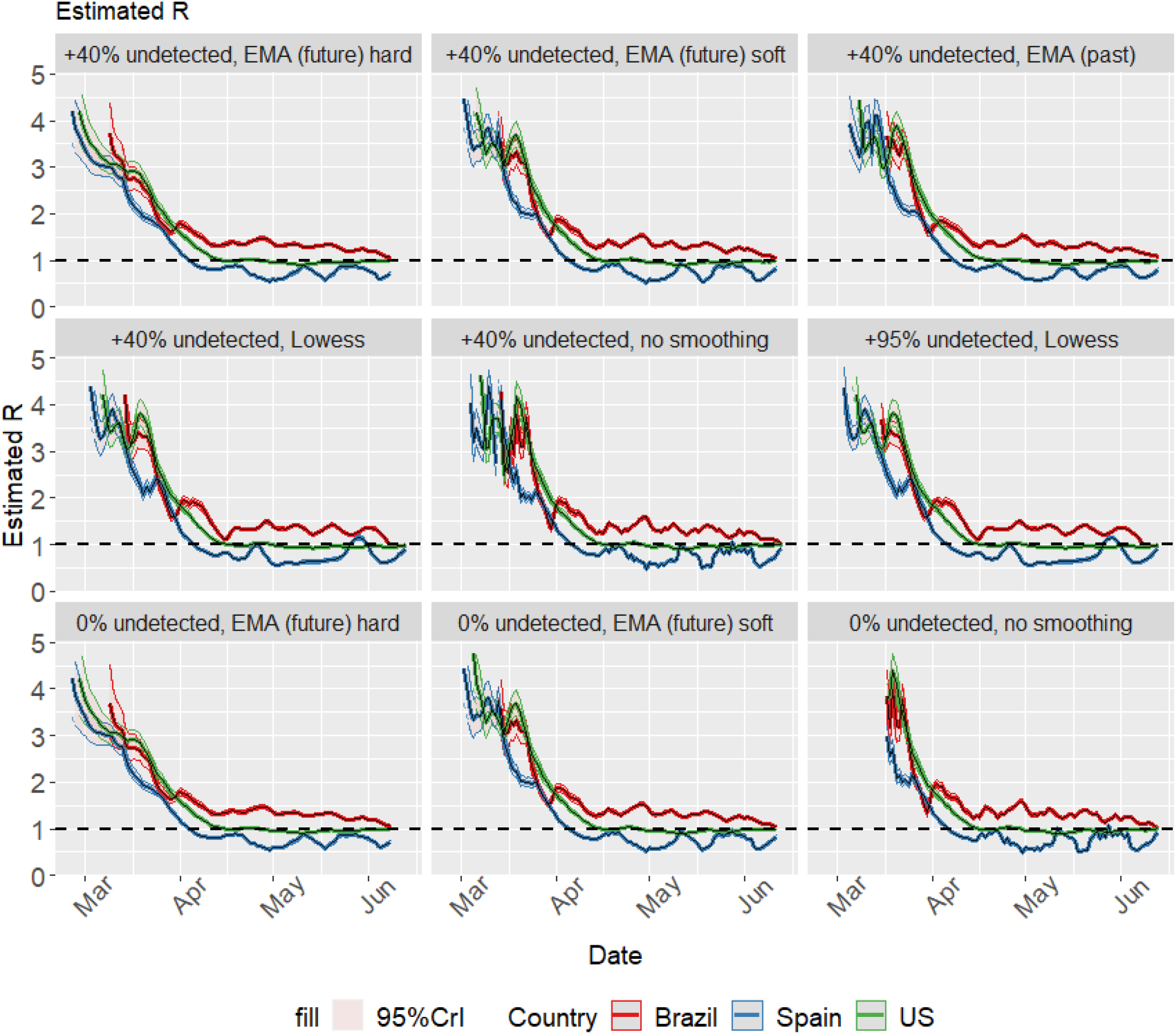
Sensitivity analysis for different preprocessing parameters.

### 5.6 Questions B, C. One-at-a-time sensitivity analysis

We numerically evaluate the differences between different conditions by using the mean daily gap between non-overlapping *R_t_* credible intervals. The Gap *G_t_* at time point *i* between two credible intervals *X* and *Y* obtained with the same data but some different parameter is defined as:

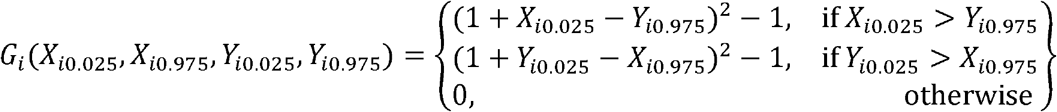

respectively, are the upper and lower posterior limits of the 95% credible interval of the reproduction number at time *i*. The Gap defined here increases with the square of the distance between non-overlapping credible intervals. The existence of this Gap between the limits of the credible intervals reflects there is room for different information and interpretations of the transmission.

The Gap between two transmission curves is the average daily gap, defined as *G — —* the total number of days in which both transmission curves are defined.

#### 5.6.1 Generation interval/serial intervals (GT/si), distributions obtained via MCMC

For each of four countries (Spain, Brazil, USA, United Kingdom) with incidences from the JHU-CSSE dataset (50) between 10-March-2020 and 11-August-2020, the Gap between pairs of *R_t_* curves obtained with different generation time distributions as the only varying parameter is calculated. As described in 2, the datasets from Du et al, Nishiura et al and Zhao et al are used to obtain estimates of the serial interval distribution via MCMC by using the original data; and the generation time estimated parameters described by Ganyani et al with data from Singapore or Tianjin are used to sample generation time distributions. 250 distributions are used due to computational constraints.

Several estimation parameters are kept constant: 0% of undetected rate, backwards distribution of negative values, no specific modification of spurious positive values, centered SMA smoothing with a period or order of 4 days. The results are displayed as triangular matrix of Gap.

**Spain:**

**Table 6:**
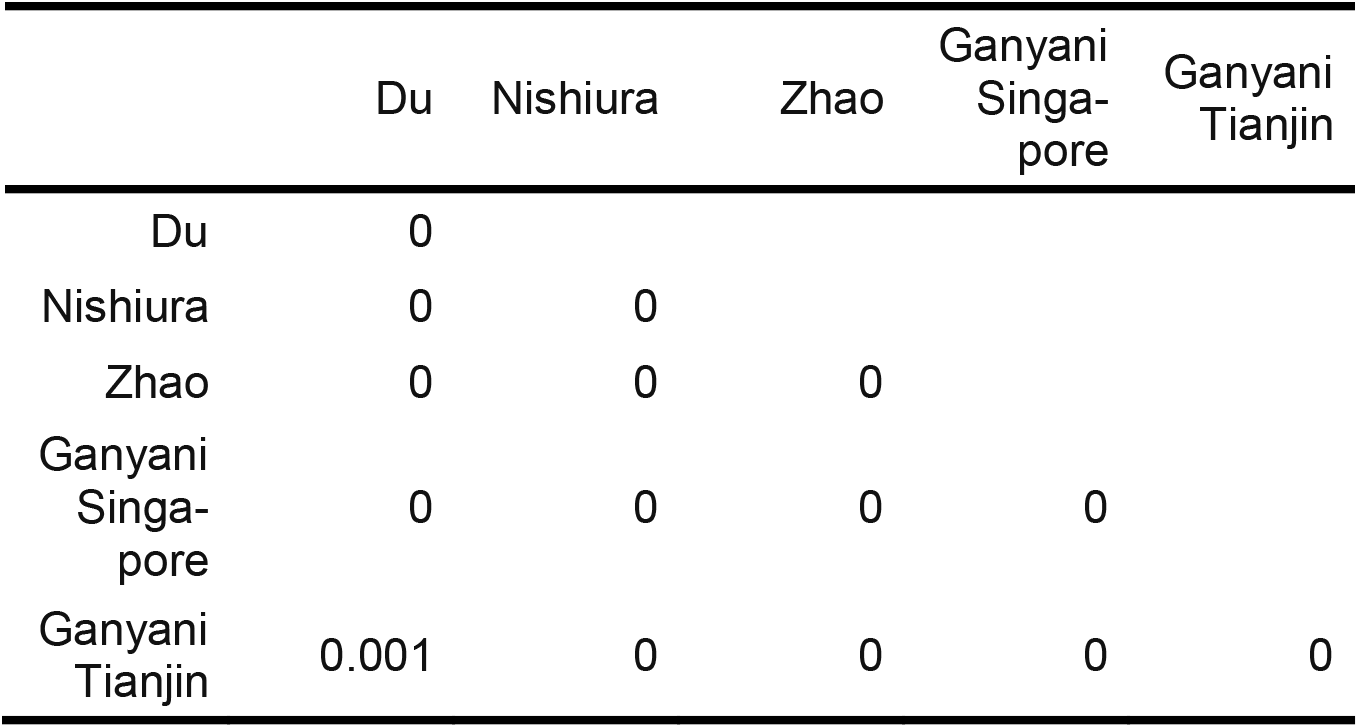
Sens. analysis: GT/si. Spain. Mean daily GAP

**Brazil:**

**Table 7:** Sens. analysis: GT/si. Spain. Mean daily GAP

| Sens. analysis: GT/si. Brazil. Mean daily GAP |  |  |  |  |  |
| --- | --- | --- | --- | --- | --- |
|  | Du | Nishiura | Zhao | Ganyani<br>Singa-<br>pore | Ganyani<br>Tianjin |
| Du | 0 |  |  |  |  |
| Nishiura | 0 | 0 |  |  |  |
| Zhao | 0 | 0 | 0 |  |  |
| Ganyani<br>Singa-<br>pore | 0 | 0 | 0 | 0 |  |

Sens. analysis: GT/si. Brazil. Mean daily GAP
|  | Du | Nishiura | Zhao | Ganyani<br>Singa-<br>pore | Ganyani<br>Tianjin |
| --- | --- | --- | --- | --- | --- |
| Ganyani<br>Tianjin | 0 | 0 | 0 | 0 | 0 |

**United States:**

**Table 8:**
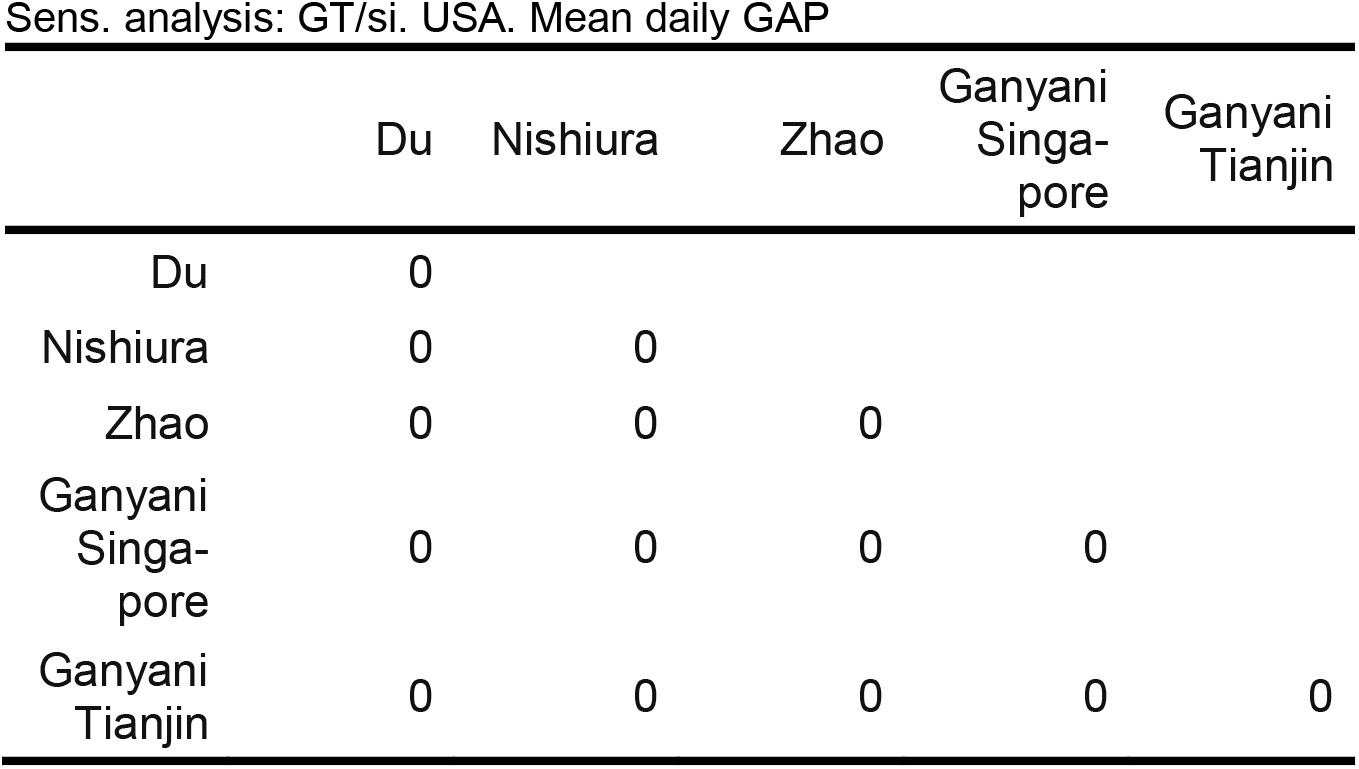
Sens. analysis: GT/si. USA. Mean daily GAP

**United Kingdom:**

**Table 9:**
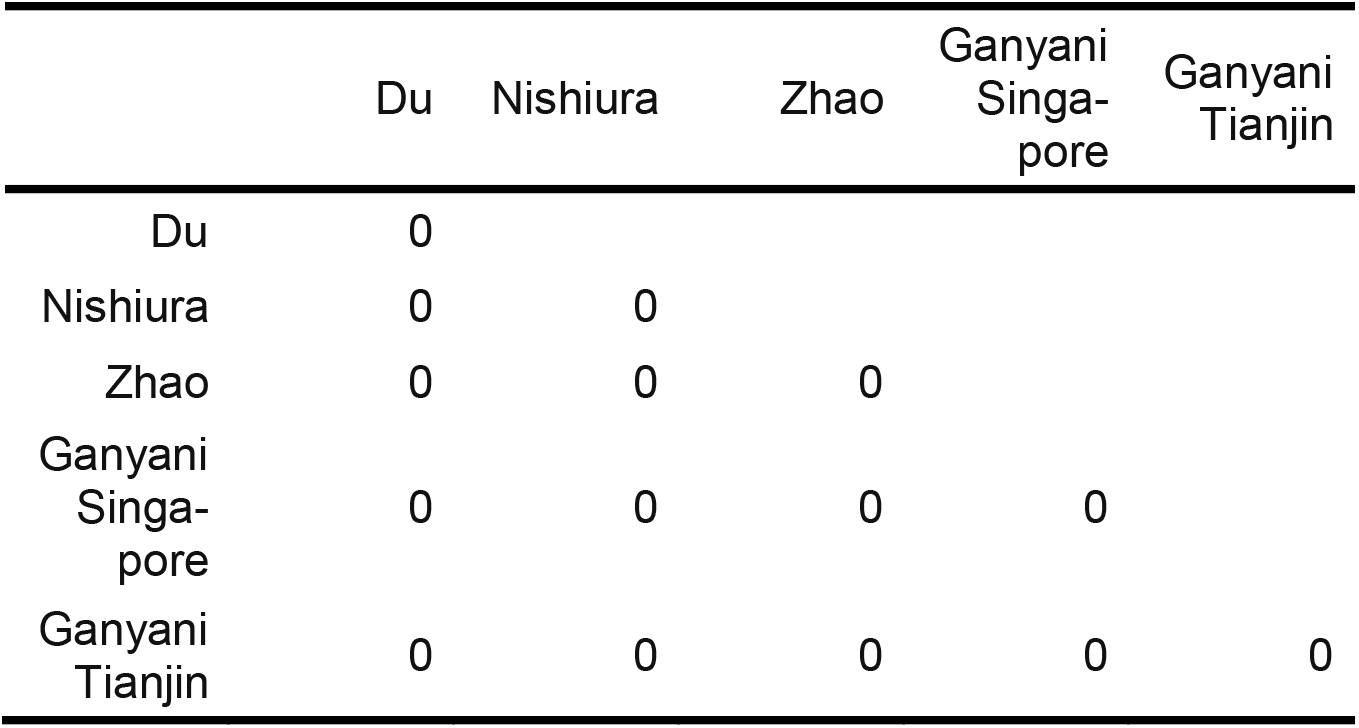
Sens. analysis: GT/si. United Kingdom. Mean daily GAP

#### 5.6.2 Generation interval/serial intervals (GT/si), point estimates

This section is identical to the previous one, with the difference that for each of the five previously mentioned *GT* or *si* sources a single distribution is obtained by using point estimates (Pt), (instead of a sample of *GT* or *si* distributions obtained via MCMC or bootstrap).

**Spain:**

**Table 10:**
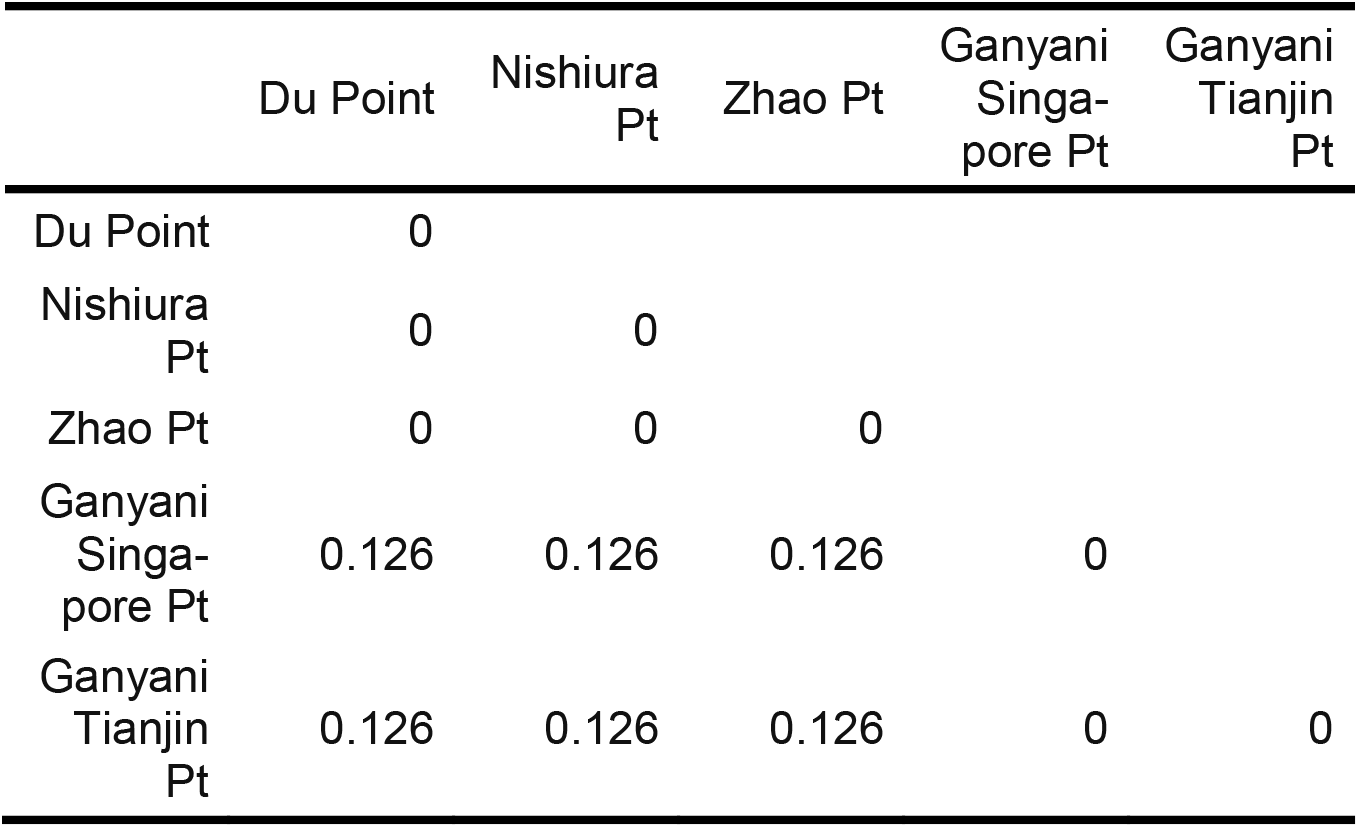
Sens. analysis: GT/si. Spain. Mean daily GAP

**Brazil:**

**Table 11:** Sens. analysis: GT/si. Spain. Mean daily GAP

|  | Du Point | Nishiura<br>Pt | Zhao Pt | Ganyani<br>Singa-<br>pore Pt | Ganyani<br>Tianjin<br>Pt |
| --- | --- | --- | --- | --- | --- |
| Du Point | 0 |  |  |  |  |
| Nishiura<br>Pt | 0 | 0 |  |  |  |
| Zhao Pt | 0 | 0 | 0 |  |  |
| Ganyani<br>Singa-<br>pore Pt | 0.023 | 0.023 | 0.023 | 0 |  |
| Ganyani<br>Tianjin<br>Pt | 0.023 | 0.023 | 0.023 | 0 | 0 |

**United States:**

**Table 12:** Sens. analysis: GT/si. USA. Mean daily GAP

|  | Du Point | Nishiura<br>Pt | Zhao Pt | Ganyani<br>Singa-<br>pore Pt | Ganyani<br>Tianjin<br>Pt |
| --- | --- | --- | --- | --- | --- |
| Du Point | 0 |  |  |  |  |
| Nishiura<br>Pt | 0 | 0 |  |  |  |
| Zhao Pt | 0 | 0 | 0 |  |  |
| Ganyani<br>Singa-<br>pore Pt | 0.133 | 0.133 | 0.133 | 0 |  |

Sens. analysis: GT/si. USA. Mean daily GAP
|  | Du Point | Nishiura Pt | Zhao Pt | Ganyani Singa-pore Pt | Ganyani Tianjin Pt |
| --- | --- | --- | --- | --- | --- |
| Ganyani Tianjin Pt | 0.133 | 0.133 | 0.133 | 0 | 0 |

**United Kingdom:**

**Table 13:**
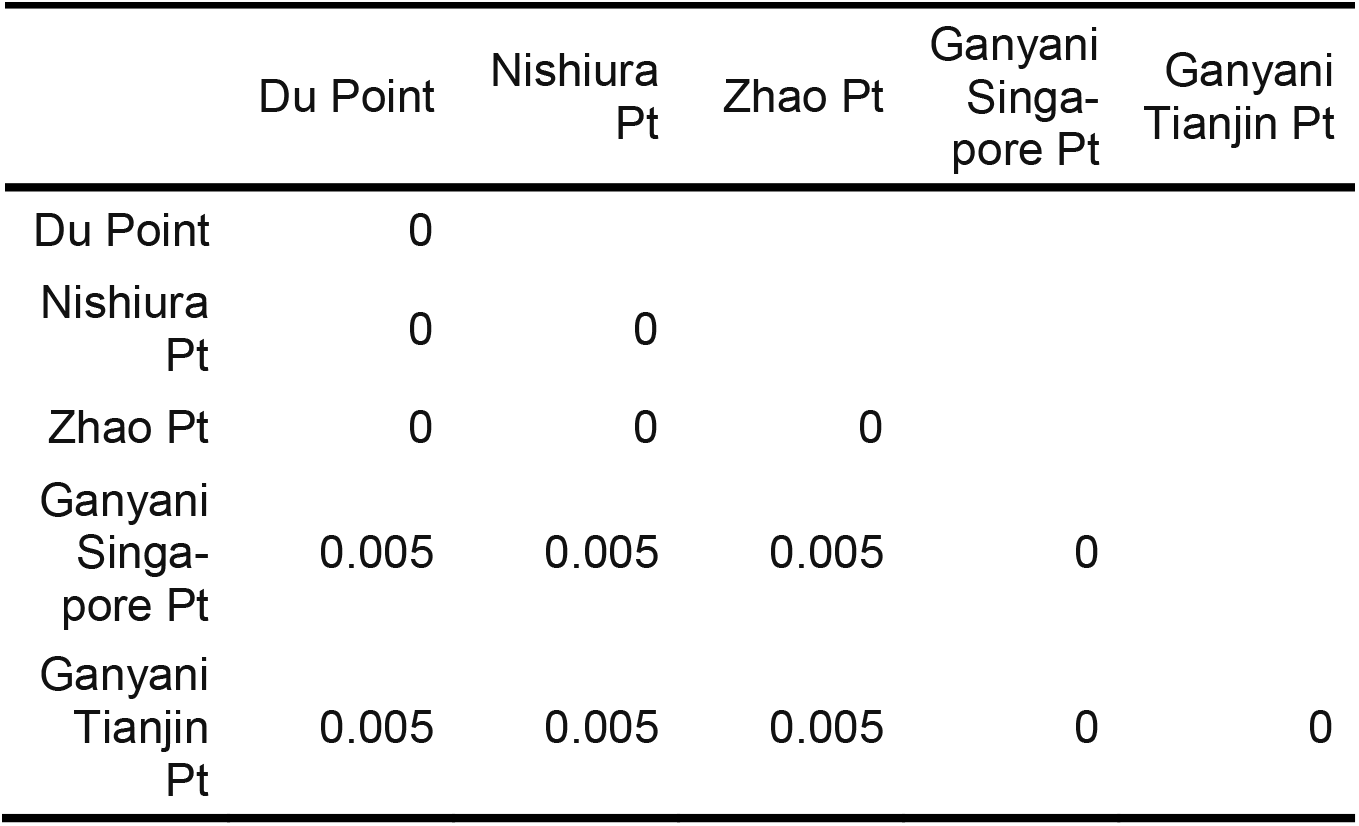
Sens. analysis: GT/si. United Kingdom. Mean daily GAP

|  | Du Point | Nishiura Pt | Zhao Pt | Ganyani Singa-pore Pt | Ganyani Tianjin Pt |
| --- | --- | --- | --- | --- | --- |
| Du Point | 0 |  |  |  |  |
| Nishiura Pt | 0 | 0 |  |  |  |
| Zhao Pt | 0 | 0 | 0 |  |  |
| Ganyani Singa-pore Pt | 0.005 | 0.005 | 0.005 | 0 |  |
| Ganyani Tianjin Pt | 0.005 | 0.005 | 0.005 | 0 | 0 |

When creating the *si* / *GT* distribution with point estimates, there was a Gap between the credible intervals obtained via Generation Time distributions, and the credible intervals obtained via Serial Intervals distributions.

#### 5.6.3 Smoothing

For each of four countries (Spain, Brazil, USA, United Kingdom), the Gap between pairs of *R_t_* curves obtained with different smoothing orders as the only varying parameter is calculated; the smoothing order is the number of points used for each point smoother. Several estimation parameters are kept constant: 0% of undetected rate, backwards distribution of negative values, no specific modification of spurious positive values, centered SMA smoothing, serial interval distribution obtained from Du et al(30). The results are displayed as triangular matrix of Gap.

**Spain:**

**Table 14:** Sens. analysis: Smoothing. Spain. Mean daily GAP

|  | 2 | 5 | 7 |
|---|---|---|---|
| 2 | 0 |  |  |
| 5 | 0 | 0 |  |
| 7 | 0 | 0 | 0 |

**Brazil:**

**Table 15:** Sens. analysis: Smoothing. Brazil. Mean daily GAP

| Sens. analysis: Smoothing. Brazil. Mean daily GAP |  |  |  |
| --- | --- | --- | --- |
|  | 2 | 5 | 7 |
| 2 | 0 |  |  |
| 5 | 0 | 0 |  |
| 7 | 0 | 0 | 0 |

**USA:**

**Table 16:** Sens. analysis: Smoothing. USA. Mean daily GAP

| Sens. analysis: Smoothing. USA. Mean daily GAP |  |  |  |
| --- | --- | --- | --- |
|  | 2 | 5 | 7 |
| 2 | 0 |  |  |
| 5 | 0 | 0 |  |
| 7 | 0 | 0 | 0 |

**UK:**

**Table 17:** Sens. analysis: Smoothing. United Kingdom. Mean daily GAP

| Sens. analysis: Smoothing. United Kingdom. Mean daily GAP |  |  |  |
| --- | --- | --- | --- |
|  | 2 | 5 | 7 |
| 2 | 0 |  |  |
| 5 | 0 | 0 |  |
| 7 | 0 | 0 | 0 |

#### 5.6.4 Undetected ratio

For each of four countries (Spain, Brazil, USA, United Kingdom), the Gap between pairs of *R_t_* curves obtained with different proportions of undetected ratio (0%, 40%, 95%; which is used to calculate the total estimated incidence) as the only varying parameter is calculated; the smoothing order is the number of points used for each point smoother. Several estimation parameters are kept constant: 0% of undetected rate, backwards distribution of negative values, no specific modification of spurious positive values, centered SMA smoothing, serial interval distribution obtained from Du et al(30). The results are displayed as triangular matrix of Gaps.

**Spain:**

**Table 18:**
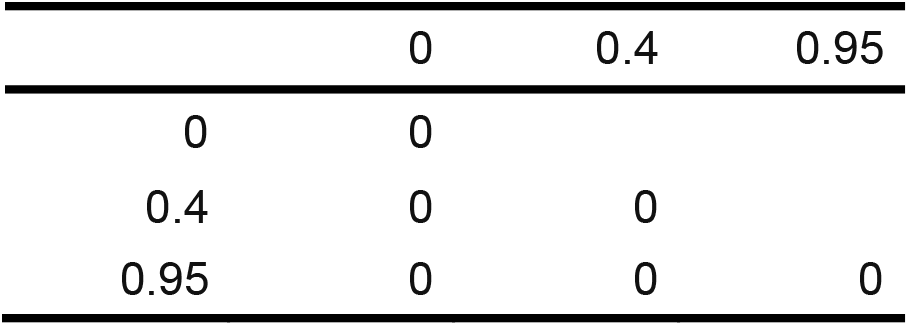
Sens. analysis: Smoothing. United Kingdom. Mean daily GAP

**Brazil:**

**Table 19:**
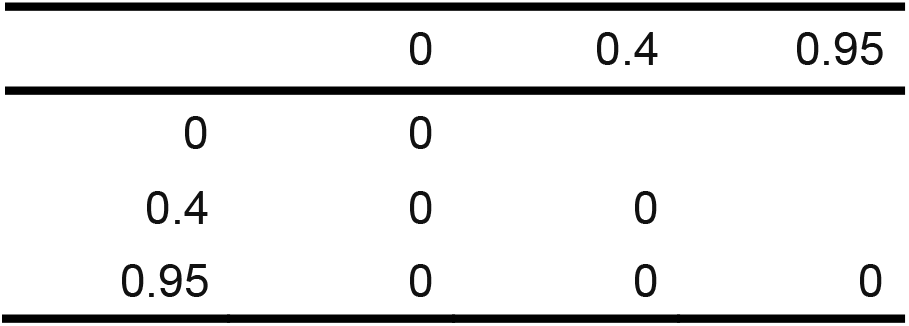
Sens. analysis: Smoothing. Brazil. Mean daily GAP

**USA:**

**Table 20:**
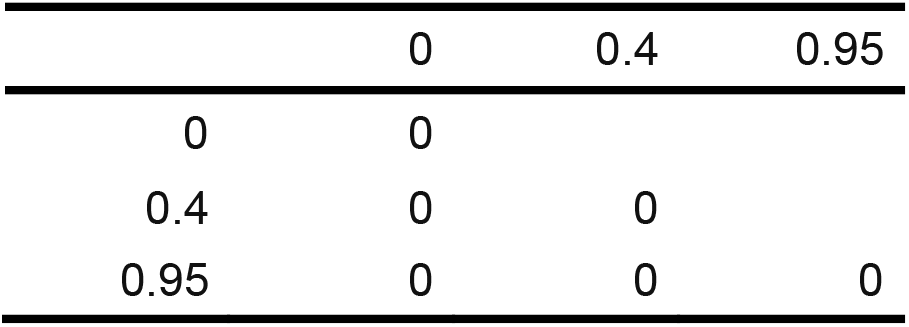
Sens. analysis: Smoothing. USA. Mean daily GAP

**UK:**

**Table 21:**
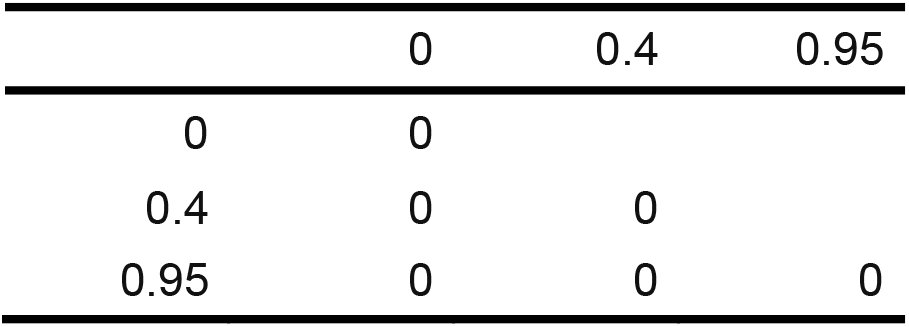
Sens. analysis: Smoothing. United Kingdom. Mean daily GAP

As apparent from the GAPs, the GT/si distribution chosen is the factor that most influences the differences in transmission curves. However the mean daily differences were small.

### 5.7 Question C, E. Spain regional estimates, three-day windows

As a counterpart for the weekly estimates for Spanish regions, three-day estimates are obtained and displayed here for reference purposes (21). The same setting as are used as in the previous case.

**Figure 21:**
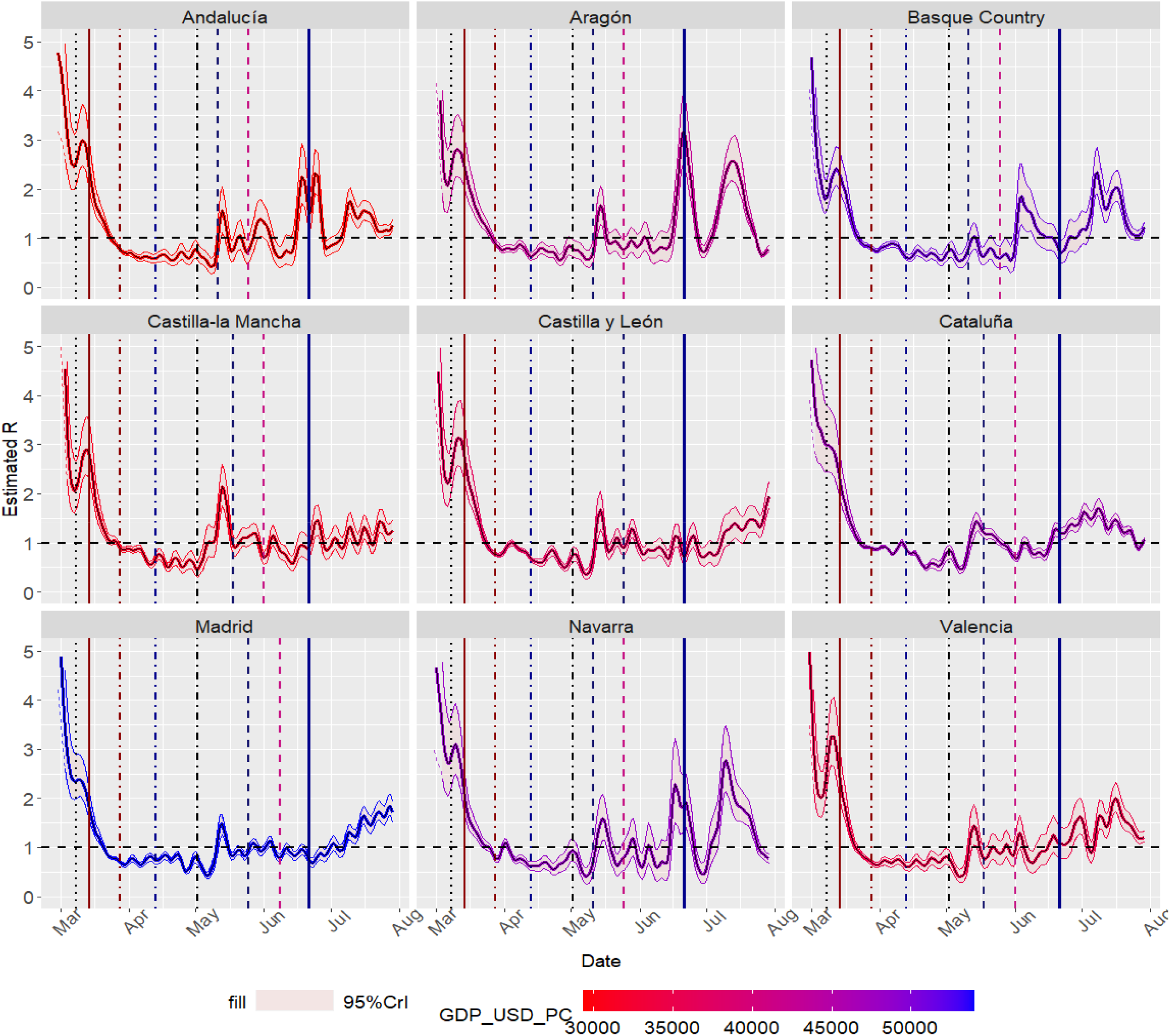
R_t_ estimates for several Spanish Autonomous Communities. Three-day estimation windows. Highlighted dates: 2020-03-08 (8M), 2020-03-14 (State of Alarm), 2020-03-28 (Economic stop), 2020-04-13 (Partial economic restart), 2020-05-04 (Return to ‘normal’, phase 0),phases 1 and 2, 2020-06-21 (End of the state of Alarm)

Given the significant weekday bias, three day windows are not recommended.

Daily estimates are more susceptible to noise and show a striking weekday/weekend bias, and don’t provide much useful information when compared to weekly *R_t_* estimates.

### 5.8 Question A, E. Spain: Public health surveillance data source

In Spain the New Normality Transition Plan established the daily automated and individual notification of cases to the national Health Surveillance authorities for some Epidemic indicators(56); thus highlighting the importance of quality individualized reports of cases, instead of plain cumulative reports. The switch between cumulative reports and individualized epidemiological reports was not easy and exemplifies the challenges that many countries faced(57). The Spanish Public Health Surveillance Network (RENAVE) collects COVID-19 reports via the SIVIES system(74), and symptoms-onset dates are used to calculate the incidence. The median diagnosis delay (interval between symptoms onset and report) was initially 6 days(58)(reported on 2020-05-21), but later descriptions were smaller (3 days, reported on 2020-08-06)(54); these values are used for imputing missing onset dates (see https://cnecovid.isciii.es/covidl9/#documentaci%C3%B3n-y-datos for details). The purpose of this section is to show the improvements in daily incidences when proper epidemiological reports are used.

Figure 22 compares the incidence reported via the RENAVE and the deprecated previous cumulative report of the government. Figure 23 displays the transmission curve (parameters include SMA smoothing, 4 day smoothing span and 40% proportion of undetected cases).

**Figure 22:**
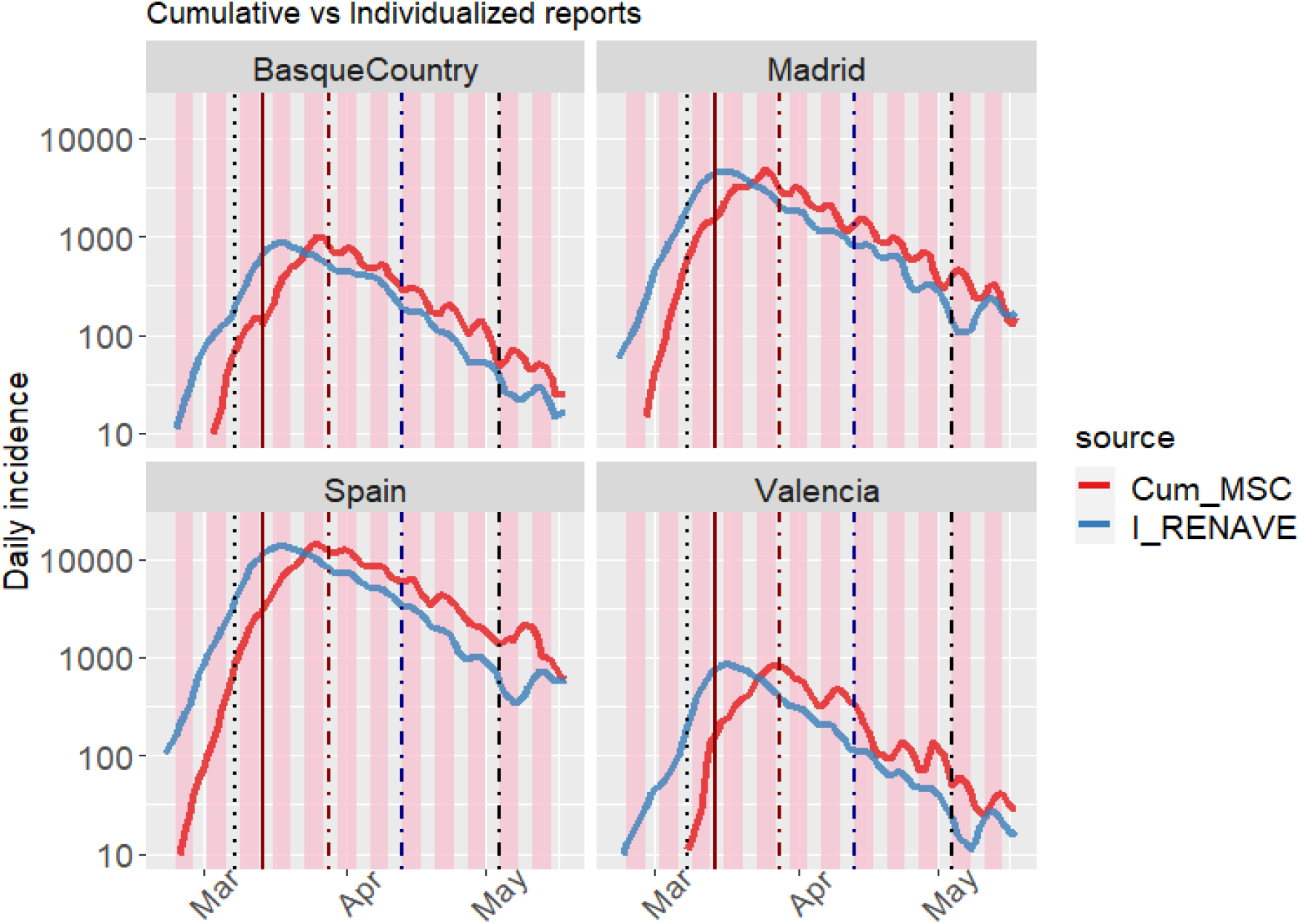
Incidences for Spain and regions (Initial cumulative reports (Cum_MSC) vs individualized rep orts(I_RENAVE)). Highlighted dates: 2020-03-08 (8M), 2020-03-14 (State of Alarm), 2020-03-28 (Economic stop), 2020-04-13 (Partial economic restart), 2020-05-04 (Return to ‘normal’, phase 0)

**Figure 23:**
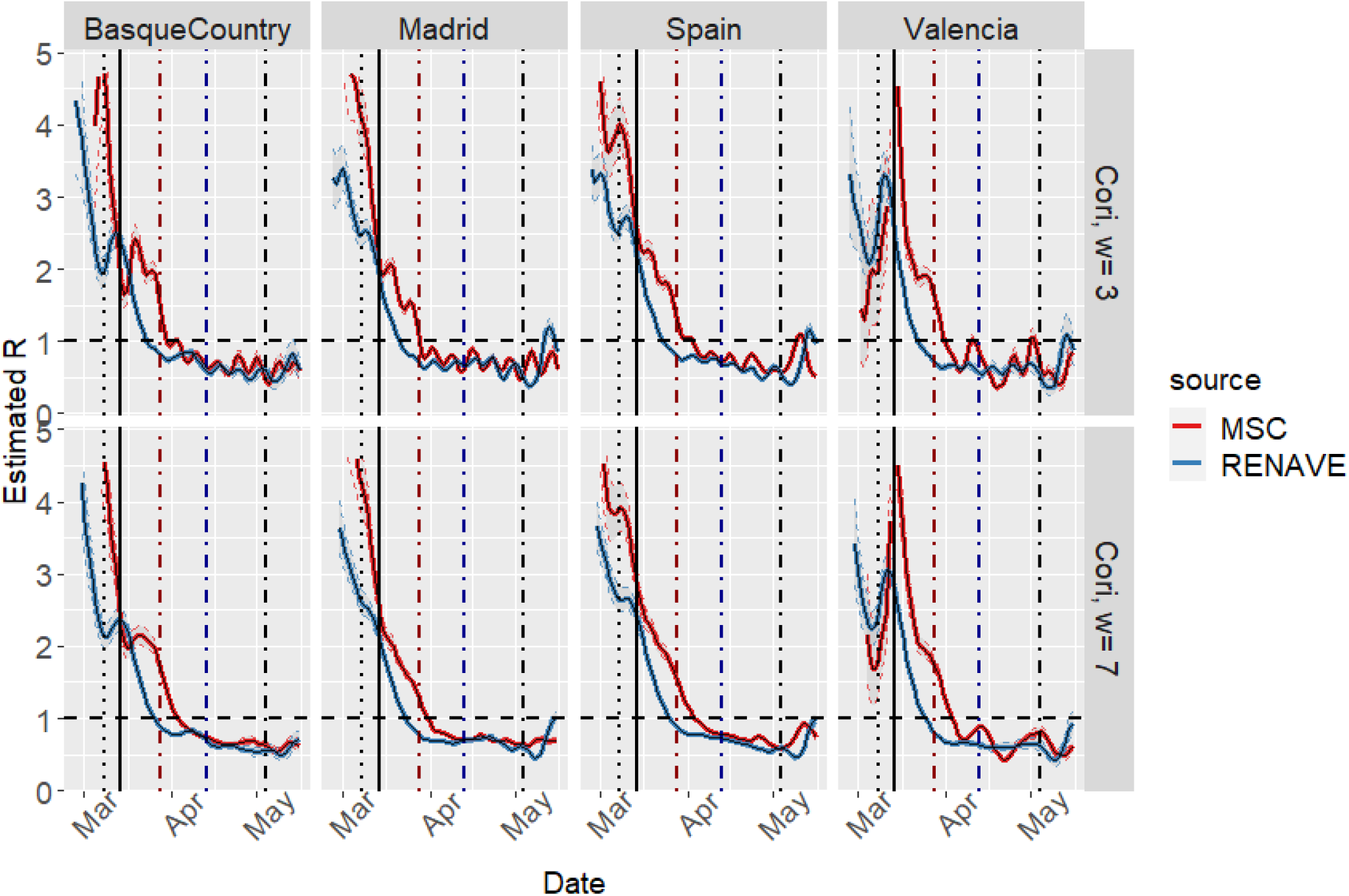
R_t_ estimates for Spain and regions (Government vs public health data source). Highlighted dates: 2020-03-08 (8M), 2020-03-14 (State of Alarm), 2020-03-28 (Economic stop), 2020-04-13 (Partial economic restart), 2020-05-04 (Return to ‘normal’, phase 0)

The epidemiological symptoms-onset report is less prone to weekday bias (which was particularly important when the incidence was decreasing) and exhibits the effect of the diagnosis/reporting delay in the cumulative data. The variability, as defined in 5.3, is greater in the initial cumulative dataset (314) compared to the improved RE NAVE dataset (174).

### 5.9 Question B. Catalonia: Confirmed vs suspected cases, weekend smoothing

The official Catalonia open data COVID-19 repository(45) has been analyzed. This data includes several time series corresponding to different case definitions: Positive PCR, antibodies test, rapid tests, suspected cases; thus providing insights on the evolution of suspected vs confirmed cases over time. Additionally, the impact of smoothing and the **weekday bias** (a decrease in the assumed daily incidence in the weekends due to the laboratories having reduced activity).

The application and code have been used to obtain epidemic and transmission curves. Mild smoothing is defined as **3** days of smoothing with the “centered SMA” method, and Strong smoothing is defined as **7** days of smoothing with the “centered SMA” method; other parameters include: negative values are distributed backwards before smoothing, spurious positive peaks are not removed and the undetected rate is zero. Figure 24 displays the epidemic curves after preprocessing and figure 25 displays the transmission curves.

**Figure 24:**
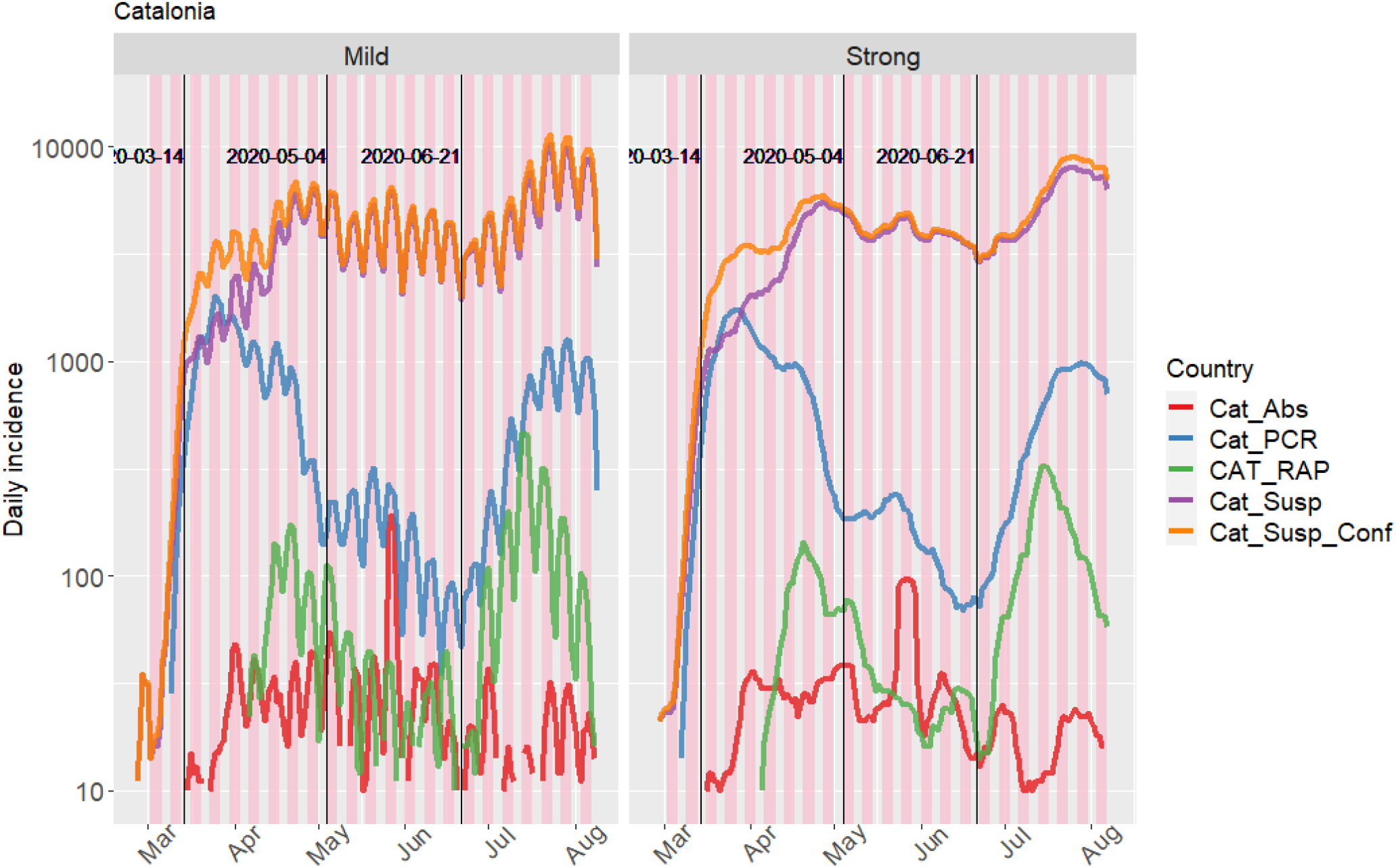
Epidemic curve of Catalonia. Highlighted dates: 2020-03-14 (State of Alarm); 2020-05-04 (Phase zero, reopening); 2020-06-21 (End of the State of Alarm)

**Figure 25:**
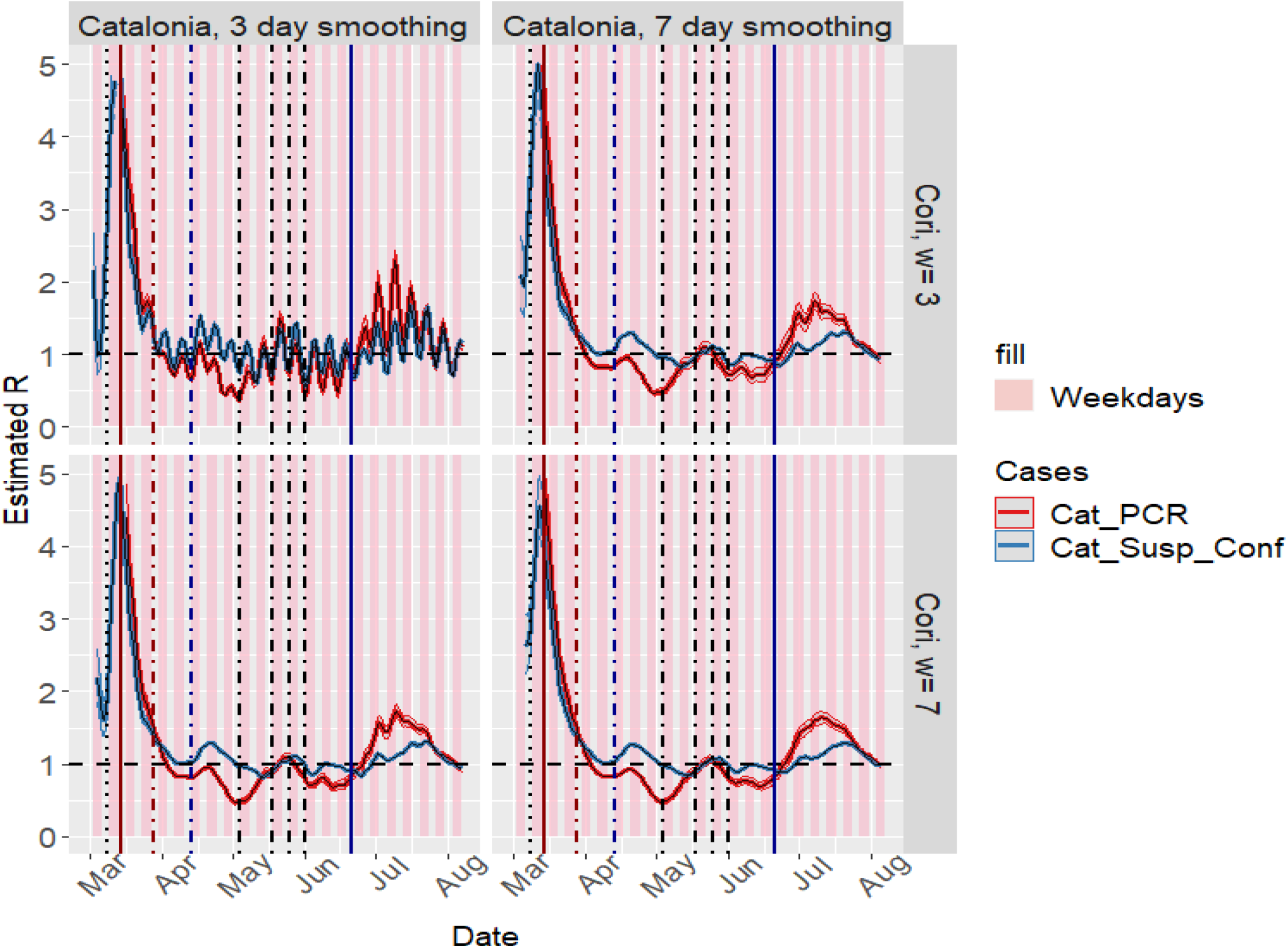
R_t_ estimates for Catalonia. Highlighted dates: 2020-03-08 (8M), 2020-03-14 (State of Alarm), 2020-03-28 (Economic stop), 2020-04-13 (Partial economic restart), 2020-05-04 (Return to ‘normal’ (phase 0)), gradual reopening, 2020-06-21 (End of State of Alarm)

In Catalonia epidemic curves show a significant show a pattern seen in other datasets: the incidence decreases in weekends, maybe due to laboratory workdlows and the role of physicians and epidemiologists.

The number of positive PCRs has decreased over time after the epidemic peak, signaling an effective epidemic control. On the other hand suspected cases have kept increasing after the epidemic peak while the confirmed cases declined; this could be explained by many reasons/hypotheses (non-specific symptoms associated with COVID-19, seasonal allergies, increased public awareness, increased Primary Care and Public Health throughput, a shifting from care-at-home to early testing…), but in any case it demonstrates Suspected cases cannot be reliably used as a surrogate for undetected cases or total cases, at least in this dataset). Truly underreported cases should specially increase near the epidemic peak due to both asymptomatic cases and laboratory limitations, but here “suspected” cases increase after the epidemic peak.

Variability exists between regions and countries regarding the management of these suspected cases. In the epidemic peak many countries and guidelines don’t require a PCR is performed in all suspected cases due to constrained resources.

Both smoothing the time series and using longer time windows for the estimations can reduce the impact of weekend fluctuations in the epidemic curves and in the transmission curves.

### 5.10 Question B. Belgium: The impact of smoothing

The official Belgium dataset, collected by the Sciensano institute, (80) has been analyzed using the application and code in order to assess the impact of smoothing and the “weekday bias”. Mild smoothing is defined as **3** days of smoothing with the “centered SMA” method, and Strong smoothing is defined as **7** days of smoothing with the “centered SMA” method; other parameters: negative values are distributed backwards before smoothing, spurious positive peaks are not removed and the undetected rate is zero. Figure 26 displays the epidemic curves after preprocessing and figure 27 displays the transmission curves.

**Figure 26:**
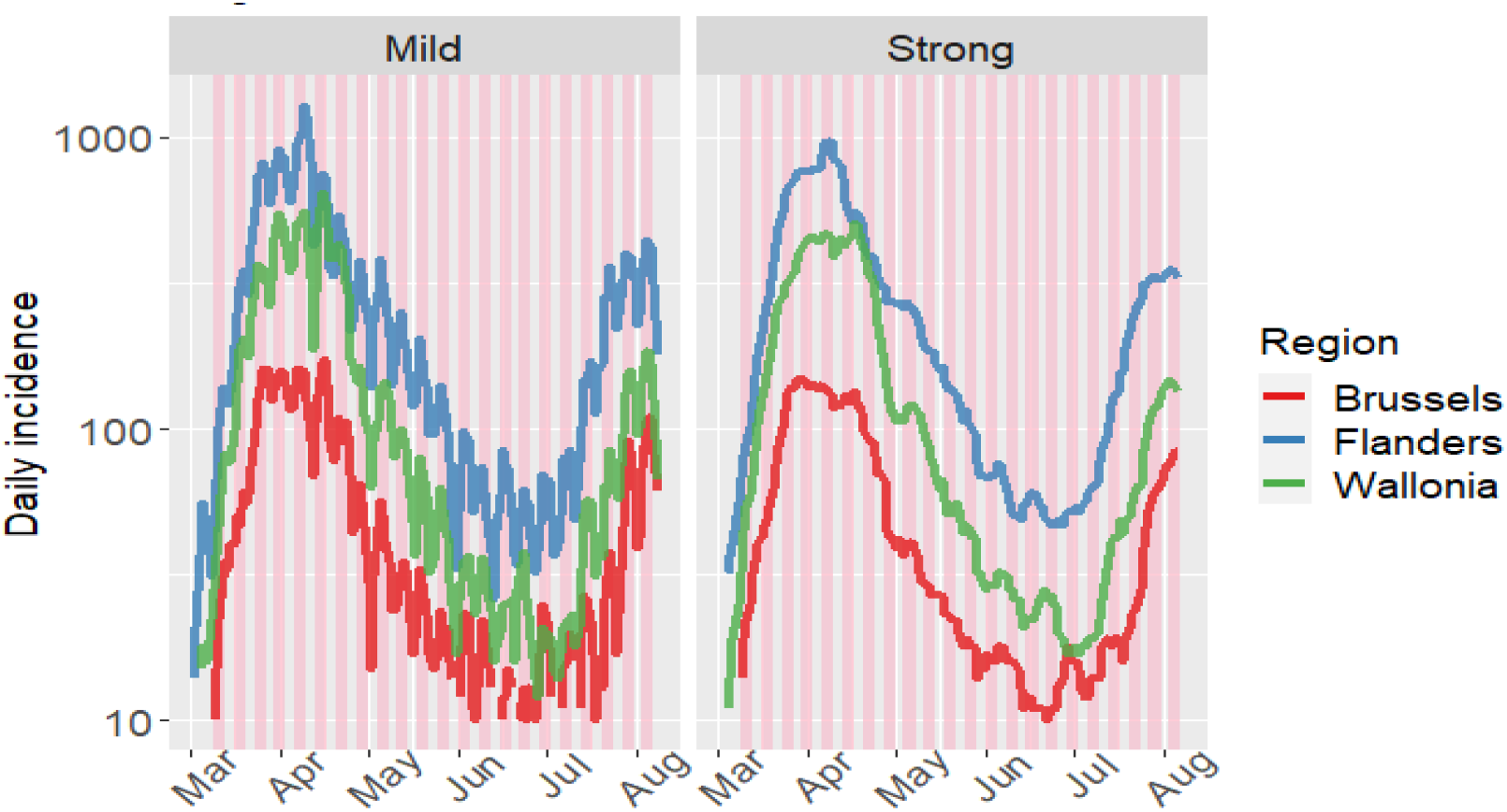
Epidemic curves of Belgium.

**Figure 27:**
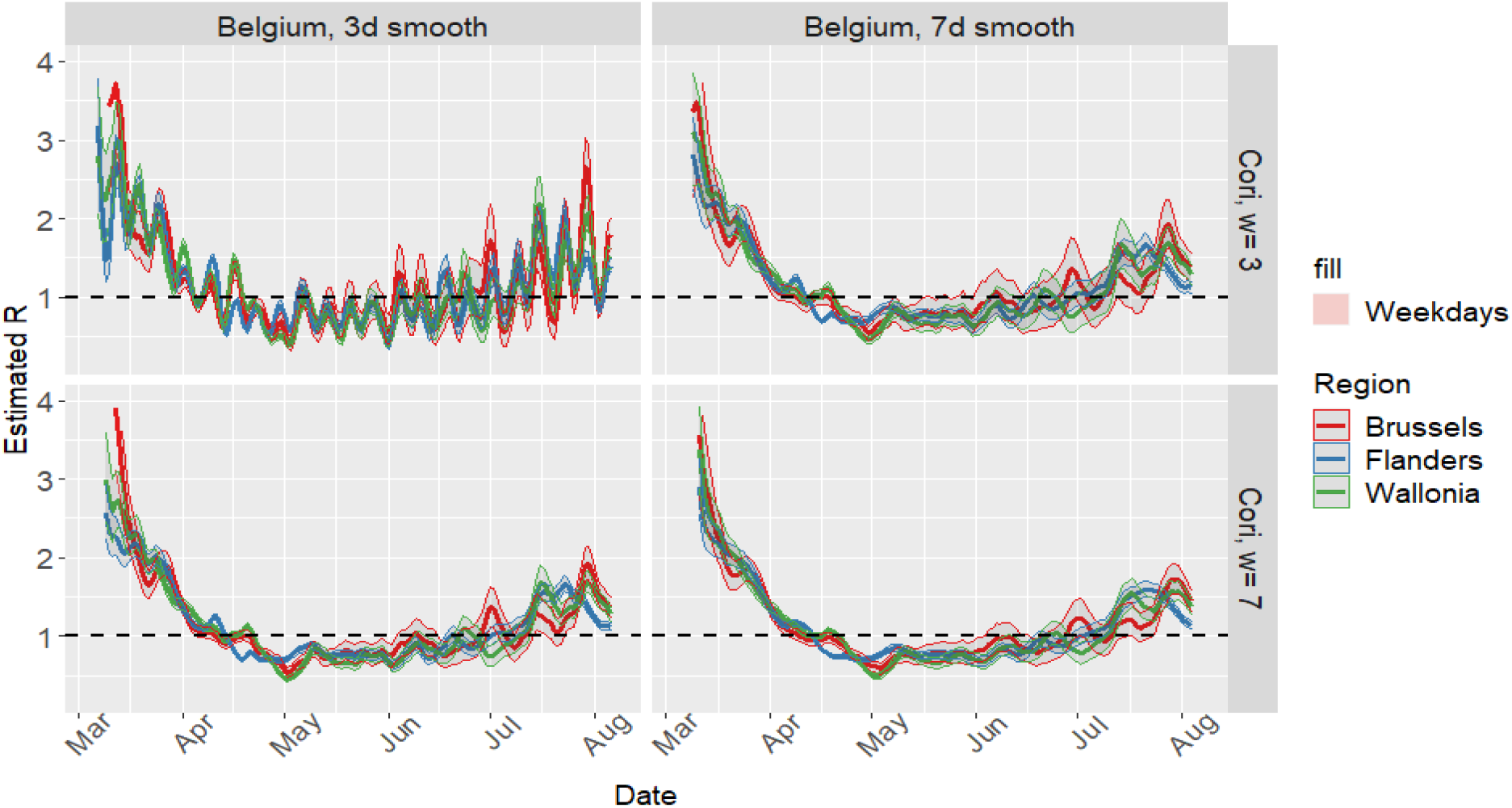
Transmission curves of Belgium.

As seen in the case of Catalonia, using smaller window periods for the *R_t_* estimation is equivalent to using smaller SMA smoothing periods. The differences between Belgian divisions are small, and the *R_t_* intervals become wider as the incidence decreases.

### 5.11 Question E. USA: The state estimates of a developed country

The state-level number of cases from the USA between 15-March-2020 and 11-August-2020 has been analyzed; dataset is from The New York Times, based on reports from state and local health agencies(81). Preprocessing options include SMA centered smoothing, 7 day as smoothing span. Figure 28 displays the epidemic curve after preprocessing and figure 29 displays the transmission curves; in the transmission curve the states have been coloured according to their GDP per capita (purchase power parity, current prices) (44) and in the epidemic curve the date of the death of George Floyd has been highlighted, after which riots ensued (section 3.6).

**Figure 28:**
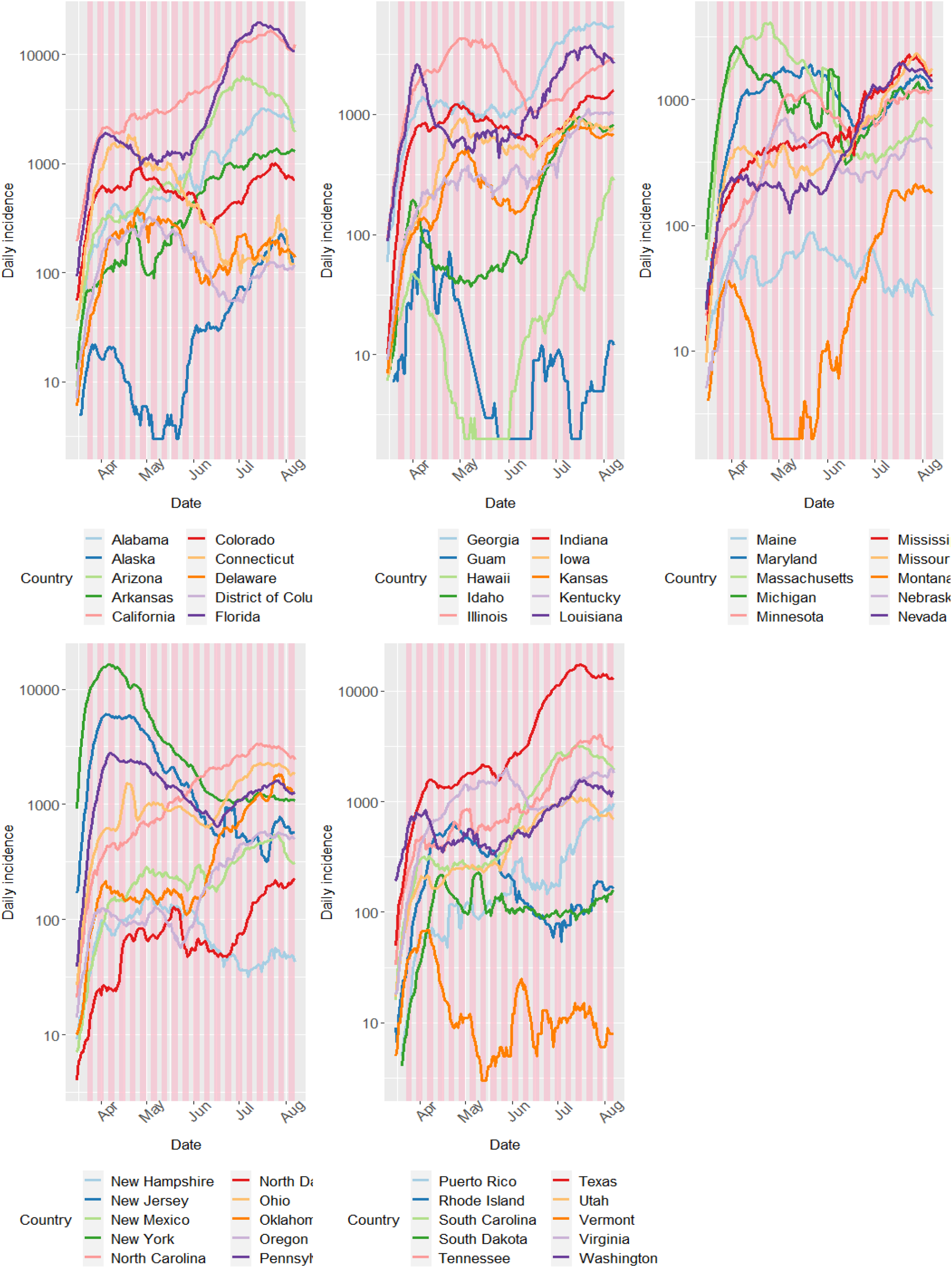
COVID-19 incidence in several USA states.

**Figure 29:**
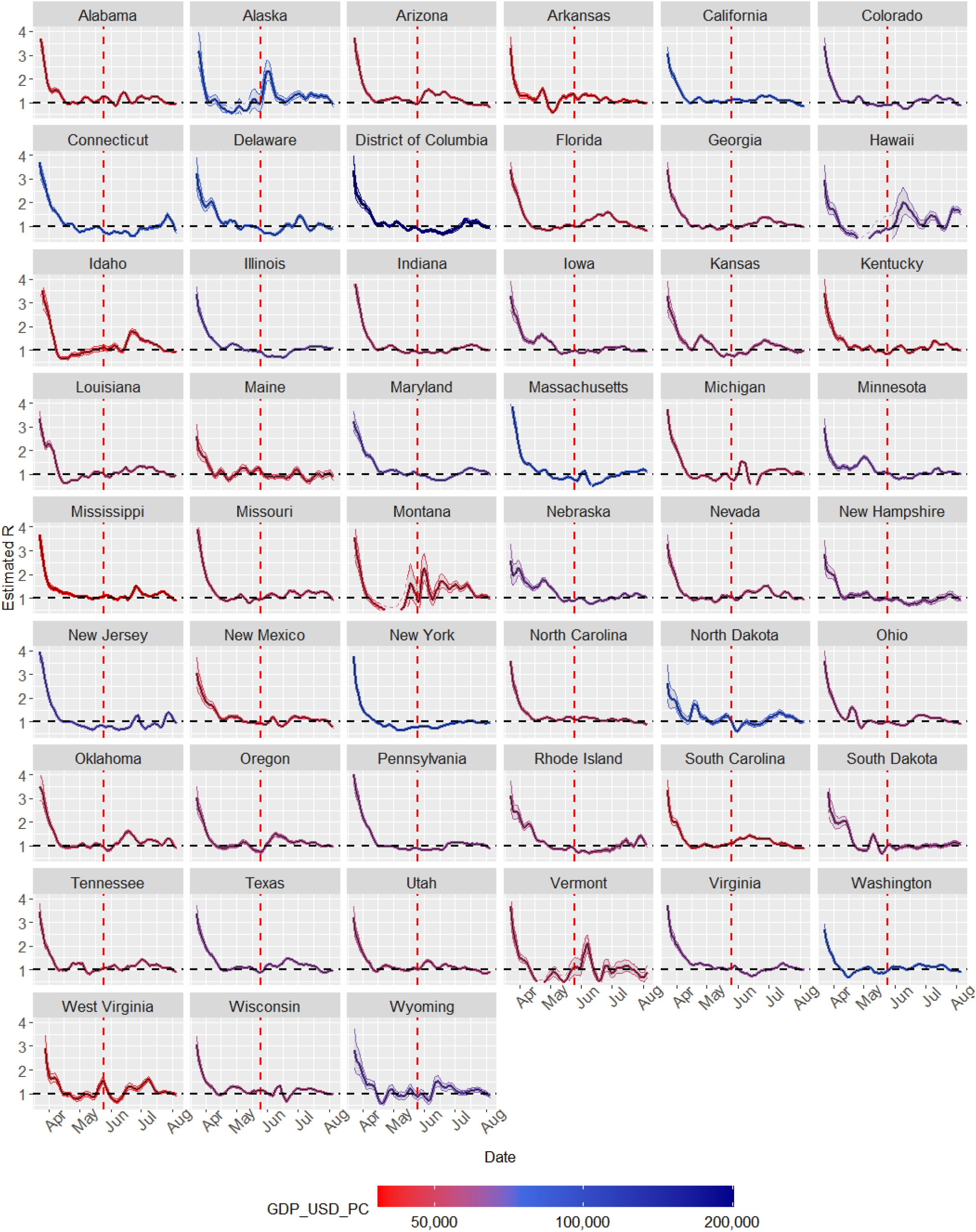
R_t_ estimates for USA states.

Some wealthy and densely-populated states (New York, New Jersey, Massachusetts, Connecticut, Columbia) have shown higher incidences than other states and they kept the epidemic under control from April to June, although most of them reached a *R_t_* close to 1 by the start of July. In other states (California, Washington, North Dakota, Delaware) epidemic control was slightly worse. In some states with lower GDP per capita the epidemic control was less consistent (Alabama, Arizona, Arkansas, South Carolina, West Virginia). However, this findings are subtle and not very well-defined; so there are other factors influencing between-state differences in transmission.

New York State has arguably reached the best COVID-19 control so far, albeit this could be explained by the peak of daily incidences:* 10000 in April. It could be speculated that some particular peaks/valleys could be attributed to real-life events, such as the meat packing plants cases in Nebraska in April. Looking for life-events to attribute them to peaks could lead to flawed conclusions due to the confirmation bias.

Many states showed an incipient rebound in transmission in late June: Florida, Oklahoma, South Carolina, Oregon…); small increases in the *R_t_* above 1 in this period were associated with thousands of new cases (Texas, California, Arizona, Florida).

### 5.12 Question E. Perú: The regional estimates of a developing country

Using the MINSAL incidence data between 15-March-2020 and 11-August-2020, compiled by JM Castagnetto(76), the regional epidemic curves (figure (30)) and transmission curves (figure 31) of Perú have been obtained. Preprocessing parameters include: SMA smoothing, 7 days as smooth span, +40% proportion of undetected cases. In the transmission curve the states have been coloured according to their GDP per capita (purchase power parity, current prices) (44).

**Figure 30:**
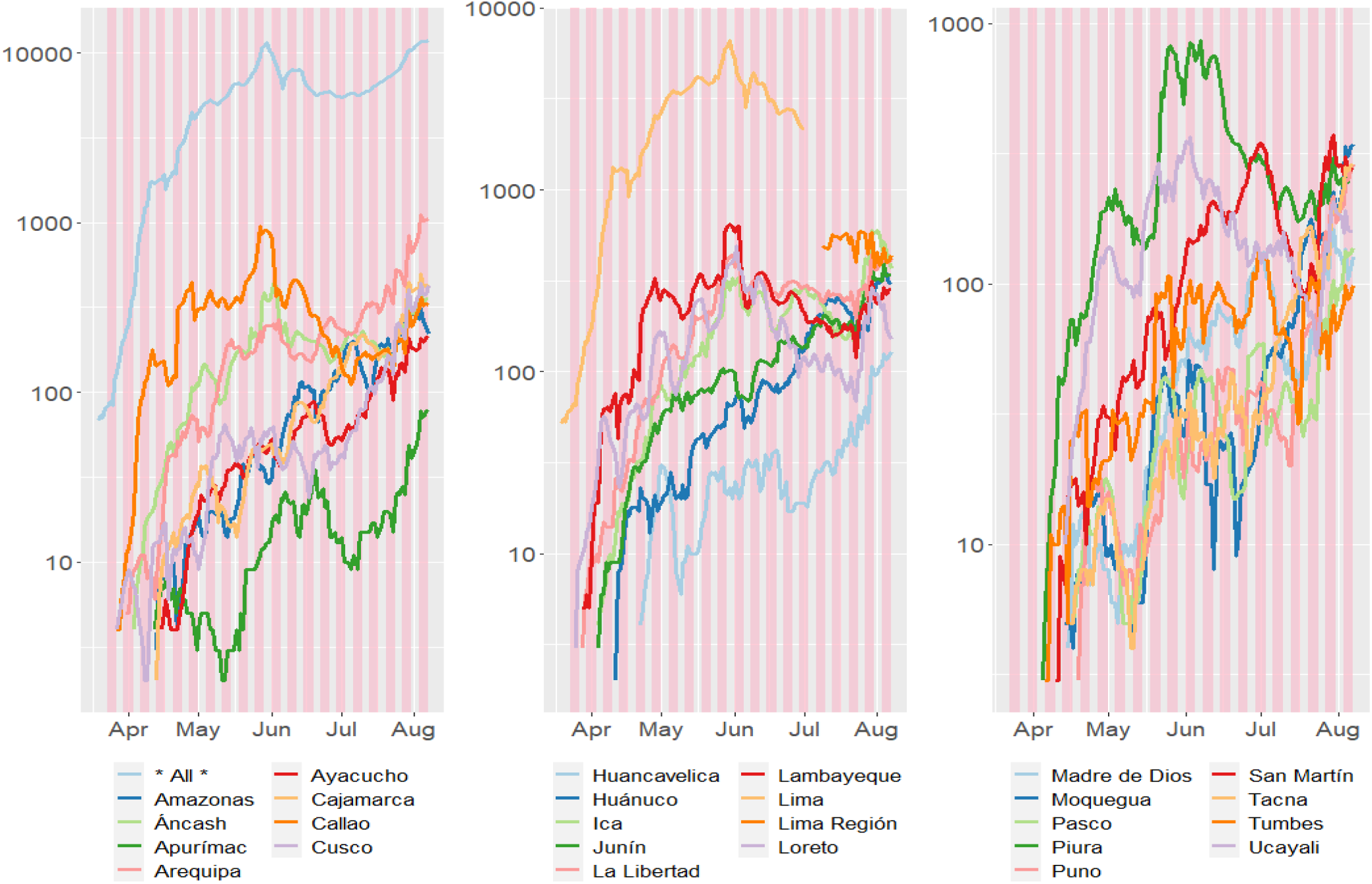
COVID-19 incidence in several Peruvian regions.

**Figure 31:**
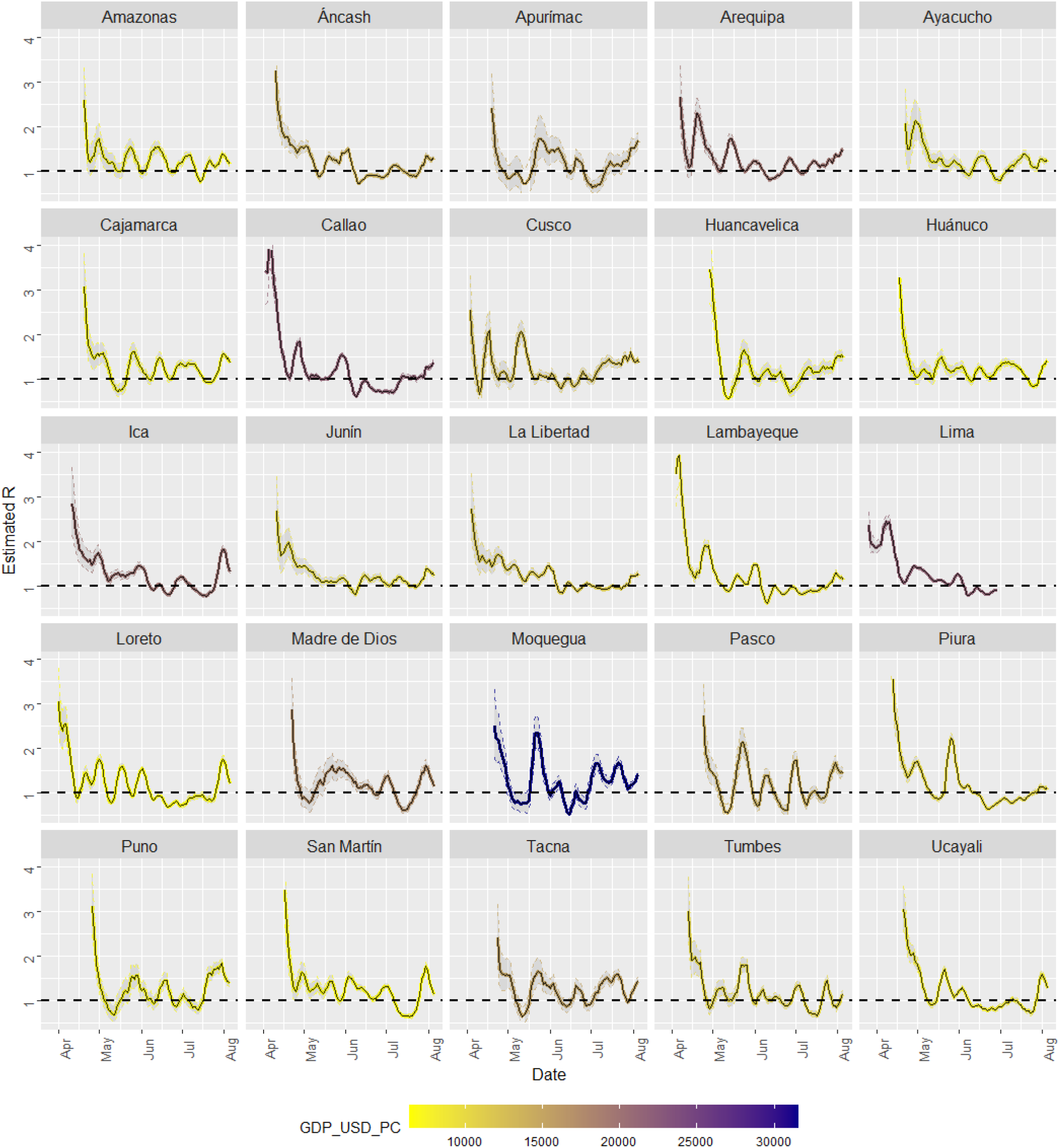
R_t_ estimates for Peruvian regions.

In regions with smaller GDP per capita the epidemic apparently started later.

When compared to the regions in developed countries (Spain, Belgium), the regions in Perú show a marked pattern of variability (more peaks and valleys, maybe due to smaller incidences) and more difficulty reaching control; similarly to the infection curves in other developing countries. Many regions exhibit peaks, but not all of them had the same impact on the incidence: The Moquegua and Pasco peaks reflected an increase from around 5 reported cases per day to 20 reported cases per day; the Piura peak was caused by an increase from less than 200 cases per day to 500 cases per day. This information should be interpreted with caution; the daily reported incidence could be influenced by testing capacity. The subtle pattern seen in USA states (wealthier states have more cases but better epidemic control) is not clearly observed here.

Overall, both regions in USA and regions in Perú follow a similar pattern, and the GDP per capita was not associated or mildly associated to differences the transmission curves.

### 5.13 Question G: Implications for reopening (Germany)

In Germany, in late June, after the reopening, several cases took place in clusters and the authorities managed them with a containment policy: In the middle-late of June an outbreak took place in an apartment building in Berlin(82), requiring quarantine measures. On the 24th of June the districts of Warendorf and Gütersloh in the state of North Rhine-Westphalia were put in lockdown following an outbreak in a meat processing plan(83). The purpose of section is to evaluate whether these outbreaks were reflected in the *R_t_* of affected areas and whether non-affected areas showed any *R_t_* impact. By using the Germany COVID-19 dataset compiled by J Gehrcke from the information provided by the Robert-Koch-Institute(84), *R_t_* estimates from 20-March-2020 to June have been obtained, for the districts of Giitersloh, Warendorf, the state of Westphalen, three “randomly” selected districts (Eisenach, Leipzig and Weimar), the state of Berlin and the state of Brandenburg. The preprocessing parameters include centerd SMA smoothing with a smoothing span of 4. Figure 32 shows the epidemic curves, figure 34 shows the transmission curves obtained the date before these measures were implemented and figure 33 shows the transmission curves obtained several weeks later.

**Figure 32:**
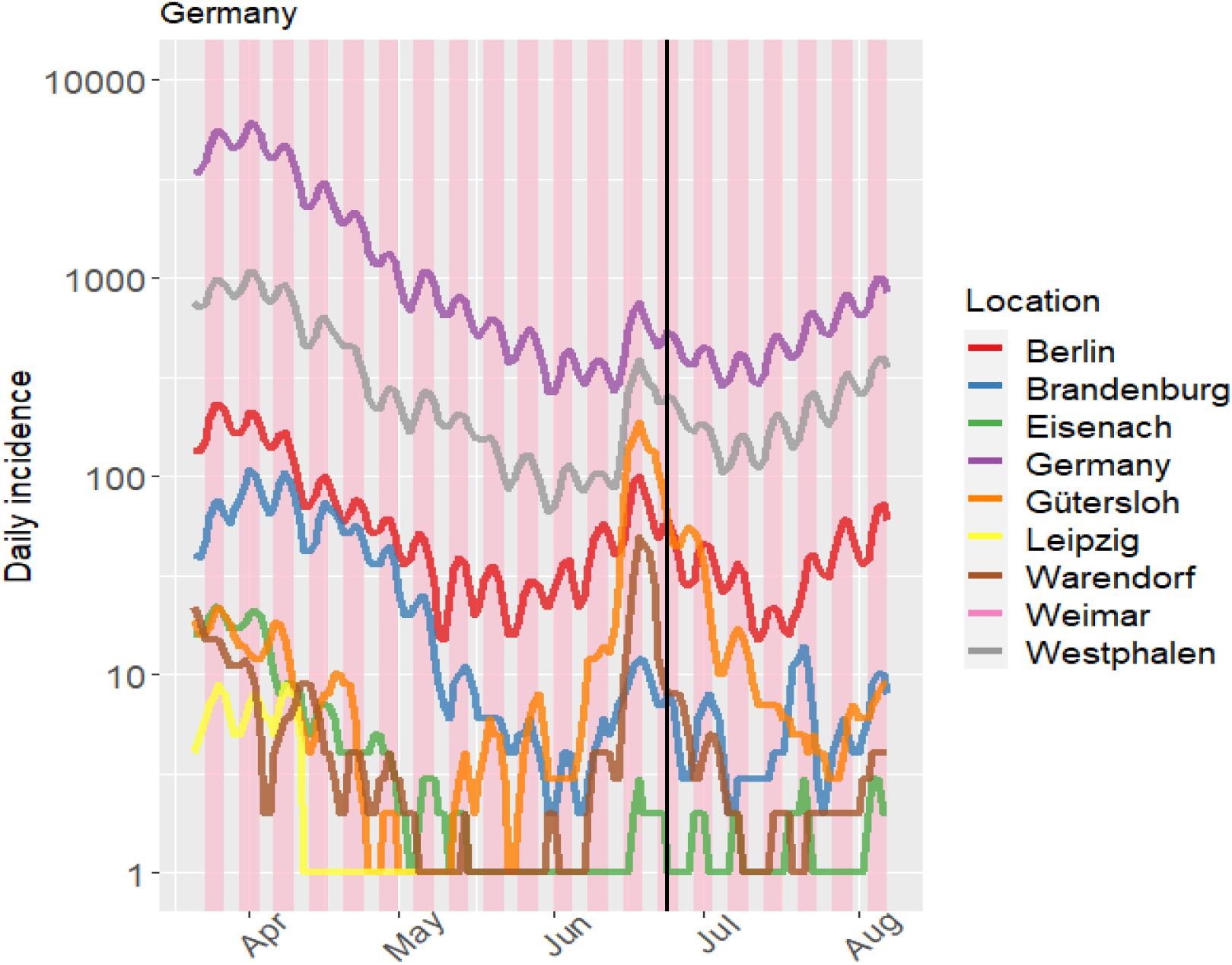
Epidemic curves of Germany.

**Figure 33:**
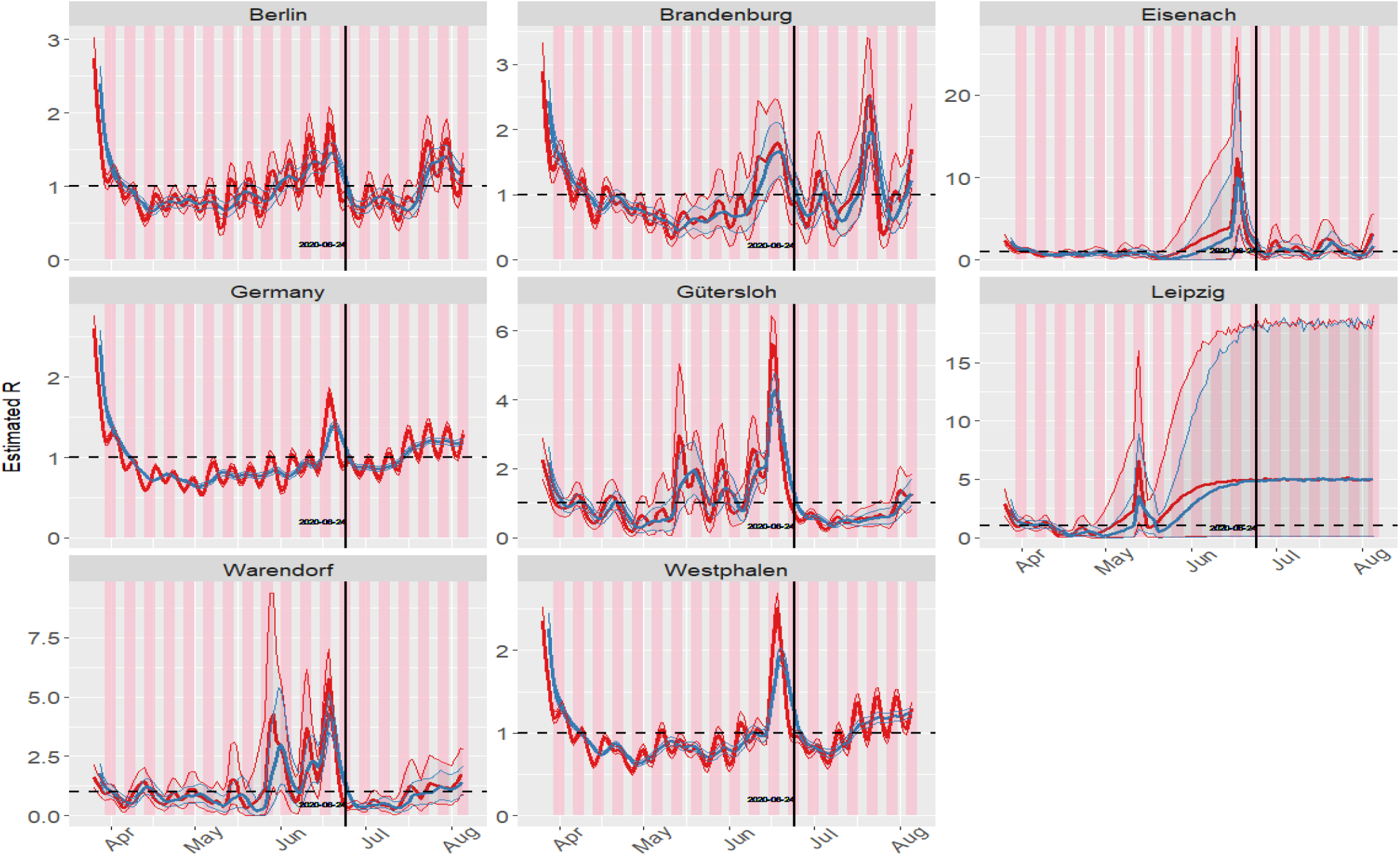
Transmission curves of Germany.

**Figure 34:**
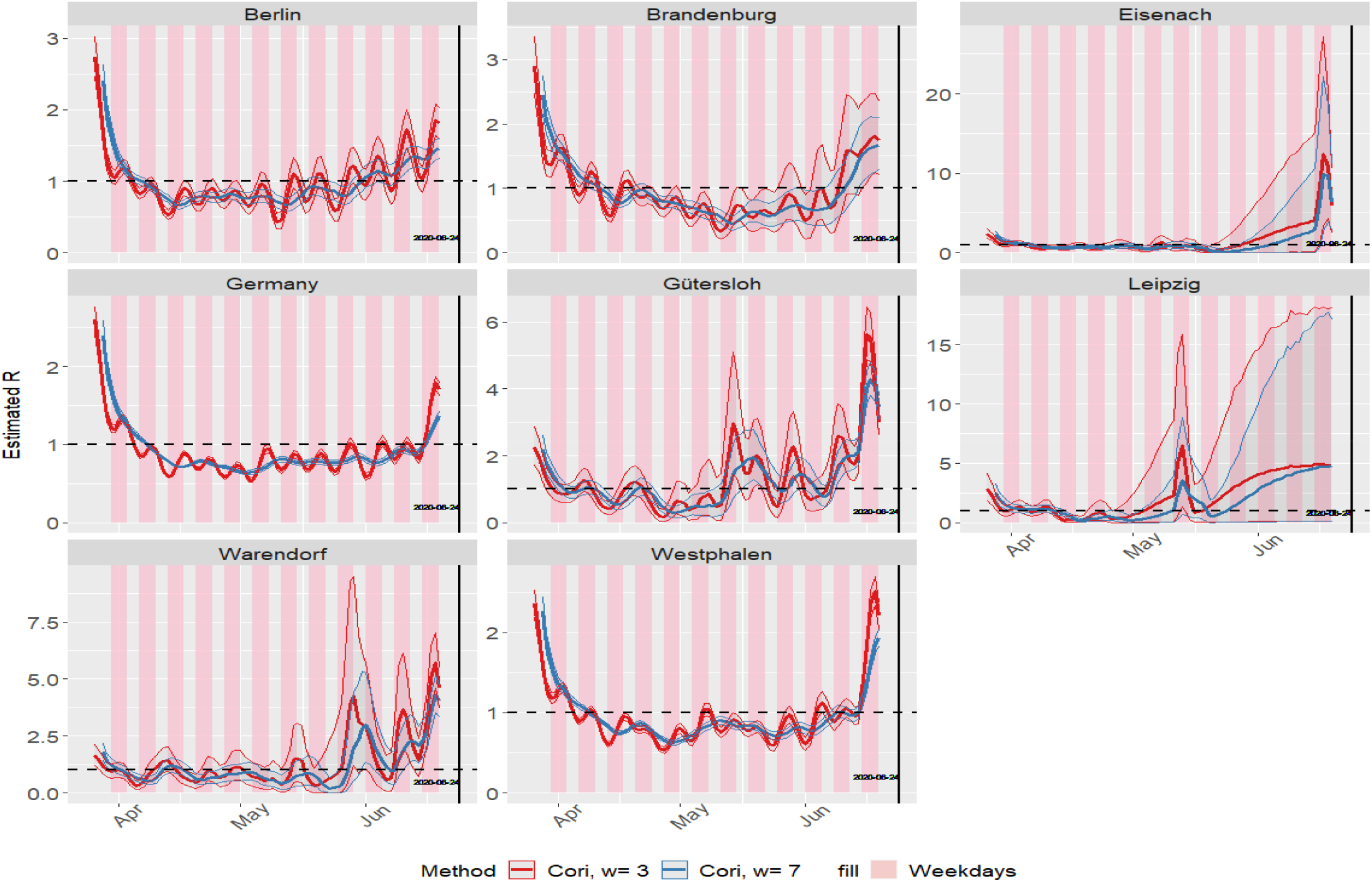
Transmission curves of Germany estimated one day before measures were implemented.

We can observe the scale of the epidemic is one-two orders of magnitude higher in states than in districts; the incidence in districts might be too low to obtain *R_t_* estimates.

The transmission curves show a significant increase in the districts of Gütersloh and Warendorf and even in the state of Westphalen in late June. An increase was also observed in Berlin and in Germany. On the other hand, these curves exhibit a non-significant increase in Eisenach, Leipzig and Weimar; this could be caused by a few “imported” cases. Three-day windows did not help to detect the outbreaks and showed more noise. Thus a surge in the epidemic curves accompanied by a significant increase in the transmission curves could be used as a signal to detect outbreaks; although we can also observe that “hindsight is always clear”, and the real-time transmission curves are not as easy to evaluate.

### 5.14 Question C, G: Implications for reopening (outbreaks in Catalonia)

On 04-July-2020 in the county of Segrià in Lerida, Catalonia, Spain, was quarantined (“cordon sanitaire”), following a surge in the number of cases(85); this comarca neighbours the Autonomous Community of Aragón, where an outbreak was taking place(48). Using the Official Catalonia Basic Health Areas dataset(45), *R_t_* have been obtained for several Basic Health Areas in Catalonia, using several *si/GT* distributions and keeping preprocessing settings constant (centered SMA smoothing with a span of 4 days, backwards distribution of negative values, no modification of spurious peaks, no correction for undetected cases). Figure 35 displays the incidences in different Basic Health Areas, with the dates of the end of the national State of Alarm and the start of the “cordon sanitaire”, and figure 36 displays the transmission curves. The date when the state of alarm ended (2020-June-21) and the day when new travel restrictions were issues have been marked.

**Figure 35:**
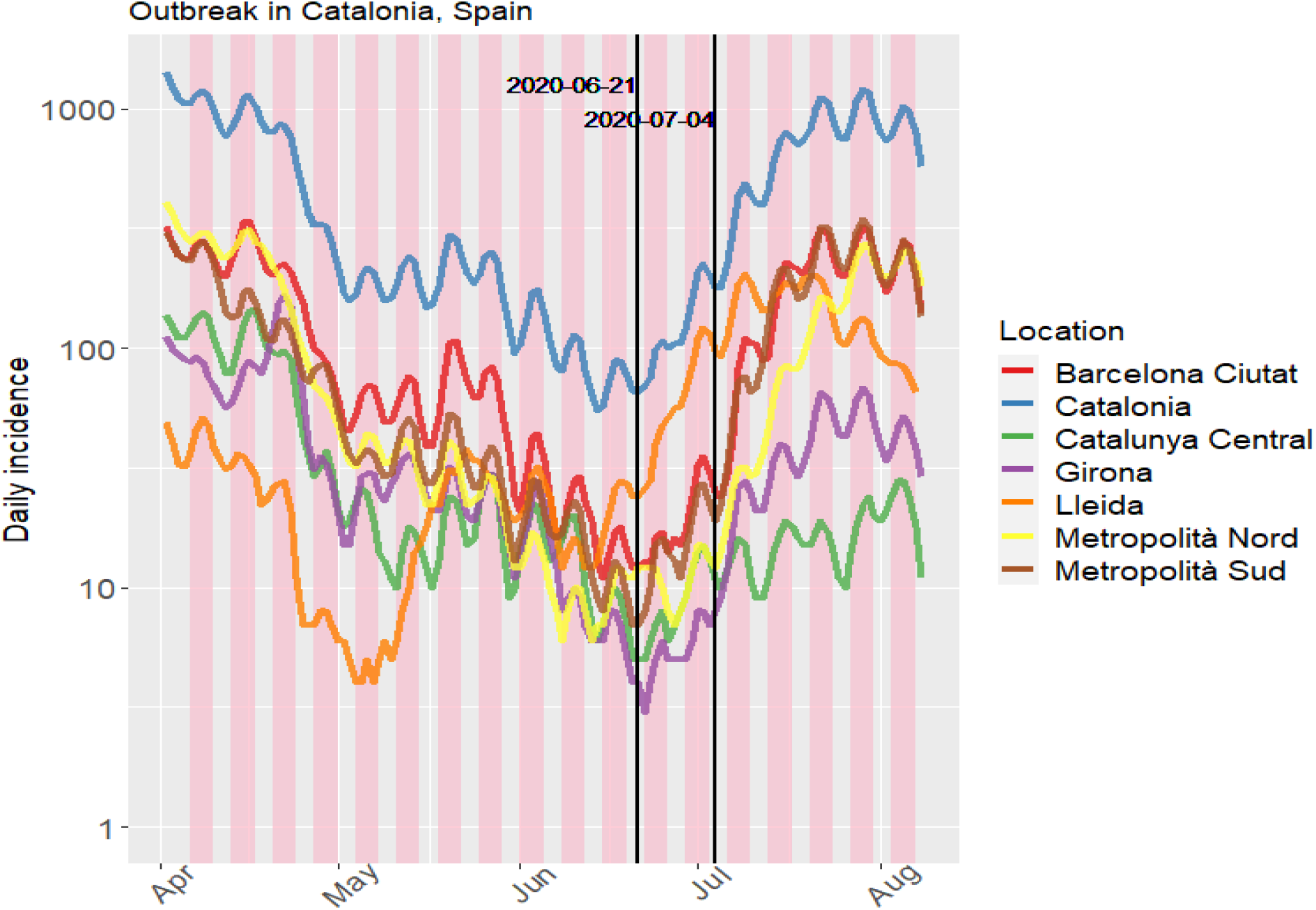
Epidemic curves of Catalonia.

**Figure 36:**
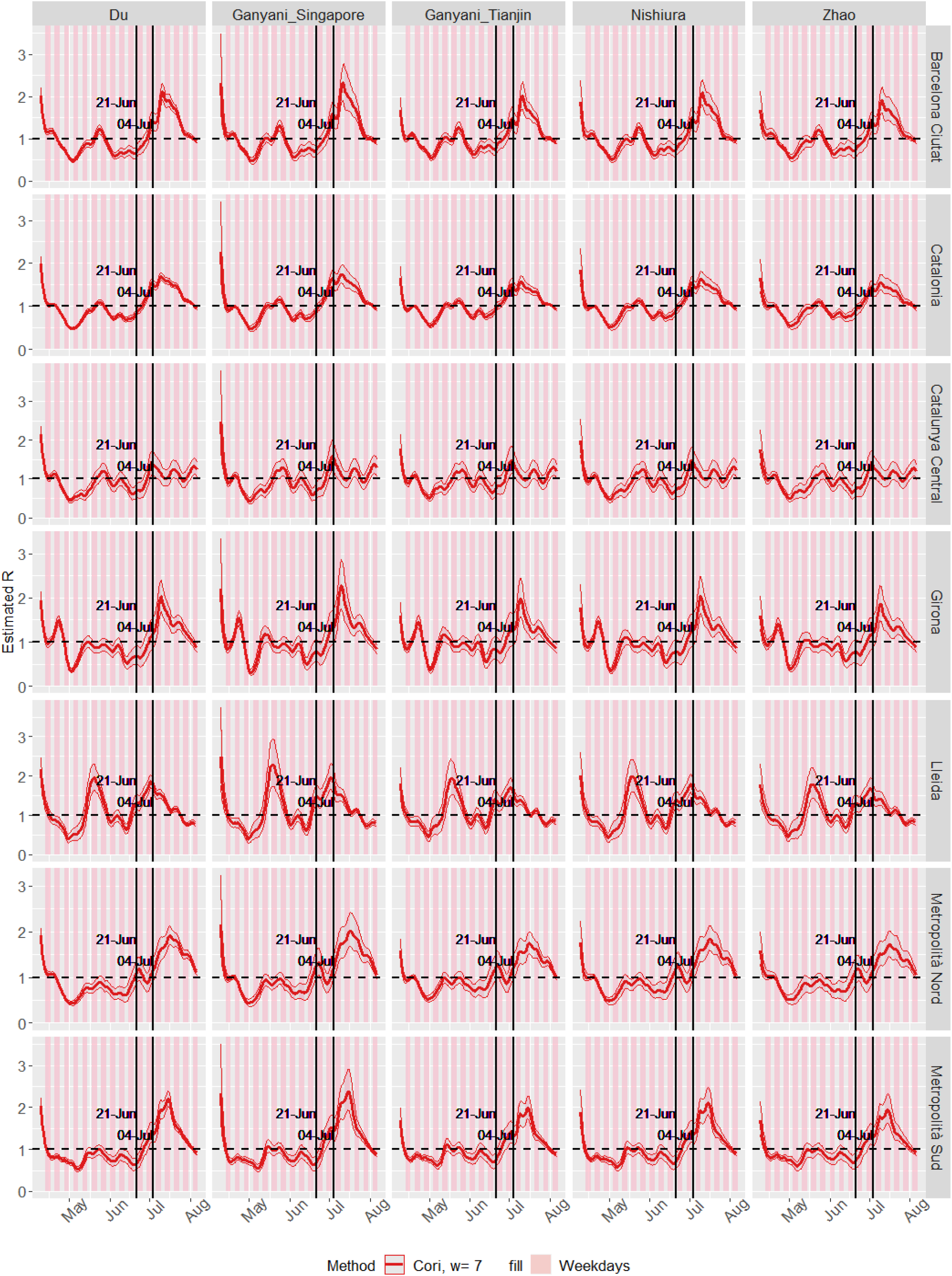
Transmission curves of Catalonia.

Although the initial outbreak of Coronavirus in March-April had a more pronounced in number of cases in regions with more population (Barcelona Metropolitan Area), the outbreak in the end of June/beginning of July affected the less populated Area of Lleida.

Two *R_t_* surges can be seen in the Lleida area: One around May, in which daily incidences rose from less than 10 cases/day to 40-50 cases/day, and one in July, in which daily incidences rose from around 10-20 cases day in mid June to 80-90 cases/day at the end of June. Therefore, the information provided by the *R_t_* needs to be complimented by the incidence information. This confussion can be solved by using the “overall infectivity” magnitude, as seen in figure 37. Overall infectivity at time t is defined as the sum of all the previously infected individuals, multiplied by their infectivity at time t (given by the serial interval distribution)(24). Overall infectivity has been calculated by using the mean of the 250 *si* or *GT* distribution samples obtained via MCMC or sampling (section 2).

**Figure 37:**
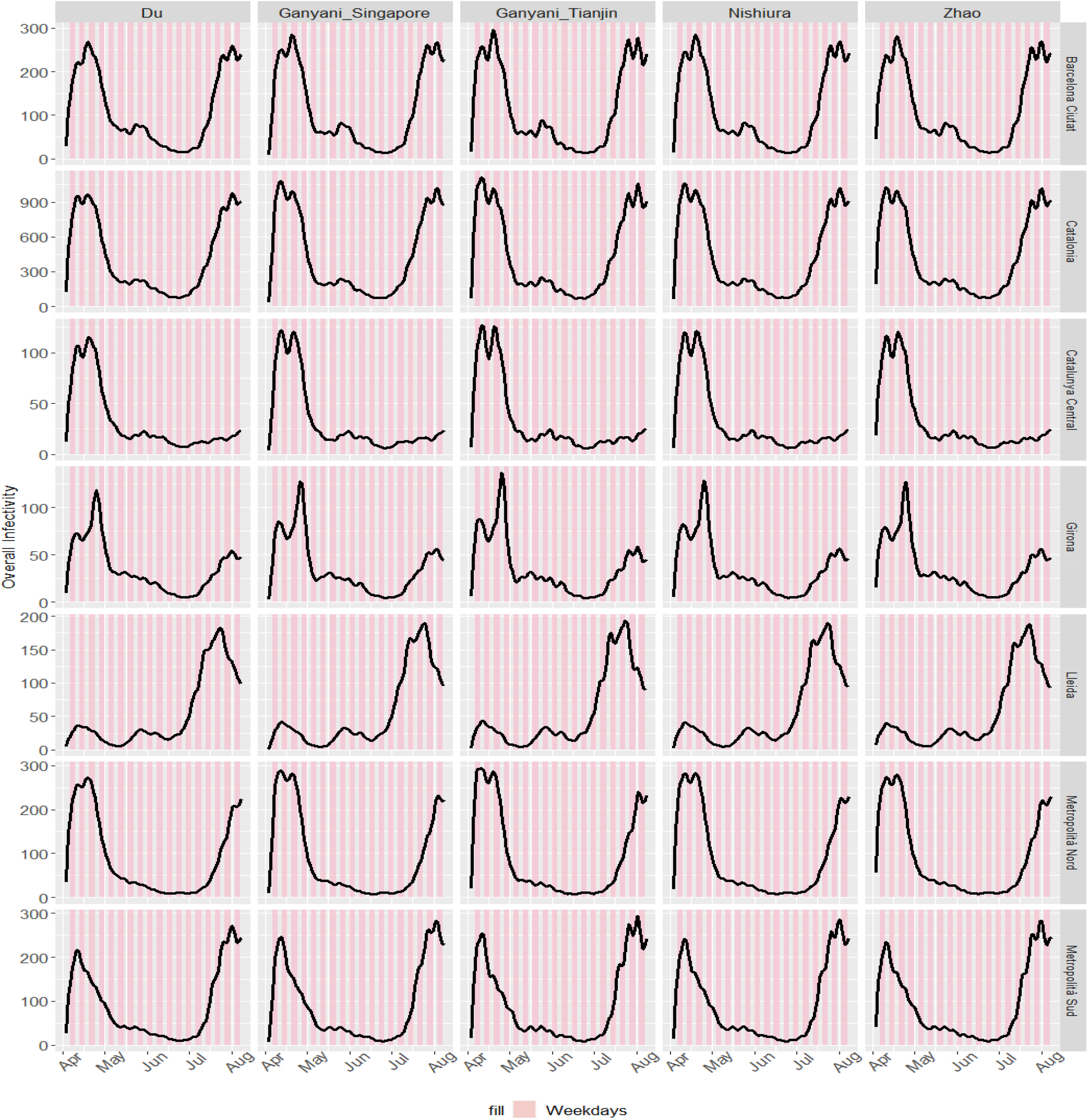
Overall infectivity curves of Catalonia.

Overall infectivity can therefore provide an accurate picture of outbreaks. It mostly follows the pattern of daily incidences epidemic curves. But compared to daily incidences, it is less influenced by weekday bias, but it might be influenced by the *si* or *GT* distribution, albeit very slightly.

## Data Availability

All data used here is already publicly available.

https://alfredob.shinyapps.io/estR0/

